# Public Health and Health Systems Impacts of SARS-CoV-2 Variants of Concern: A Rapid Scoping Review

**DOI:** 10.1101/2021.05.20.21257517

**Authors:** J Curran, J Dol, L Boulos, M Somerville, H McCulloch

## Abstract

**Background:** As of April 2021, three SARS-CoV-2 variants of concern (VOC: B.1.1.7, B.1.351 and P.1) have been detected in over 132 countries. Increased transmissibility of VOC has implications for public health measures and health system arrangements. This rapid scoping review aims to provide a synthesis of current evidence related to public health measures and health system arrangements associated with VOC.

**Methods:** Rapid scoping review. Seven databases were searched up to April 7, 2021 for terms related to VOC, transmission, public health and health systems. A grey literature search was conducted up to April 14, 2021. Title, abstracts and full text were screened independently by two reviewers. Data were double extracted using a standardized form. Studies were included if they reported on at least one of the VOC and public health or health system outcomes.

**Results:** Of the 2487 articles and 59 grey literature sources retrieved, 37 studies and 21 guidance documents were included. Included studies used a wide range of designs and methods. Most of the studies and guidance documents reported on B.1.1.7, and 18 studies and 4 reports provided data for consideration in relation to public health measures. Public health measures, including lockdowns, physical distancing, testing and contact tracing, were identified as critical adjuncts to a comprehensive vaccination campaign. No studies reported on handwashing or masking procedures related to VOC. For health system arrangements, 17 studies were identified. Some studies found an increase in hospitalization due to B.1.1.7 but no difference in length of stay or ICU admission. Six studies found an increased risk of death ranging from 15-67% with B.1.1.7 compared non-B.1.1.7, but three studies reported no change. One study reported on the effectiveness of personal protective equipment in reducing VOC transmission in the hospital. No studies reported on screening staff and visitors, adjusting service provisions, or adjusting patient accommodations and shared spaces, which is a significant gap in the literature. Guidance documents did not tend to cite any evidence and were thus assumed to be based on expert opinion.

**Conclusion:** While the findings should be interpreted with caution as most of the sources identified were preprints, findings suggest a combination of non-pharmaceutical interventions (e.g., masking, physical distancing, lockdowns, testing) should be employed alongside a vaccine strategy to improve population and health system outcomes. While the findings are mixed on the impact of VOC on health system arrangements, the evidence is trending towards increased hospitalization and death.

## Introduction

The SARS-CoV-2 virus, responsible for COVID-19, was declared a global pandemic by the World Health Organization (WHO) in March 2020.^1^ By April 2021, over 143 million cases of COVID-19 had been reported worldwide according to Johns Hopkins University,^2^ with 4.5 million new cases identified in the first week of April alone.^3^ A further three million people have died as a result of COVID-19 since the start of the pandemic.^2^ Increased numbers of COVID-19 cases is causing significant concerns around identifying and enforcing public health measures to control the spread of the virus and ensuring health systems can manage current and new admissions.

So far, three variants of the original SARS-CoV-2 lineage were declared variants of concern (VOC) by the WHO, with other variants under ongoing assessment.^4^ VOC are defined by their increased potential for transmission, presence of genomic mutations and rapid spread across countries or regions leading to possible decreased effectiveness of public health measures.^5^ In December 2020, the B.1.1.7 VOC (201/501.1.V1 or 2020/12/01) was first identified in the United Kingdom (UK)^6^ and as of April 13, 2021, 132 countries had reported cases of the B.1.1.7 variant.^3^ A second VOC was identified in South Africa (SA), known as B.1.351 (20H/501Y.V2) and has since been identified in 82 countries,^3^ while the P.1 VOC (previously known as B.1.1.28.1) which originated in Brazil, has been identified in 52 countries.^3^ While evidence is continuing to emerge on the impact of the circulating VOC on population health and health systems arrangements, early data suggests an increased risk of transmission associated with all three VOC.^3, 7–9^ Specifically, B.1.1.7 is estimated to be between 43-90% more transmissible than non-VOC,^3, 7, 8^ while B.1.351 is between 1.5^3, 10^ and 2.57 times more transmissible than non-VOC. There is limited evidence on the transmissibility of P.1, but early trends suggest it also has transmission advantage over non-VOC.^3, 7, 9^ Clearly, these circulating VOC present a risk to public health and safety.

The increased transmissibility of VOC has led to increase in surges in COVID-19 incidence and consequently, hospitalizations and mortality.^8^ The first wave of the pandemic demonstrated the potential for even well-equipped health systems to experience overwhelmed intensive care units (ICUs) and system disruption with wide ranging health consequences.^11^ For example, as of April 23, 2021, in Canada, cases of COVID-19 have been increasing and ICUs in some regions are at increasing risk of exceeding capacity to provide the usual calibre of critical care.^12^ However, due to the emergent nature of SARS-CoV-2 and the VOC, health systems and public health must make pragmatic decisions before evidence is available. This leaves many public health officials and healthcare administrators with uncertainty about the priority actions to minimize increased risk of spread of VOC and particularly whether there needs to be any existing modification to public health recommendations.^13^ There is also increasing pressure on the health system,^14^ despite a lack of evidence to inform measures needed to minimize the burden on the healthcare system. Therefore, this rapid scoping review aims to provide a synthesis of current evidence related to VOC in the context of public health and health system impacts. This review is a follow-up to the rapid scoping review on transmission conducted by this team.^9^

### Objective

To identify, appraise and summarize evidence related to the following questions about public health and health system impacts of the three major SARS-CoV-2 VOC as known in April 2021 (B.1.1.7, B.1.351, and P.1):

1. What is known about the implications of the three priority VOC for public-health measures on:

a. Modifying approach to vaccination (e.g., using vaccines that offer greater protection against variants, using different vaccines for first and second doses and/or re-vaccinating those initially vaccinated with vaccines with limited efficacy for new strains)
b. Modifying infection-prevention (i.e., public-health) measures in the community (e.g., changing duration of hand washing; changing mask type and characteristics, double masking, or other changes to masking; and changes to physical and temporal distancing)
c. Modifying infection-control procedures, such as:

▪ Changing duration for quarantining of exposed or potentially exposed individuals
▪ Changing duration for isolating suspected or confirmed cases (e.g., for exposed health workers)
▪ Changing testing strategy, including approach to testing, frequency of testing, and turn-around time for test results
▪ Changing approach to contact tracing
▪ Changing approach to outbreak management
2. What is known about the implications of the three priority VOC for health system arrangement (particularly for hospitals) on:

a. Adjusting capacity planning to accommodate changes in the risk of re-infection and the risk of severe disease (e.g., hospitalization, admission to ICU, and death)
b. Adjusting personal protective equipment (PPE) procedures for health workers
c. Adjusting restrictions and screening of staff and visitors (e.g., visitor policy changes, approach to and frequency of screening)
d. Adjusting service provision (e.g., cohorting patients in hospitals based on the VOC they have)
e. Adjusting patient accommodations, shared spaces, and common spaces (e.g., improvement to HVAC (heating, ventilation and air conditioning) systems)

### Design

Rapid scoping review, following standardized rapid and scoping review guidelines.^15–17^ This review will be updated in June 2021; the most up-to-date version will be listed on the COVID-END website.

## Methods

A broad, comprehensive search was designed by an information specialist to retrieve all literature related to VOC. The electronic database search was executed on March 15, 2021 and again on April 7, 2021 in MEDLINE (Ovid MEDLINE All), Embase (Elsevier Embase.com), the Cochrane Database of Systematic Reviews (CDSR) and Central Register of Controlled Trials (CENTRAL) (Cochrane Library, Wiley), Epistemonikos’ L·OVE on COVID-19, and medRxiv and bioRxiv concurrently. The electronic database search was followed up by a grey literature search, executed on March 18-19, 2021 and again April 12-14, 2021, using a list^18^ of specific COVID-19 resource websites in addition to broader searches of Google and Twitter. Only English-language searches were conducted, but non-English results were considered for inclusion. Full search details are available in Appendix 1.

Evidence specific to public health and health system arrangement impacts were identified and tagged during the screening process related to any of the protocol questions. Studies that reported on immune escape (vaccine/prior infection protection), non-VOC impacts, testing approaches, transmission, case studies without public health or health system impacts, or animal studies were excluded. Reviews, overviews, and news articles that presented no original data were excluded but checked for references to primary studies.

Title/abstract and full-text screening was completed by two reviewers in Covidence. The data extraction form was designed in consultation with knowledge user partners; data were extracted by two reviewers and verified by a third. The final report was reviewed by health system and infectious disease experts engaged on our team.

Quality appraisal was conducted using the Newcastle-Ottawa scale (NOS)^18^ and the AGREE II tool.^19^ Case-control or cohort design studies were assessed using the NOS, cross-sectional studies were assessed using the adapted NOS,^20^ and guidance documents were assessed using AGREE II. Two team members independently conducted quality appraisal for all eligible studies. Reviewers met to discuss scores and a third, independent team member was consulted to assist with resolving conflicts. Modeling studies, lab-based studies and other grey literature sources were not appraised.

Cohort studies were awarded a maximum of nine stars and cross-sectional studies awarded a maximum of 10 stars, based on three scoring categories: selection, comparability, and outcome. Two stars were subtracted from pre-print studies as an added layer of quality assessment due to the emerging nature of studies on this topic. Final scores for observational studies were presented as a percentage, based on an average between the two appraiser scores. An overall quality rating of low, medium or high was reported for each observational study, which correlated with a score of <50%, 50-80% or >80% respectively.

Guidance documents were awarded a maximum of 161 points on the AGREE II tool, by scoring quality on a scale of 1-7 across 23 separate items within six domains. Guidance documents were scored following the AGREE II formula for each domain: [(Obtained score - Minimum possible score)/(Maximum possible score - Minimum possible score)] x100%. This scaled domain score was presented for each of the six domains. The same formula was used to calculate the final overall assessment score, where each appraiser gave an overall rating from 1-7 and indicated whether they would recommend the guideline. An overall quality rating of low, medium, or high was given based on a function of the overall guidance score and decision to recommend the guidance document.

## Results

The search retrieved 2487 electronic database records and 59 grey literature records, of which 37 studies and 21 guidance documents were included (see Appendix 2 for PRISMA Flow Diagram). Of the 37 studies identified, 25 were preprints, eight were published in peer-reviewed journals, and four were reported in grey literature sources (see Appendix 3 – Tables 1 and 2 for a summary table of included studies). Three sources reported solely on P.1, one source reported on B.1.351, 25 sources reported on B.1.1.7, six reported on all three, one on B.1.1.7 and B.1.351, and one reported on a non-specific VOC. There was a wide variation in countries, including the UK (n=12), the United States (US) (n=4), France (n=3), Brazil (n=3), Canada (n=2), Germany (n=2), Israel (n=2), and one each from Denmark, Italy, Lebanon, the Netherlands, Portugal, and South Africa. Three studies reported on multiple countries.

Of the 21 guidance documents, most discussed all VOC, except for one that focused solely on B.1.1.7 and two that focused on B.1.1.7 and B.1.351 (Appendix 3 – Table 3 summary of guidance documents). Most of the guidance documents originated from public health agencies or health authorities within Canada (n=14), with others from the UK (n=3), Ireland (n=1), the US (n=1), Europe (n=1) and international (n=1).

### A note about the guidance documents included in this review

- The guidance documents included in this rapid scoping review are not the result of a comprehensive jurisdictional scan. They were retrieved via Google using a general search for keywords related to VOC. As a result, the guidance featured throughout this review is under-representative of Canadian provinces whose guidance was not specific to VOC at the time of the search (e.g., Atlantic provinces where VOC may not have been identified or prevalent at the time), or whose guidance was not optimally indexed for Google searching. A comprehensive jurisdictional scan, including hand-searching of provincial websites and consultation with personal contacts, will be produced in an upcoming separate report.

### Critical Appraisal

Of the 33 included non-grey literature studies (preprints and peer-reviewed), 11 were cohort studies and five used a cross-sectional design, and thus subject to appraisal using the NOS. Cohort studies scored six to nine stars out of a possible nine, which is a 67-100% in overall quality. Cross-sectional studies scored one to eight stars out of a possible 10, with a range of 10-80% for overall quality. Four studies scored 80% or higher,^24–27^ indicating high quality. The majority (n=8) scored 50-80%,^28–35^ suggesting medium quality, while four studies were considered low quality, scoring 10-44%.^36–39^ Twelve of the 33 studies were pre-prints, meaning they had not yet been peer reviewed. As the quality of preprints should be interpreted with caution, efforts were made to reflect this in the overall score through the removal of two points. A complete overview of NOS scores by study can be found in Appendix 4. Of note, four studies were laboratory-based and 13 were epidemiological modeling studies and were therefore not included in the quality assessment.

Of the 25 grey literature resources included in this review, 21 were guidance documents and appraised using AGREE II. The overall quality scores of included guidance documents ranged from 16.7% to 83.3%, indicating a range of low quality to high quality. However, it is important to note that no guidance documents included in this review were clinical practice guidelines, but rather public health directives presented in a variety of formats. A complete overview of guidance document scores based on the AGREE II tool can be found in Appendix 4.

#### Question 1: Public Health Measures

##### Question 1A

Modifying approach to vaccination (e.g., using vaccines that offer greater protection against variants, using different vaccines for first and second doses and/or re-vaccinating those initially vaccinated with vaccines with limited efficacy for new strains).

Seven studies contributed data which may be relevant to modifying the current approach to vaccine scheduling and delivery (see Table 1). Of the seven studies, 5 were modeling^40–42^ or lab-based studies^43, 44^ which were not critically appraised. The two studies that were critically apprised were of low quality.^36, 37^

**Table 1.**
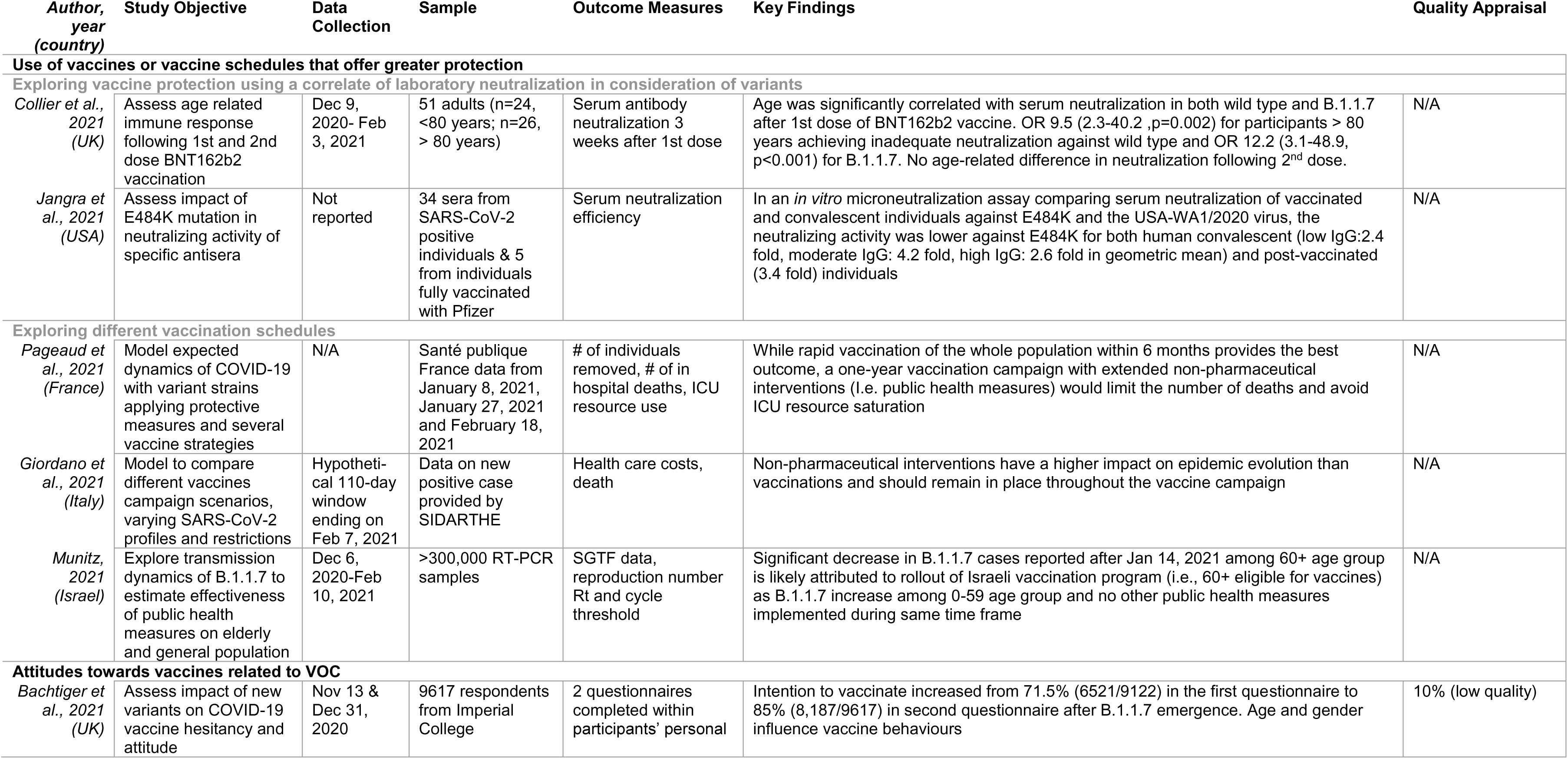
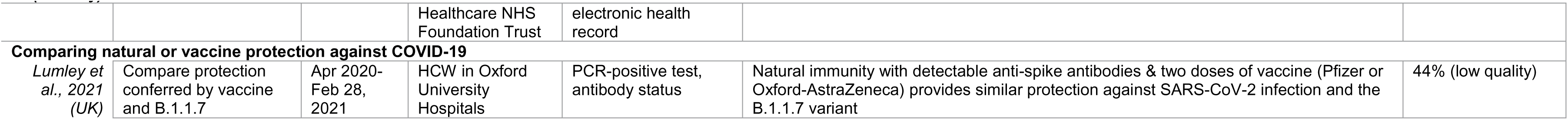
Study summary on findings related to modifying approach to vaccination, categorized by study topic

**Key findings for consideration include:**

▪ Age appears to be a factor in immune response after the first dose of mRNA-based vaccines
▪ Optimal vaccine schedules combined with non-pharmaceutical interventions (i.e., restrictions, masks, physical distancing) is expected to limit the number of COVID-19 related deaths and preserve ICU capacity
▪ Age and gender may influence response to public health messages regarding vaccine uptake

Four guidance documents (two low-quality,^45, 46^ one medium-quality,^47^ and one high-quality^48^) contained information related to vaccination approaches, but none explicitly recommended changing to vaccines or vaccination schedules that offer greater protection, using different vaccines for first and second doses, or re-vaccinating. It is acknowledged that current guidance is predicated on a rapidly changing evidence base.

###### Use of vaccines or vaccine schedules that offer greater protection

A total of five studies provide data which may be useful to consider when designing how vaccines are rolled out with the emergence of the variant strains. All five studies were modeling or lab-based studies and thus were not critically appraised.

**<u>Exploring vaccine protection using a correlate of laboratory neutralization in consideration of variants</u>**

Two small studies provided data related to serum neutralization protection against VOC. Collier et al. conducted a prospective cohort study in the UK including 51 participants (median age 81 years; n=24, <80 years; n=27 > 80 years), to assess immune response following the first and second dose of mRNA-based vaccines.^43^ Vaccine elicited serum antibody neutralization was measured as a dilution of serum required to inhibit infection by 50% in an *in vitro* neutralization assay at least 3 weeks after the first dose of vaccine. Age was found to be statistically correlated with serum neutralization in both the wild type and B.1.1.7 after the first dose. The adjusted odds ratio (OR) for participants 80 years and older versus younger than 80 for achieving inadequate neutralization against wild type was 9.5 (2.3-40.2, p=0.002) and against the B.1.1.7 variant was 12.2 (3.1 – 48.9, p<0.001). Re-testing 3 weeks after the second dose showed no age-related differences in neutralization activity.

Jangra et al. examined the impact of the E484K mutation on the neutralization activity of SARS-CoV-2 specific antisera using a sample of 34 sera from SARS-CoV-2 positive individuals and sera from 5 individuals who were fully vaccinated with the Pfizer vaccine.^44^ *In vitro* microneutralization were performed in a blinded manner with both the USA-WA1-2020 virus (similar to strains in the early phase of the COVID-19 pandemic) and an identical recombinant SARS-CoV-2 except for the E484K mutation on the spike receptor binding domain. Serum neutralizing activity of human convalescent and post-vaccinated donors was significantly lower against E484K (convalescent low IgG: 2.4-fold, moderate IgG: 4.2-fold, high IgG: 2.6-fold; vaccinated samples: 3.4-fold based on geometric means) when compared with USA-WA1-2020.

**<u>Exploring different vaccination schedules</u>**

Two studies modeled the impact that changes in the vaccine scheduling would have on VOC. Pageaud et al. used a stochastic agent-based model (ABM), stratified by age, which considered the influence of the variant strains, three different non-pharmaceutical intervention (NPI) protocols (relaxed, intensive, extended), and four different vaccine schedules (6,12, 18, 24 months) to examine impact on number of cases, deaths, hospitalizations and ICU resource use.^41^ A 6-month vaccination campaign with an intensive-NPI resulted in the least number of deaths (∼18000) and avoided ICU resource saturation. With a 12-month vaccine schedule, the number of deaths were 3 times higher and extended-NPI was needed to avoid ICU resource saturation. Vaccine campaigns up to 18 and 24 months would lead to 81 and 93 thousand deaths respectively and saturation of the ICU resources, even with intensive-NPI. In all models with vaccine schedules longer than 6 months, extended-NPI was needed to avoid ICU resource saturation.

Giordano et al. employed a SIDARTHE compartmental model using Italian field data to predict the impact of VOC based on various vaccination campaigns in Italy.^42^ Authors reported 20 unique scenarios associated with differing speeds of vaccine rollout, transmissibility profiles and public health measure strategies. Containment strategies (i.e., lockdowns, physical distancing) had a 5-fold impact on reducing human losses in the period of February 2021 to January 2022 in slow, medium and fast vaccine schedules indicating NPIs have a larger effect than vaccination speed. The model demonstrates that in consideration of the highly transmissible VOC, NPIs are crucial for controlling the epidemic. Preemptive strategies (first close when case numbers start to rise and then open at low case numbers) will reduce hospitalizations and deaths when compared with delayed interventions (keep open then close when case numbers start to rise to prevent ICU saturation). Early closures drastically reduce death and healthcare system costs compared with delayed closures.

One study, Munitz et al., provided real-world evidence of the effectiveness of a vaccine roll-out through reporting a correlation of Israel’s vaccination campaign with rates of variant B.1.1.7 using data from >300,000 RT-PCR samples collected between December 6, 2020, and February 10, 2021. ^49^ B.1.1.7 had become the dominant variant (92%) up to January 14, 2021 among all age groups (r>0.99). After January 14 (with 50% of 60+ age group receiving first dose of vaccine), there was a decline in the 60+ age group compared with individuals 0-19 years old (r=0.35) and 20-59 years old (r=0.28). A national lockdown implemented in January 2021 and a surveillance testing program in nursing homes and the community enabled early detection and helped to contain viral spread in at risk populations.

###### Different vaccines for first and second dose

No studies to date have reported on this outcome.

###### Attitudes towards vaccine related to VOC

Changes in COVID-19 vaccine hesitancy related to VOC emergence were assessed in the UK by Bachtiger et al. as part of an ongoing cross-sectional longitudinal study involving 18581 participants examining the effects on well-being of the COVID-19 pandemic.^37^ Study participants were invited to complete weekly surveys through their personal electronic health record, Care Information Exchange. Questionnaires related to vaccine behaviour were sent on November 13, 2020 (following Pfizer vaccine reported efficacy of >90%) and on December 31, 2020 (after first reports of B.1.1.7). Intention to receive the vaccine increased from 71.5% (n=6521 of 9122 participants) in the first questionnaire to 85.1% (n= 8187 of 9617 participants) in the second questionnaire. Three hundred seventy-five participants in the second questionnaire indicated they changed their minds to wanting vaccination considering news of the new VOC B.1.1.7. Yearly increase in age (adjusted-OR: 1.045 [95% confidence interval (CI):1.039-1.050]) and female gender (adjusted-OR: 0.540 [95%CI:0.461-0.632]) increased and decreased vaccine acceptance, respectively. This study was critically appraised as low quality, so these findings should be interpreted with caution.

###### Comparing natural and vaccine protection against COVID-19

Lumley et al. followed a sample of 13109 health care workers (HCW) from Oxford University Hospitals to determine the protection conferred following infection from B.1.1.7 and one and two doses of vaccines.^36^ HCW were offered asymptomatic nasal and oropharyngeal swab PCR testing every two weeks and serological testing every two months from April 2020 and the staff vaccination program began December 8, 2020. Data is reported up to February 28, 2021. Anti-timeric spike IgG ELISA was used to determine antibody status. The rates of PCR-positive tests were highest in the unvaccinated seronegative HCWs and 85% lower in unvaccinated seropositive HCWs (alRR=0.10 [0.08-0.26, p<0.001]). The incidence of any PCR-positive result was reduced by 64% in seronegative HCWs following first vaccination (alRR=0.36 [0.26-0.50; p<0.001]) and 90% following second vaccination (alRR=0.10 [0.02-0.38; p<0.001]). B.1.1.7 did not significantly alter the extent of protection for PCR positive infection in those who were seropositive (aIRR =0.40 [95%CI 0.10-1.64; p=0.20]) or following a first vaccine dose (aIRR=1.84 [0.75-4.49; p=0.18]). Overall, findings suggest immunity induced by natural infection with detectable anti-spike antibodies, including B.1.1.7, and vaccine is robust. This study was critically appraised as low quality, so these findings should be interpreted with caution.

###### Guidance documents related to vaccination approach

- As the influence of VOC on vaccine effectiveness remains under investigation and vaccines against VOC-specific mutations are in development but not yet available, no guidelines or guidance documents are able to explicitly recommend changing to vaccines that offer greater protection. Likewise, there was no guidance related to using different vaccines for first and second doses, or to re-vaccinating due to evolving studies in these areas.
- Instead, guidelines focused on vaccine approaches in general, offering scenario-based recommendations. If whole genome sequencing reveals that the effectiveness of vaccines against VOC is deteriorating,^48^ Health Canada suggests vaccines and vaccination approaches should be modified as quickly as possible.^45^ In countries in which the spread of the virus remains slow, jurisdictions may decide to begin by targeting at-risk groups, or by targeting “key transmitters”; however, in places where spread of VOC is rapid, targeting key transmitters becomes less feasible and less effective.^47^
- At the end of February 2021, the Center for Infectious Disease Research and Policy (CIDRAP) at the University of Minnesota offered the following suggestions to strategically deploy the vaccine supply in the US:^46^

○ Allocating vaccine with people ≥65 years given highest priority;
○ Deferring second doses of mRNA vaccines until after the surge;
○ Deferring the second dose of mRNA vaccines in people with confirmed COVID-19 infections;
○ Authorization and use of half-dose regimen for Moderna vaccine.
- Health Canada recognizes that to react quickly to changes in vaccine effectiveness against VOC, especially when little is known about a new variant, “harmonising all the vaccines on one or a few sequences may not be straight-forward, and vaccines with a variety of sequences, developed as quickly as possible by the manufacturers may be a pragmatic and rapid means of introducing updated vaccines at this stage in the pandemic. More sophisticated regulatory control could be introduced once the virus is better understood.”^45^

##### Question 1B

Modifying infection-prevention (i.e., public-health) measures in the community (e.g., changing duration of hand washing; changing mask type and characteristics, double masking or other changes to masking; and changes to physical and temporal distancing)

Six sources reported on identifying infection-prevention measures in the community, particularly around physical distancing (see Table 2). Four sources were modeling studies^40, 42, 50, 51^ and the other two were grey literature,^52, 53^ thus no quality appraisal was completed.

**Table 2.** Study summary on public health infection-prevention measures in the community

| **Author, year**<br>**(country)** | **Objective** | **Data collection**<br>**period** | **Sample** | **Outcome**<br>**measures** | **Relevant key findings** | **Quality**<br>**appraisal** |
| --- | --- | --- | --- | --- | --- | --- |
| *Borges et al., 2021*<br>*(Portugal)* | Investigate the proportion of SGTF cases to gain insight on B.1.1.7 frequency and spread in Portugal | Week 49, 2020 to week 3, 2021 | Data set from Portuguese National Institute of Health Dr. Ricardo Jorge Dec 2020 to Feb 5, 2021 | SGTF & SGTL test | Physical distancing measures (general lockdown) implemented in weeks 2 & 3 of 2021 decelerated the growth rate of SGTF positive cases | N/A |
| *Domenico et al., 2021*<br>*(France)* | Assess the impact of social distancing on historical and variant strain through modeling | N/A | Flash1 survey data from Santé publique France on Jan 28, 2021 | B.1.1.7 prevalence | Strong social distancing measures (e.g., curfews, lockdowns, work from home) including mild lockdown are needed to decelerate the surge of B.1.1.7 in the third wave | N/A |
| *Vazquez et al., 2021*<br>*(Germany)* | Estimate SARS-CoV-2 rate of transmission per proximity contact and generate a model to simulate infectious disease outbreaks in workplaces | Not reported | 605 individuals from one workplace | Proximity data between two coworkers tracked for 44 days through Bluetooth wearable devices | Using single case workplace proximity data and reproductive numbers for SARS-CoV-2 and B.1.1.7, the transmission rate per contact was determined to be 3 times higher for B.1.1.7 (0.041) vs SARS-CoV-2 (0.014). Workplaces can use proximity data to simulate disease outbreaks and management strategies | N/A |

**Key findings for consideration include:**

▪ Evidence supporting modification to infection-prevention measures related to VOC is sparse, particularly related to hand washing and masking protocols
▪ Physical distancing and other non-pharmaceutical interventions are important in reducing the spread of VOC in the community
▪ Opportunities exist to expand evidence related to infection-prevention measures in the workplace, particularly in the presence of more highly transmissible VOC

Five guidance documents (low to medium quality) offer recommendations on infection prevention measures in the community, encompassing general community settings, personal services such as hair salons, and the retail sector.^54–57^ None of these guidance documents cited supporting evidence, suggesting expert consensus was used to derive recommendations.

###### Hand washing and mask protocols

We did not identify any published or preprint studies relevant to modifying hand washing or mask protocols related to the variants of concern. However, a Public Health Ontario Environmental Scan identified 3 out of 14 jurisdictions reviewed had changed the type of mask recommended in public in response to the VOC (from cloth masks to medical masks or respirators).^53^ No evidence was identified or cited from these jurisdictions to support the recommended change. A Centers for Disease Control and Prevention communication from the Epidemiology Taskforce on the VOC indicates that current mitigation strategies including masking and hand washing work.^52^

###### Physical distancing

Three studies contributed data related to physical distancing in consideration of the VOC (Table 2). In a surveillance study evaluating the spread of B.1.1.7 in Portugal between December 20, 2020 and January 20, 2021, Borges et al. concluded that the physical distancing measures implemented in weeks 2 and 3 of 2021 strongly decelerated the growth of B.1.1.7.^50^ While models had forecasted the proportion of SGTF/SGTL cases to reach up to 68% (95% CI:65-71), the proportion of SGTF and SGTL positive cases remained below 50% until week 7 of 2021.

Domenico et al. used a discrete, stochastic model integrating demography, age profile, social contacts, and mobility data over time to model the impact of social distancing measures on two strains of SARS-CoV-2 (historical, B.1.1.7).^40^ Strain circulation dynamics for France, Ile-de-France regions and Nouvelle Aquitaine were secured through Santé Publique France on January 28, 2021. The model estimated that the progressive social distancing implemented in January 2021 brought the reproductive number of the historical strain below 1 but the B.1.1.7 cases increased exponentially with the estimated reproduction (R) about 1 in all three regions. The authors suggest that strengthening social distancing measures with the addition of restrictive measures such as weekend lockdown will be needed to decelerate the resurgence of B.1.1.7 in a third wave.

Vazquez et al. conducted a novel modelling study at the level of a workplace using co-worker proximity data gathered through Bluetooth technology.^51^ Proximity data, collected from button devices worn by 605 workers for a period of 44 days, provided a temporal network to model the spread of airborne viruses. This data is combined with an infection transmission model developed by the team to estimate the SARS-CoV-2 transmission rate per proximity contact. Social distancing is modelled by removing different fractions of the proximity contacts. Based on the proximity data from the sample workplace, the model estimated the transmission rate per proximity contact for SARS-CoV-2 cases as 0.014 and B.1.1.7 as 0.041 per proximity contact, approximately 3 times higher. While the introduction of infection-prevention measures such as social distancing and mask wearing reduces the infection rate, B.1.1.7 transmissibility remained 2 times larger than the wild-type.

###### Guidance documents related to infection control in the community

- Hand washing guidelines remain relatively unchanged considering VOC. Mask use in all public places remains important. Multi-layer masks are recommended. Eye protection is now recommended in some circumstances in the community. Physical distancing remains important, especially in cases of VOC with higher viral load. In Ontario, infection control measures remain generally unchanged because of VOC. Cleaning of surfaces is still recommended by some guidelines. None of the guidelines cited in Table 3 cite any evidence. **Please note that this table is not representative of all provinces (see note about guidance documents included in this review, page 4).**

##### Question 1C

Modifying infection-control procedures, such as, changing duration for quarantining of exposed or potentially exposed individuals, changing duration for isolating suspected or confirmed cases (e.g., for exposed health workers), changing testing strategy, including approach to testing, frequency of testing, and turnaround time for test results, changing approach to contact tracing, changing approach to outbreak management.

**Table 3.** Summary of guidance documents from select jurisdictions on hand washing, masking, and physical distancing guidelines in community settings

|  | Hand washing | Mask protocols | Physical distancing | Other |
| --- | --- | --- | --- | --- |
| *In the community (general)* | Hand washing remains important.<sup>54</sup> | Multi-layer masks are better than single-layer masks. Masks should snugly fit the face with both nose and mouth covered. It may be necessary to extend masking requirements to places not currently mandated as of Dec 23, 2020 (e.g., workplaces, schools).<sup>54</sup> Masks (medical or non-medical) should continue to be worn in all public spaces and washed daily.<sup>57</sup> | If B.1.1.7 is associated with a higher viral load (speculated at the time of writing - Dec 23, 2020, later proven),<sup>9</sup> that would increase the amount of virus generated by respiratory activities, making physical distancing and ventilation very important. Reconsider 2m as the default distance (rather than 1m+) to reduce the risk of close-range transmission, however there is little benefit to increasing distancing beyond 2m.<sup>54</sup> Work from home if possible. If not possible, follow all physical distancing guidelines in the workplace. Avoid unnecessary travel, even within the province.<sup>57</sup> | At this time, there is no change to infection prevention and control measures recommended for COVID-19 based on the identification of a VOC as part of the outbreak. Health units should continue to follow setting-specific outbreak guidance.<sup>58</sup> Frequent, focused, cleaning of high hand-touch surfaces is likely to be more effective than cleaning surfaces where contact with hands is rare.<sup>54</sup> |
| *Personal services (e.g., hair salons, spas, tattoo parlors)* | For direct client services requiring gloves, it is important to properly remove gloves safely, and to wash hands thoroughly afterwards.<sup>56</sup> | It is recommended that service providers wear eye protection, such as a face shield or goggles, in addition to a non-medical mask when providing direct client services.<sup>56</sup> | NR* | NR |
| *Retail sector* | Handwashing supplies should be provided for workers and customers in the retail sector.<sup>55</sup> | Wear masks over nose and mouth at work, and eye protection - the latter especially when physical distancing cannot be maintained.<sup>55</sup> | Physical distancing remains the most effective way to reduce the risk of spreading COVID-19.<sup>55</sup> | Surfaces should be cleaned frequently, and retail areas should be well-ventilated.<sup>55</sup> |

\*NR: Not Reported

We identified 10 studies which contributed data relevant to modifying infection-control procedures which are summarized in Table 4. Of these 10 studies, 8 were modeling,^50, 59–62^ lab-based^63, 64^ or grey literature^65^ and thus not critically appraised. Of the two that were appraised, one was high quality,^27^ and the other was medium quality.^29^

**Table 4.**
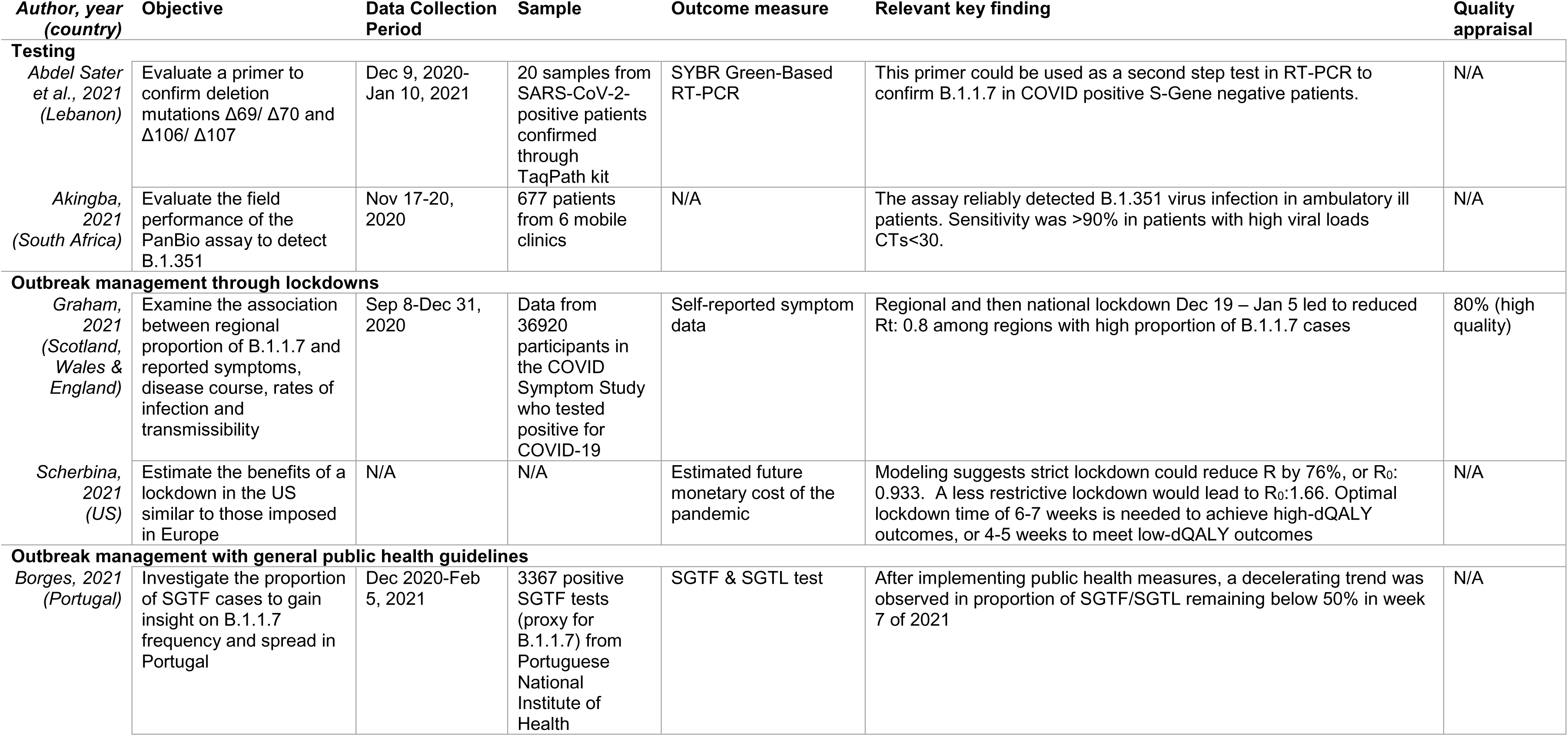
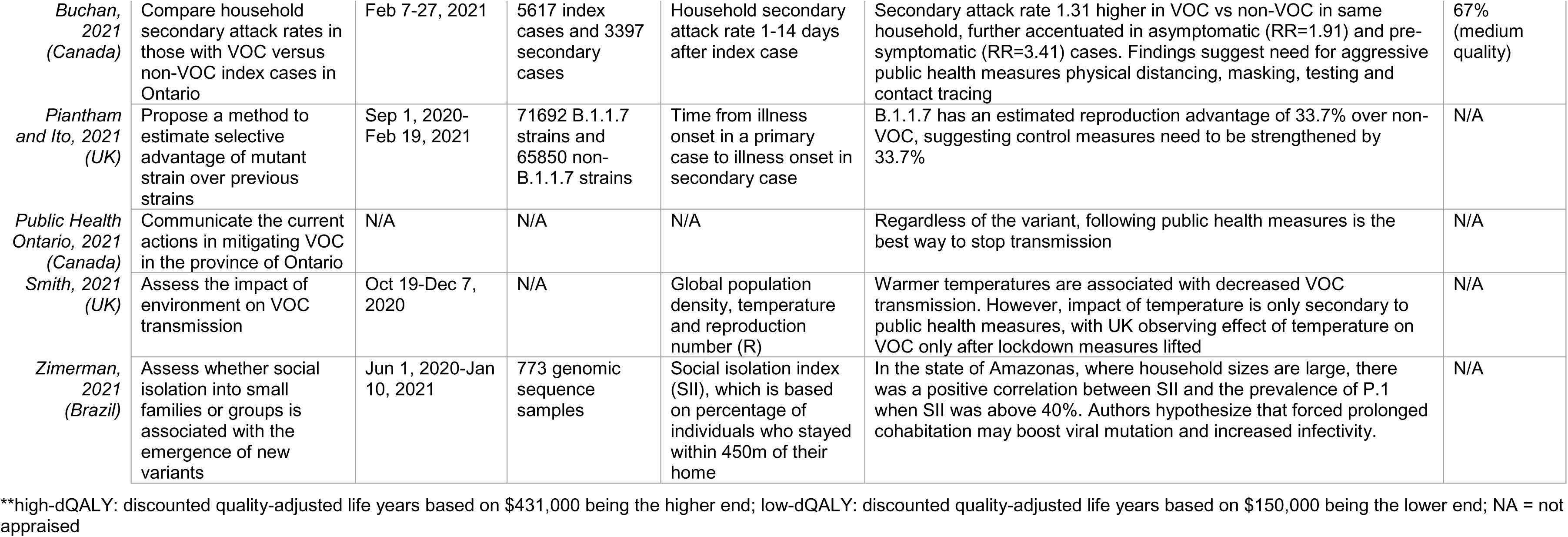
Summary of studies presenting findings on testing, public health measures, and infection-control procedures related to VOC

| Author, year<br>(country) | Objective | Data Collection<br>Period | Sample | Outcome measure | Relevant key finding | Quality<br>appraisal |
| --- | --- | --- | --- | --- | --- | --- |
| **Testing** |  |  |  |  |  |  |
| Abdel Sater<br>et al., 2021<br>(Lebanon) | Evaluate a primer to confirm deletion mutations $\Delta 69/ \Delta 70$ and $\Delta 106/ \Delta 107$ | Dec 9, 2020-Jan 10, 2021 | 20 samples from SARS-CoV-2-positive patients confirmed through TaqPath kit | SYBR Green-Based RT-PCR | This primer could be used as a second step test in RT-PCR to confirm B.1.1.7 in COVID positive S-Gene negative patients. | N/A |
| Akingba,<br>2021<br>(South Africa) | Evaluate the field performance of the PanBio assay to detect B.1.351 | Nov 17-20,<br>2020 | 677 patients from 6 mobile clinics | N/A | The assay reliably detected B.1.351 virus infection in ambulatory ill patients. Sensitivity was >90% in patients with high viral loads CTs<30. | N/A |
| **Outbreak management through lockdowns** |  |  |  |  |  |  |
| Graham,<br>2021<br>(Scotland,<br>Wales &<br>England) | Examine the association between regional proportion of B.1.1.7 and reported symptoms, disease course, rates of infection and transmissibility | Sep 8-Dec 31,<br>2020 | Data from 36920 participants in the COVID Symptom Study who tested positive for COVID-19 | Self-reported symptom data | Regional and then national lockdown Dec 19 – Jan 5 led to reduced Rt: 0.8 among regions with high proportion of B.1.1.7 cases | 80% (high quality) |
| Scherbina,<br>2021<br>(US) | Estimate the benefits of a lockdown in the US similar to those imposed in Europe | N/A | N/A | Estimated future monetary cost of the pandemic | Modeling suggests strict lockdown could reduce R by 76%, or $R_0$: 0.933. A less restrictive lockdown would lead to $R_0$:1.66. Optimal lockdown time of 6-7 weeks is needed to achieve high-dQALY outcomes, or 4-5 weeks to meet low-dQALY outcomes | N/A |
| **Outbreak management with general public health guidelines** |  |  |  |  |  |  |
| Borges, 2021<br>(Portugal) | Investigate the proportion of SGTF cases to gain insight on B.1.1.7 frequency and spread in Portugal | Dec 2020-Feb<br>5, 2021 | 3367 positive SGTF tests (proxy for B.1.1.7) from Portuguese National Institute of Health | SGTF & SGTL test | After implementing public health measures, a decelerating trend was observed in proportion of SGTF/SGTL remaining below 50% in week 7 of 2021 | N/A |

| **Author, year**<br>**(country)** | **Objective** | **Data Collection**<br>**Period** | **Sample** | **Outcome measure** | **Relevant key finding** | **Quality**<br>**appraisal** |
| --- | --- | --- | --- | --- | --- | --- |
| *Buchan, 2021*<br>*(Canada)* | Compare household secondary attack rates in those with VOC versus non-VOC index cases in Ontario | Feb 7-27, 2021 | 5617 index cases and 3397 secondary cases | Household secondary attack rate 1-14 days after index case | Secondary attack rate 1.31 higher in VOC vs non-VOC in same household, further accentuated in asymptomatic (RR=1.91) and pre-symptomatic (RR=3.41) cases. Findings suggest need for aggressive public health measures physical distancing, masking, testing and contact tracing | 67%<br>(medium quality) |
| *Piantham and Ito, 2021*<br>*(UK)* | Propose a method to estimate selective advantage of mutant strain over previous strains | Sep 1, 2020-Feb 19, 2021 | 71692 B.1.1.7 strains and 65850 non-B.1.1.7 strains | Time from illness onset in a primary case to illness onset in secondary case | B.1.1.7 has an estimated reproduction advantage of 33.7% over non-VOC, suggesting control measures need to be strengthened by 33.7% | N/A |
| *Public Health Ontario, 2021*<br>*(Canada)* | Communicate the current actions in mitigating VOC in the province of Ontario | N/A | N/A | N/A | Regardless of the variant, following public health measures is the best way to stop transmission | N/A |
| *Smith, 2021*<br>*(UK)* | Assess the impact of environment on VOC transmission | Oct 19-Dec 7, 2020 | N/A | Global population density, temperature and reproduction number (R) | Warmer temperatures are associated with decreased VOC transmission. However, impact of temperature is only secondary to public health measures, with UK observing effect of temperature on VOC only after lockdown measures lifted | N/A |
| *Zimmerman, 2021*<br>*(Brazil)* | Assess whether social isolation into small families or groups is associated with the emergence of new variants | Jun 1, 2020-Jan 10, 2021 | 773 genomic sequence samples | Social isolation index (SII), which is based on percentage of individuals who stayed within 450m of their home | In the state of Amazonas, where household sizes are large, there was a positive correlation between SII and the prevalence of P.1 when SII was above 40%. Authors hypothesize that forced prolonged cohabitation may boost viral mutation and increased infectivity. | N/A |

\*\*high-dQALY: discounted quality-adjusted life years based on \$431,000 being the higher end; low-dQALY: discounted quality-adjusted life years based on \$150,000 being the lower end; NA = not appraised

**Key findings for consideration include:**

- The U.S. Food and Drug Administration (FDA) issued a statement of concern on January 28, 2021 related to three tests: Accula SARS-CoV-2 test, TaqPath COVID-19 Combo Kit, Linea COVID-19 Assay Kit. Jurisdictions should ensure local testing is adequate to detect VOC
- Non-pharmaceutical interventions (e.g., lockdowns) appear to be important to limit transmission during vaccine rollout particularly if rapid mass vaccination is not feasible
- VOC transmission among individuals living in the same household, particularly among pre-symptomatic and asymptomatic cases, is concerning and warrants aggressive public measures (rapid testing, contact tracing, masking, quarantining and support for out of household quarantine)

We identified six guidance documents (low to high quality) related to quarantine/isolation (terminology often used interchangeably), testing, and contact tracing from five jurisdictions (Alberta,^66^ Ontario,^58^ Manitoba,^67^ UK,^68, 69^ and Ireland^70^) and one encompassing all of Canada. Guidance around these infection-control procedures varied across jurisdictions, but typically included a quarantine or isolation period of 10 to 14 days, mandatory testing at two points during quarantine/isolation, and enhanced contact tracing for suspected VOC cases.

Additionally, there were seven guidance documents related to outbreak management, including three that focused on recommendations for genomic surveillance methods, appropriate tests, and sampling approaches.^46, 47, 54, 58, 65, 71, 72^

###### Duration of quarantine and/or isolation

No studies were found relevant to the impact of VOC on duration of quarantine or isolation. However, according to a Centers for Disease Control and Prevention report, the increased transmissibility of VOC demonstrates the need for higher adherence to current mitigation strategies, including quarantine.^52^

###### Frequency or change of testing

We identified a wide range of studies evaluating different genome sequencing strategies, antigen tests or assays and primers for use in rapid PCR tests in our search. However, studies regarding testing were only included in this report if they explicitly identified implications for potentially modifying existing public health testing measures. Two studies and one report identified through the grey literature were deemed relevant for this sub-question.

Abdel Sater et al. designed and evaluated a primer set that could be used in a rapid, low-cost screening protocol to confirm deletion of mutations Δ69/ Δ70 in the spike gene and Δ106/ Δ107 in the NSP6 gene.^63^ The method was tested using 20 clinical samples from previously tested SARS-CoV-2 positive patients, 16 of which were S-negative and 4 were S-positive. The primer set successfully identified the presence and absence of S deletions Δ69/ Δ70 in 100% of both the S-negative and S-positive profiles. This protocol may be of particular benefit in areas where access to laboratories to conduct genome sequencing is limited.

Between November 17 and 20, 2020, Akingba et al. tested the field performance of the PanBio SARS-CoV-2 Rapid Antigen Test (RTD) with 677 patients attending one of 6 mobile clinics in Nelson Mandela Bay, South Africa.^64^ This rapid test is available and validated for use in Canada. At this time, South Africa was experiencing their second wave of the pandemic and B.1.351 was responsible for 84% of infections in Nelson Mandela Bay. The same nasopharyngeal swab used in the RTD was also sent for PCR for direct comparison. The antigen test had an overall sensitivity of 69.17% (95%CI:61.44,75.80) and specificity of 99.02% (95%CI:98.78,99.26). However, sensitivity improved in clinical samples with a high viral load (CT), with 100% detection when CT was <20, 95.5% when CT was 20-25 and 89.3% when CT was between 26-30.

On January 8, 2021, the FDA issued an emergency use authorization (EUA) statement containing caveats about the following three tests: 1) the Accula SARS-CoV-2 test performance might be impacted if the patient sample contains genetic variant at position 28881; 2) the TaqPath COVID-19 Combo Kit has significantly reduced sensitivity to certain mutations including B.1.1.7; and 3) the Linea COVID-19 Assay Kit has significant reduced sensitivity to certain mutations including B.1.1.7.^73^ Notably, the report did not include primary evidence to support these statements.

###### Contact tracing

No studies were identified related to impact of VOC on contact tracing.

###### Changing approach to outbreak management

Eight studies reported on outbreak management across a range of outcomes. Two studies discussed managing outbreaks by stricter NPIs. Graham et al. conducted an ecological study to explore the rate of infection and transmissibility of B.1.1.7 in the UK.^27^ Between September 28 and December 27 2020, B.1.1.7 was found to increase R_t_ (effective reproduction number) to 1.35 compared with historical SARS-CoV-2 variants. However, following a strict lockdown between December 19, 2020 and January 5, 2021 the estimated R_t_ of B.1.1.7 had decreased to 0.8 in three regions in England where 80% of infections were related to the variant. A modeling study conducted by Scherbina et al. identified the impact of different lockdown measures on community infection rates of B.1.1.7 with assessment of future monetary costs, in the form of missed work days, direct medical costs and the value of lost lives.^60^ The authors suggested that a strict lockdown could reduce the transmission rate to below one (R_0_=0.933), while a less strict lockdown would see the reproductive number exceed one (R_0_=1.66) and worsen the impact across all measures.

Five research articles and one grey literature source reported on managing outbreaks through the implementation of general public health measures. Borges et al. associated the implementation of public health measures, namely physical distancing, in Portugal with a decrease in proportion of SGTF/SGTL (i.e. an indicator of B.1.1.7).^50^

Another study by Buchan et al. conducted in Ontario, Canada compared the number of secondary attack rates among households with reported VOC cases versus households with non-VOC index cases.^29^ VOC secondary attack rates were 1.28 times higher for VOC versus non-VOC. Further, the secondary attack rates were observed to be higher for asymptomatic index cases (RR=1.91, 95%CI:0.96,3.80) and pre-symptomatic cases (RR=3.41, 95%CI:1.13,10.26). This significant increase transmission of VOC in households particularly for asymptomatic and pre-symptomatic cases suggests a need to support strict household quarantine guidance and provision of outside the home quarantine support and monitoring of cases.

Somewhat supporting that household transmission may be a key feature to monitor in VOC outbreaks, a study conducted by Zimerman et al. in Brazil, reported contrasting findings in terms of outbreak management.^62^ In their study comparing prevalence of the P.1 variant with social isolation data, authors found that prevalence of P.1 actually increased when individuals remained within 450m of their home. This study therefore highlights the need to tailor public health measures to specific populations and that vigilance regarding household transmission is warranted.

In a modeling study conducted by Piantham and Ito, it was estimated that B.1.1.7 has a reproductive advantage of 33.7% over non-VOC.^59^ Authors reported that public health measures should therefore be strengthened by 33.7% to account for the increased transmissibility of B.1.1.7.

Smith et al. assessed the impact of temperature on VOC prevalence in the UK.^61^ Warmer temperatures were found to be associated with lower VOC transmission, but this was only secondary to the impact of public health measures.

In addition to these four articles, a guidance document from Public Health Ontario broadly suggested that despite the VOC in circulation, following all existing public health measures would be the best way to stop transmission.^65^

###### Guidance documents related to quarantine/isolation, testing, and contact tracing

- Quarantine and/or isolation requirements vary across Canada and in other countries. Both 10-day (Alberta, Manitoba, UK) and 14-day (Ontario, Ireland) isolation periods are in effect in various jurisdictions; it is unclear whether these isolation periods were enacted in response to VOC, or if they pre-dated VOC. Testing is commonly required on day 0 and on or after day 10 of isolation (Alberta, Ontario, Manitoba, Ireland); UK requires testing on day 2 and day 8. Contact tracing approaches are variable but appear generally enhanced in response to VOC. In the UK and Ireland, more emphasis is placed on requiring a recent travel history from anyone presenting for care. The recommendations across jurisdictions are summarized in Table 5. **Please note that this table is not representative of all provinces (see note about guidance documents included in this review, page 4).**

###### Guidance documents related to outbreak management and prevention

- Engineering, procedural, and personal controls are essential for reducing transmission when interactions between people are unavoidable. This means people need to reassess their environments to ensure all precautions have been taken considering emerging VOC. Additionally, an indoor humidity of 40-60% is recommended.^54^
- Callan et al. defined Canada as a “Category G” country, in which COVID-19 was controlled or kept out until recently, but the future is uncertain as cases are currently increasing or peaking. In Category G countries, the authors recommend keeping all current disease control measures in place until vaccines become widely available and strengthened to compensate for new virus variants.^47^
- In Ireland, single VOC cases are managed as outbreaks by the Department of Public Health, triggering a full epidemiological investigation. This investigation focuses on whether the person has a history of travel in the preceding month; if not linked to travel, extensive efforts are required to identify any epidemiological links with other cases.^70^
- In Ontario, although no environmental cleaning protocols have been adapted in response to VOC, all VOC infections identified within the health system trigger immediate testing of all associated patients and staff, and sample sequencing of 1-3 cases to check for VOC is required with every COVID-19 outbreak. If a VOC outbreak is confirmed, all associated patients and staff should be tested every 3-5 days.^72^ When considering reopening or loosening of restrictions, Public Health Ontario recommends that “re-opening in a green-level public health unit (PHU) adjacent to a PHU with higher COVID-19 incidence may yield higher risks of COVID-19 than opting for a larger geographic region where all included PHUs have achieved similar COVID-19 control” and that, generally, gradual re-opening is more successful at controlling COVID-19 than rapid re-opening.^65^

###### Genomic surveillance guidelines

- The Canadian COVID Genomics Network (CanCOGen) recommends targeted genomic surveillance in the following scenarios, using multi-target COVID-19 RT-PCR tests with S-gene target dropouts (TaqPath three-target assay):^71^

○ All international travellers and their close contacts, prospectively
○ All international travellers and their close contacts, retrospectively
○ Cases of suspected reinfection
○ Severe acute COVID-19 cases in individuals younger than 50 years old without significant comorbidities
○ Cases in vaccinated individuals with subsequent laboratory-confirmed SARS-CoV-2 infection
○ Cases linked to known or suspected super spreading events
○ Geographic sampling in sub-regions with a pronounced increase in the case notification rate
○ Random sampling for routine national genomic surveillance
- Public Health Ontario is now using a VOC PCR test on eligible SARS-CoV-2 positive samples to detect both N501Y and E484K mutations at the same time; this will identify B.1.1.7, B.1.351, and P.1 and mitigates the need for whole genome sequencing (WGS) of all samples. WGS is best suited to population surveillance and not individual identification of VOC cases, as it is complex and takes 4-5 days.^75^
- The European Centres for Disease Control (ECDC) and WHO have co-produced a comprehensive guidance document outlining recommended approaches to screening and sequencing for VOC. The organizations recommend WGS to confirm VOC infection, and also support Sanger or partial next generation sequencing (NGS) amplicon-based sequencing, with which targeted whole or partial S-gene sequencing can be performed using a genetic analyzer. For the early detection and prevalence calculation of VOC, the organizations support several approaches to diagnostic screening assays, including S-gene drop out or target failure (SGTF), multiplex RT-PCR (including SGTF), screening SNP assays, screening SNP by specific real time RT-PCR melting curve analysis, and reverse transcription loop-mediated and transcription-mediated amplification isothermal amplification (RT-LAMP). Sequencing should be performed on at least a subset of these assays to confirm VOC identification. While the document acknowledges that rapid antigen detection tests appear to be as effective at identifying VOC as they are at previous variants, they do not differentiate between variants. The document includes several recommendations for VOC screening, including timely testing of people with symptoms, targeted or convenience sampling to aid early detection, and regular representative sampling to assess the level of VOC circulating in the community (and therefore reducing risk of bias in VOC screening results). The document ends with recommendations for assessing the quality of new testing methods or assays.^48^
- Ireland is developing a WGS program with plans to sequence up to 1500 samples per week. The samples will be collected both reactively (e.g., targeting travellers, clusters) and proactively (e.g., random representative sampling).^70^

#### Question 2: Health System Arrangements

Health system impacts due to VOC were reported in 17 original studies. The sections below are divided by sub-objectives and are discussed in relation to the relevant objective. Overall, 13 studies related to health system impacts were eligible for critical appraisal. Two were appraised as low quality,^38, 39^ seven as medium,^28, 30–35^ and four as high.^24–27^

**Table 5.**
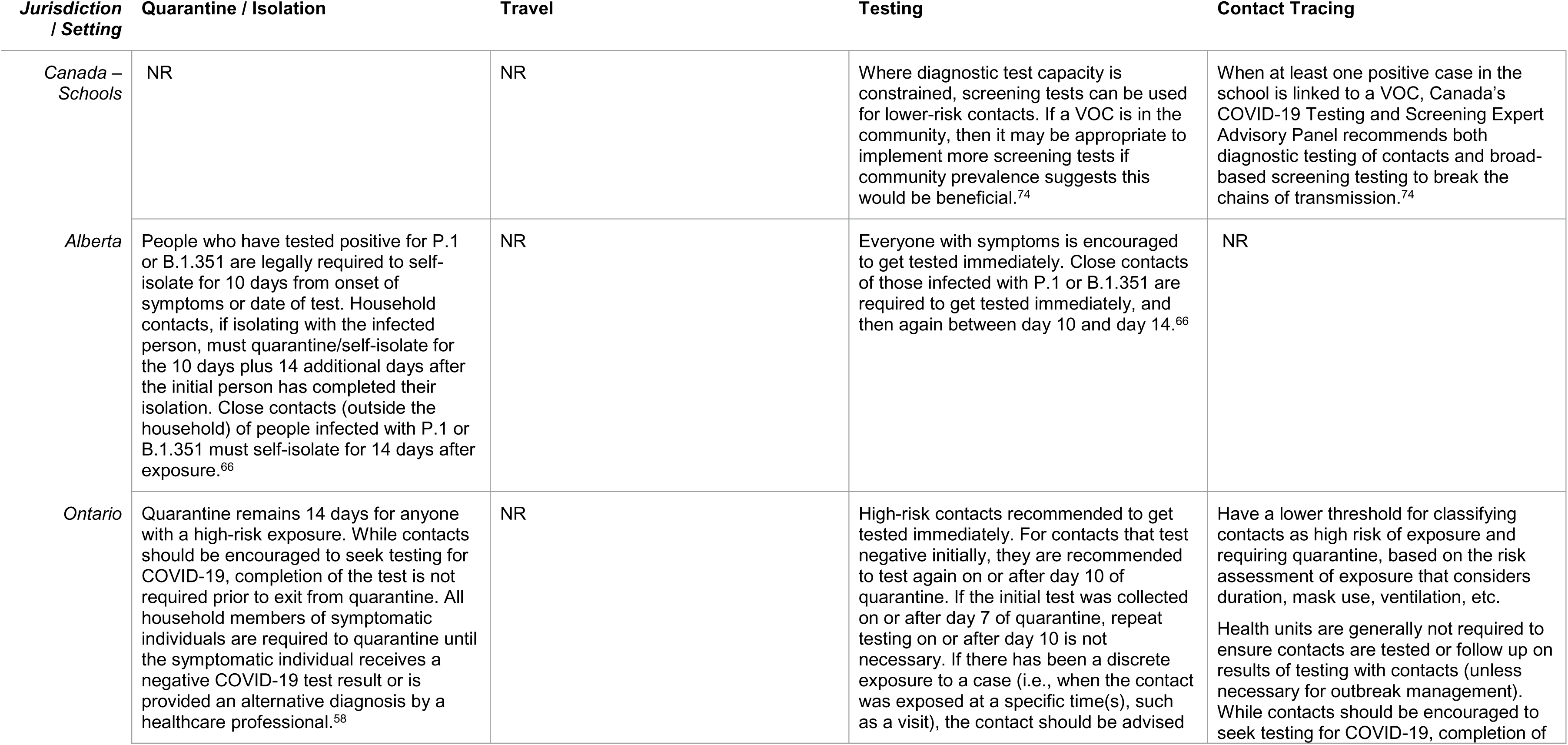
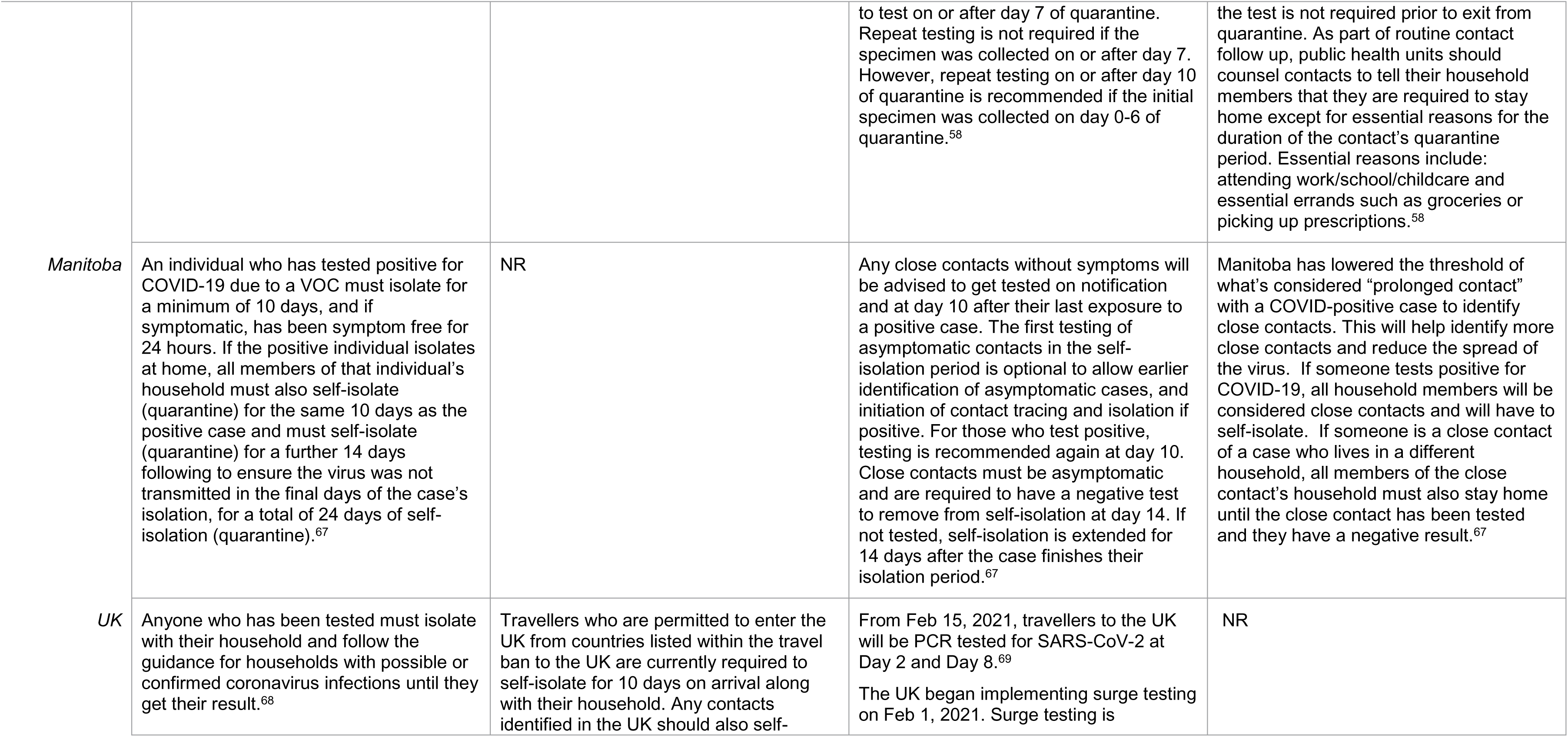
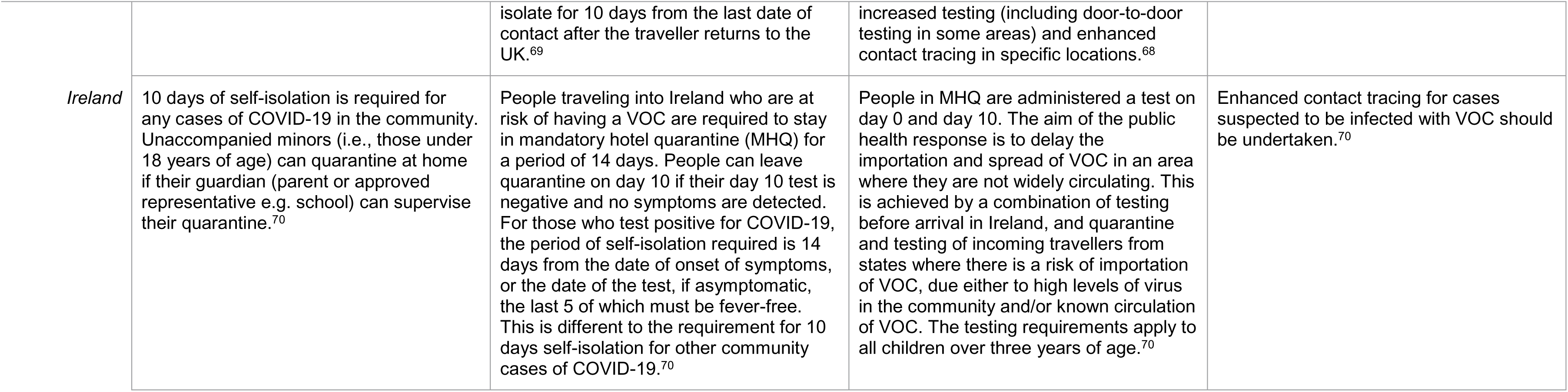
Guidance related to quarantine/isolation, testing, and contact tracing requirements in a selection of provinces and countries

##### Question 2A

Adjusting capacity planning to accommodate changes in the risk of re-infection and the risk of severe disease (e.g., hospitalization, admission to ICU, and death).

**Key findings for consideration include:**

- NPI (e.g., curfew, lockdown) may minimize risk of reaching hospital capacity due to VOC infections
- data is somewhat conflicting but overall suggests there is an increase in hospitalization due to B.1.1.7 but no difference in length of stay
- are mixed on the impact of B.1.1.7 on intensive care unit admissions
- While there are mixed findings on impact of VOC (B.1.1.7 and P.1) on death, there were more studies (n=6) that found an increased risk compared to no change (n=3). In studies that reported an increase in death, B.1.1.7 was found to increase the risk between 15% to 67% compared to non-VOC. The impact of rationed critical care and health system capacity strain on mortality may be difficult to separate in some of these studies
- guidance documents from Ontario refer to health facility capacity planning; neither refer specifically to increased hospitalization as a result of VOC, but rather communicate general guidance for moving COVID-19 patients between units

While most studies related to this sub-question reported on the impact of VOC on hospitalization, admission to ICU and death (see Table 6), Haas et al. also discussed impact of vaccine efficiency (VE).^32^ Haas et al. conducted the first nationwide estimates on the effectiveness of two doses of the Pfizer vaccine against a range of SARS-CoV-2 outcomes, including hospitalization and deaths, in Israel. Haas was appraised as medium quality in the critical appraisal. Between December 2020 and March 2021, there were 202,684 COVID-19 infections, 139,835 (69.0%) in people over 15 years. There were 6,040 hospitalizations, of which 3,470 were severe and critical, and 754 deaths in people over 15 years. The prevalence of B.1.1.7 of tested cases was 93.9%. During the study period, over half (51.5%) of people over 15 years and 82.8% of people over 65 years received two doses of Pfizer. Among COVID-19 related hospitalizations, most were in people unvaccinated (4,382, 72.5%) with a small number of admissions who had received two doses in the prior 7 days (421, 7.0%). The incidence rate (IR) (per 100 000 person-days) of COVID-19 among people over 15 years was 116.2 in unvaccinated and 5.3 in vaccinated at least 7 days after the second dose. The adjusted VE was 94.1% (95% CI 93.4–94.7) against COVID-19 infection. Adjusted VE against COVID-19 hospitalization was 96.0% (95% CI 95.2–96.6) and VE against severe and critical hospitalization was 96.2% (95% CI 95.5–96.8). In relation to deaths, 457 (60.6%) were in unvaccinated individuals and 99 (13.1%) in individuals who had received the second dose at least 7 days ago. Adjusted VE against death was 93.3% (95% CI 91.5–94.8). VE estimates against all outcomes were slightly higher when measured 14 days after the second dose than VE estimates that were done after 7 days; however, overall, this study suggested there is high VE 7 days after the second doses of Pfizer against hospitalizations, severe and critical hospitalizations, and deaths.

**Table 6.**
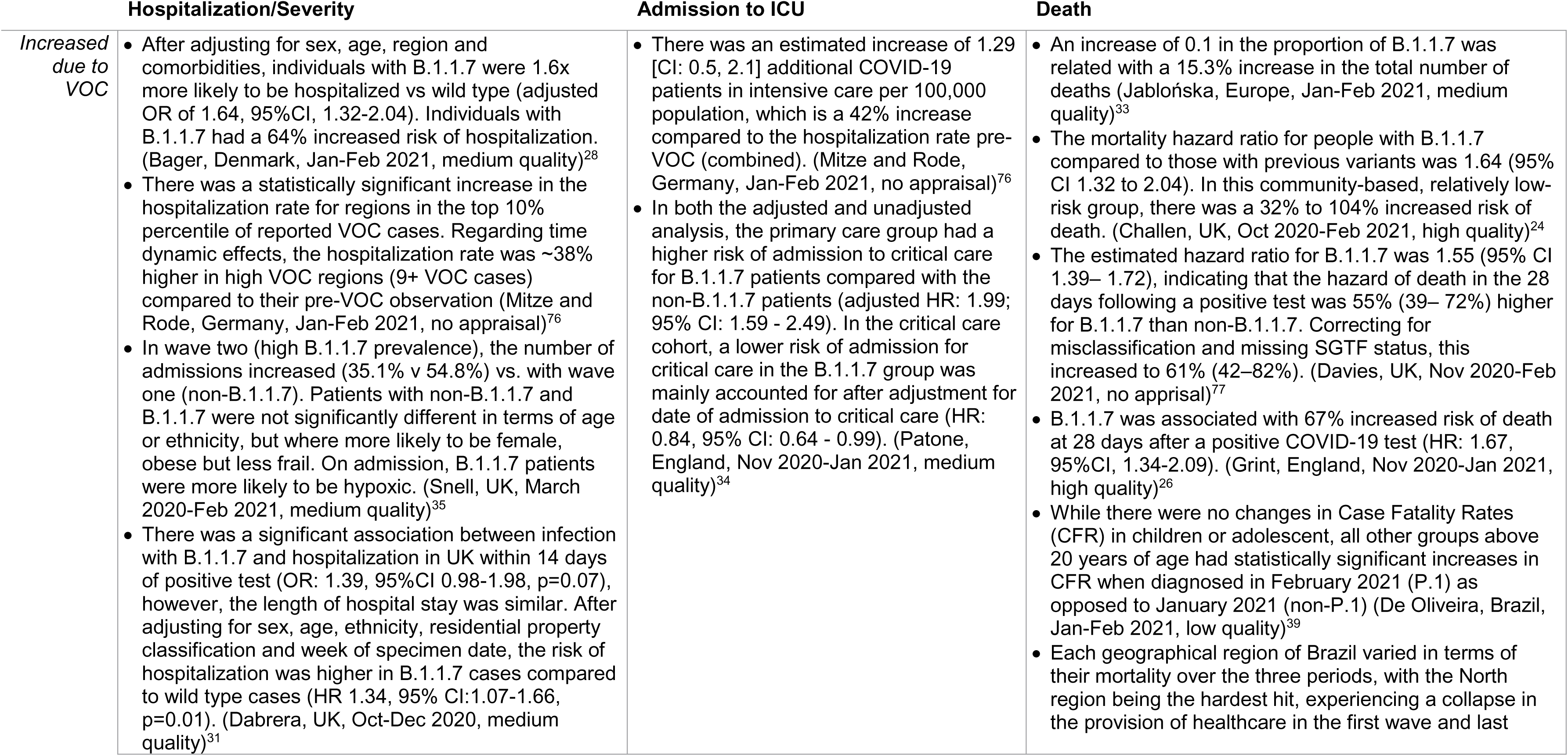
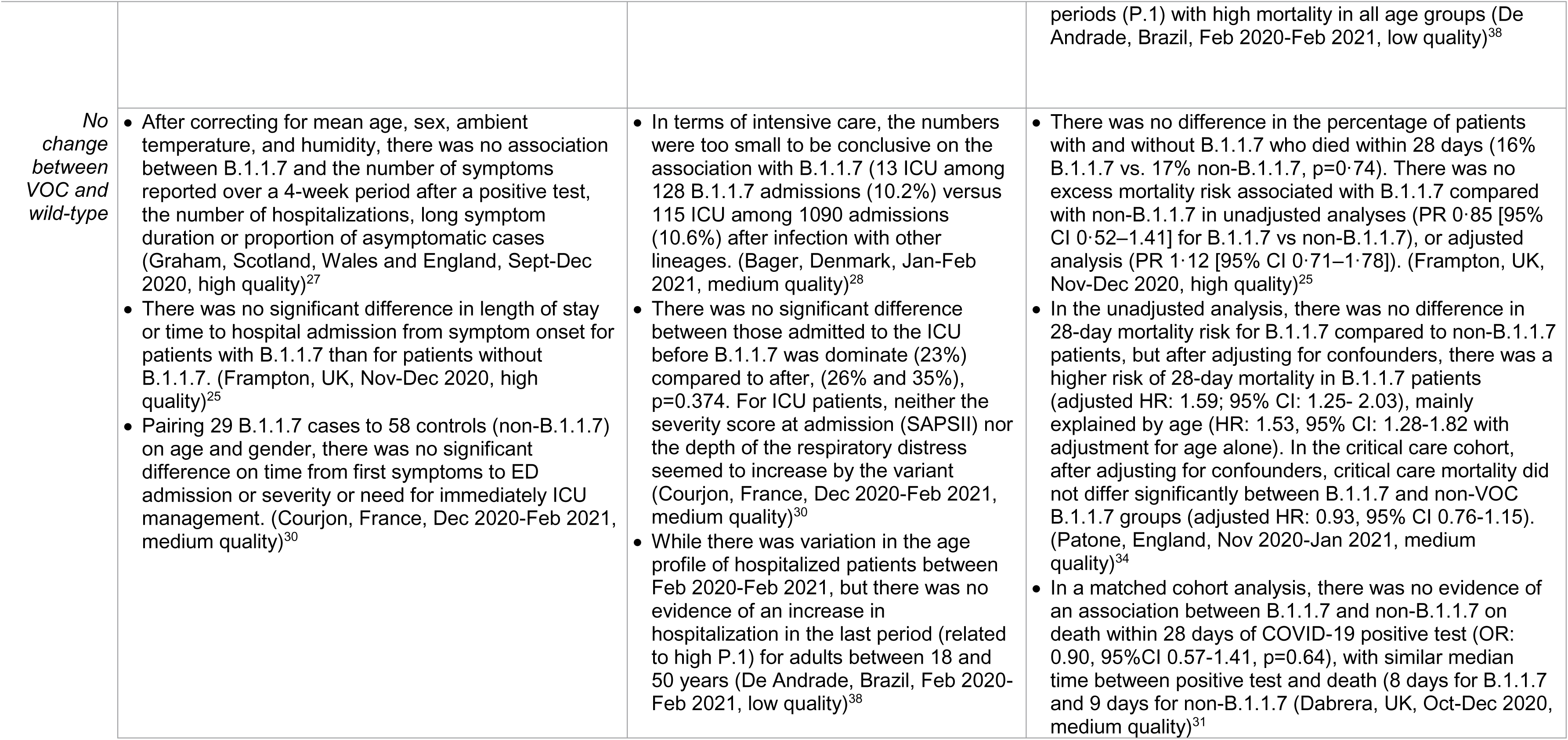
Summary of findings for capacity planning and health systems arrangements

Domenico et al. provided a unique mathematical model to estimate the role that curfew measures could have on hospitalization in France.^40^ They found that if the epidemic progressed under curfew conditions (6pm nightly, implemented nationwide January 16^th^) before school holidays and vaccination is accelerated, hospital capacity would be reached around week 13 in France (which had 2.2% B.1.1.7 penetration), week 12 in Île-de-France (which had the highest B.1.1.7 penetration, 6.9%), and week 14 in Nouvelle Aquitaine (which had the lowest B.1.1.7 penetration 1.7%). The partial relaxation of social distancing (estimated at 15% increase in effective reproduction number) would shorten these estimates by at least 1 week. Stronger social distancing, equivalent to the efficacy measured during the second lockdown (estimated 15% reduction in effective reproduction number), would maintain hospitalizations below the peak of the second wave in Île-de-France and Nouvelle Aquitaine but would not be enough to avoid a third wave in France, even under accelerated vaccination (100k-200k doses/day). Accelerated (200,000 first doses/day) and optimistic vaccination rollouts (300,000 first doses/day) would reduce weekly hospitalizations by about 20% and 35% in week 16 (i.e., April 19-25, 2021) compared to a stable vaccination campaign without acceleration (100,000 first doses/day).

###### Hospitalization/Severity

Seven studies reported on health system impacts related to hospitalization and severity of illness. Four studies in Europe reported increases in hospitalization and/or severity of illness associated with B.1.1.7 compared to the wild type; however, another three studies found no difference. Among the seven studies that reported on hospitalization, four were appraisal as medium quality studies and two as high-quality studies (one not appropriate for appraisal). Table 6 below provides a summary of findings with additional detail provided in text below.

**<u>Increases in risk of hospitalization/severity</u>**

Bager et al. conducted an observational cohort study of 35,887 SARS-CoV-2 positive individuals in Denmark between January 1^st^ to February 9^th^, 2021.^28^ While Denmark was in a lockdown after December 16^th^ and it was effective in reducing case numbers and hospital burden, B.1.1.7 increased during this time and was found to have a reproduction number of 1.25 on February 16^th^. 11.6% of their sample had B.1.1.7 and the proportion of individuals with B.1.1.7 increased from 1.9% to 45.1% during their data collection period. They found no significant difference in hospitalization between B.1.1.7 vs non-B.1.1.7 in their crude analysis (OR 0.87; 95%CI, 0.72-1.05), but after adjusting for sex, age, region and comorbidities, B.1.1.7 was 1.6 times more likely to be associated with hospitalized than wild type (adjusted OR of 1.64, 95%CI, 1.32-2.04). They concluded that individuals infected with B.1.1.7 had a 64% increased risk of hospitalizationcompared to individuals infected with other lineages.

An epidemiological and modeling study was conducted in Germany on the 7-day incidence rate (per 100,000 population), and the hospitalization rate (per 100,000 population).^76^ All three VOC were combined for power due to the low spread in Germany at the time of the study – by February 4^th^, 2021, 204 out of 401 (50.1%) of the HUT-3 regions had at least one VOC case. Hard-hit cities included Flensburg where the 7-day incidence rate drastically increased in January 2021 relative to the average development in Germany due to illegal parties and NRW cities (Cologne, Leverkusen and Duren). The hospitalization rate for Flensburg shows a significant increase ∼16 days after treatment start (considered one week before the first VOC case). However, this should be interpreted with caution as it was calculated on a relatively small number of hospitalized patients per 100,000 population at the local area level (i.e., 2 patients before VOC vs. 8 cases after VOC). Interestingly, treatment effects estimate, for the NRW cluster, an increase in the 7-day incidence rate by ∼40% but no significant increase in hospitalization rates. Difference-in-difference analysis point to positive but insignificant correlations for all regions but in regions with an early VOC reporting before January 22^nd^, they found a statistically significant increase in the hospitalization rate for regions in the top 10% percentile of reported VOC cases. Regarding time dynamic effects for the hospitalization rate, significant effects were found ∼15 days after treatment start for regions with 9+ VOC cases. After 20 days, the hospitalization rate was ∼38% higher in high VOC regions compared to their pre-VOC observation. This corresponds with two additional patients in intensive care per 100,000 in high VOC regions, suggesting that hospitalization tends to grow over time with B.1.1.7 cases.

Snell et al. conducted a retrospective cohort study in the United Kingdom on 2,341 individuals admitted to the Guy’s Hospital and St. Thomas Hospital within 14 days following a positive test during first wave (March 13, 2020-May 12, 2020, n=838, considered non-B1.1.17) and second wave (October 2020-Feb 2021, n=1503, predominantly B.1.1.7).^35^ In wave two, the number of admissions increased compared with wave one (54.8% vs. 35.1%). In the second wave, B.1.1.7 made up 83% of all sequenced isolates and 85% of sequenced isolates from admitted cases. Snell et al. examined differences between the two waves in terms of patient composition. While the general statistics were similar, they found that wave two hospitalized patients (regardless of VOC status) were slightly younger (60 vs. 62 years, p=0.019) and more likely to be female (47.3% vs. 41.8%, p=0.011). Considering comorbidities, individuals in wave two were less likely to have a diagnosis of frailty (11.5% v 22.8%, p<0.001), have a history of stroke (4.3% v 8.6%, p<0.001) or cancer (4.8% v 7.2%, p=0.022) but were more likely to be obese (29.1% v 24.6%, p=0.02). There was no significant difference on other comorbidities of diabetes, kidney disease, hypertension, cardiovascular disease, or respiratory disease. Shifting to comparing admitted patients with B.1.1.7 (n=400) or non-B.11.7 VOC (n=910), the groups were not significantly different in age (62 years vs 64 years, p=0.22) or ethnicity. However, patients with B.1.1.7 were more likely to be female (48.0% vs 41.8%, p=0.01), less likely to be frail (14.5% vs 22.4% p=0.001), more likely to be obese (30.2% v 24.8%, p=0.048), and more likely to be hypoxic on admission (70.0% vs 62.5%, p=0.029), the main indicator of severe disease, than non-B.1.1.7 patients.

Dabrera et al. conducted a matched cohort study in the UK to explore whether B.1.1.7 was associated with more severe clinical outcomes compared to wild type COVID-19.^31^ In 5,642 cases, 131 individuals had a hospital admission within 14 days of their positive test: 76 (2.7%) VOC and 55 (1.9%) wild-type cases (p=0.006). There was a significant association between infection with B.1.1.7 and hospitalization within 14 days of positive test (OR: 1.39, 95%CI 0.98-1.98, p=0.07); however, the length of hospital stay was comparable (B.1.1.7 median length of stay (LOS) 5 days (IQR 3-10, range 0-37) vs wild type LOS 8 days (IQR 4-13.5 days, range 0-31), p=0.07). In univariable analysis, B.1.1.7 infection and risk of hospitalization within 14 days were not associated (HR: 1.07, 95%CI: 0.89-1.29, p=0.48); however, adjusting for potential confounders (sex, age, ethnicity, residential property classification and week of specimen date) resulted in the risk of hospitalization being higher for B.1.1.7 cases compared to wild type cases (HR 1.34, 95% CI:1.07-1.66, p=0.01).

**<u>No changes in risk to hospitalization/severity</u>**

Graham et al. conducted a longitudinal cohort study on 36,920 users of the COVID symptom study mobile app in Scotland, Wales and seven regions in England who tested positive for COVID-19 between September 28^th^ and December 27^th^, 2020 while the proportion of B.1.1.7 was exploding, along with surveillance data from 98,170 sequences from the COVID-19 UK Genetics Consortium (COG-UK), 16% of which were B.1.1.7.^27^ Therefore this study took place before health system strain was excessive. They compared the proportion of B.1.1.7 in each region and the proportion of reports per week for each symptom, disease burden and self-reported hospitalization. After adjusting for mean age, sex, ambient temperature, and humidity, there was no association between B.1.1.7 and the number of symptoms reported over a 4-week period after a positive test, the number of hospitalizations, long symptom duration or proportion of asymptomatic cases.

Frampton et al. conducted a cohort study in the UK to describe emergence of B.1.1.7 in two North Central London hospitals including clinical outcomes in patients with and without the VOC.^25^ Of the 341 samples sequenced and positive for SARS-CoV-2, 198 (58%) were B.1.1.7 and 143 (42%) were non-B.1.1.7. Eighty-eight (44%) of B.1.1.7 patients received oxygen by mask or nasal prongs compared with 42 (30%) of non-B.1.1.7 patients. While length of stay, risk of hospitalization within 14 days of a test, and time to hospital admission from symptom onset were similar, B.1.1.7 patients were younger, had fewer comorbidities and more likely to be from an ethnic minority compared to non-B.1.1.7 patients.

Courjon et al. conducted an observational study in France (Nice) with 1,247 patients admitted to the emergency department to analyze changes in clinical profile and outcomes.^30^ Of those admitted by February 22^nd^, 29 had B.1.1.7 (12.5%). This reflects the prevalence of B.1.1.7 in the area, which increased from 2.6% in December 2020 to 79.1% in February. In hospitalized patients, the mean age of admission was significantly lower in the period between Feb 8^th^-22^nd^ (considered high B.1.1.7) at 59.2 years (SD=14.0) compared to December 7^th^-21^st^ (no/low B.1.1.7) at 70.7 years (SD=13.6), p < 0.001. Patients were also more likely to have no comorbidity (42% vs. 16%, p =0.04). When pairing 29 cases to 58 controls on age and gender, there was no significant difference between B.1.1.7 patients and non-B.1.1.7 on time from first symptoms to emergency department admission or severity or need for immediate ICU management.

###### Admission to ICU

Five studies reported on health system impacts related to admission to ICU. Two studies in Europe reported increases in admission to ICU with B.1.1.7 compared to the wild type; however, another three studies found no difference. Among the five studies that reported on admission to ICU, three were apprised as medium quality and one as low quality (one not appropriate for appraisal). Table 6 provides a summary of findings with additional detail provided in text below.

**<u>Increases in ICU Admission</u>**

Mitze and Rode conducted an epidemiological and modeling study in Germany on the hospitalization rate per 100,000 population.^76^ All three VOC were combined for power due to the low spread in Germany at the time of the study. Mitze and Rode found that an increase of 1.29 [CI: 0.5, 2.1, p<0.05] additional COVID-19 patients in intensive care per 100,000 population, which is a 42% rise in hospitalization in VOC regions compared to pre-VOC regions (3.08 patients in intensive care per 100,000 population).

Patone et al. conducted a retrospective cohort study in England to explore the risk of critical care admission for patients with B.1.1.7 compared with wild type.^34^ They compared two cohorts: a ‘primary care cohort’ which was patients in primary care with a positive community COVID-19 test reported between November 1^st^, 2020 and January 26^th^, 2021 (n=381,887, 52.0% B.1.1.7, and 712 were admitted to critical care, 63.1% B.1.1.7). The ‘critical care cohort’ were patients admitted for critical care with a positive community COVID-19 test reported with an identified SGTF status between November 1^st^, 2020 and January 27^th^, 2021 (n=3432, 58.8% B.1.1.7). In both the adjusted and unadjusted analysis, the primary care group had a higher risk of admission to critical care for B.1.1.7 patients compared with the non-B.1.1.7 patients (adjusted HR: 1.99; 95% CI: 1.59 - 2.49). Considering time varying HR, it increased from 1.20 (95% CI: 0.58 - 2.48) at one day to 3.29 (95% CI: 1.17 - 6.29) at fifteen days after a positive test. There was no significant interaction between B.1.1.7 and sex, ethnic group, or age group. When adjusting only for the date of positive test, it did not account for the increased risk of admission for critical care (adjusted HR 1.28 95% CI: 1.05 - 1.56). In the critical care cohort, B.1.1.7 ICU patients tended to be younger (means 57.8 versus 59.3 years) and less obese than non-B.1.1.7 patients. Acute severity of illness, as measured by the APACHE II score, tended to be lower in B.1.1.7 patients, but the proportion receiving invasive mechanical ventilation within the first 24 hours of critical care and organ support was similar between the two groups. After adjusting for date of admission to critical care, the lower risk of admission for critical care in the B.1.1.7 group was accounted for (adjusted HR: 0.84, 95% CI: 0.64 - 0.99).

**<u>No Changes in ICU Admission</u>**

Bager et al. conducted an observational cohort study in 35,887 SARS-CoV-2 positive individuals in Denmark between January 1^st^ to February 9^th^, 2021.^28^ While Bager et al. was able to find an increased risk of general hospitalization due to B.1.1.7, the numbers were inconclusive on the impact of B.1.1.7 on intensive care admission due to small sample sizes. They reported only 13/128 ICU among B.1.1.7 admissions (10.2%) compared to 115/1090 ICU admissions after infection with wild type (10.6%).

Courjon et al. conducted an observational study in France (Nice) with 232 patients hospitalized in the infectious disease ward and ICU to analyze modification in clinical profile and outcome traits.^30^ They found no significant difference between percentage of COVID-19 hospitalized patients admitted to the ICU before B.1.1.7 was dominate (23% [December 7-21, 2021]) compared to after (26% [January 24-February 7, 2021] and 35% [February 8-22, 2021]), p=0.374. For ICU patients, neither the severity score at admission (SAPSII) nor the depth of the respiratory distress seemed to increase with B.1.1.7.

De Andrade et al. conducted an epidemiological study during the first year of the pandemic to compare the age profile of patients hospitalized with COVID-19 as well as hospital mortality and ICU use by age group in large geographic regions of Brazil.^38^ They compared across three time periods: (1) February 16^th^- June 20^th^ 2020; (2)June 21- October 24^th^ 2020; (3) October 25^th^, 2020 and February 20^th^, 2021. The third timepoint corresponds with the increasing prevalence of P.1 in Brazil. Of the 620,363 completed records of patients hospitalized, 244,611 (34.0%) had indication for use of ICU. While there was variation in the age profile of hospitalized patients between the three periods, there was no evidence of an increase in hospitalization the last period for adults between 18 and 50 years. Geographically, they report that in the North and Northeast regions, the proportion of 18–50-year-olds in the last period was similar to the first period during which they were also substantially affected by the pandemic. In the Southeast and South, there was a consistent reduction in these age groups over the periods. In the Central West region, which experienced the pandemic more during the second period, hospitalization with adults between 18 and 50 years old was higher in the first period, decreasing and maintain during the second and third period. They conclude that the rise in the North aligns with a collapse of the health care system in some areas, which was likely associated with disease severity due to P.1.

###### Death

Nine studies reported on health system impacts related to risk of death. Four studies in Europe and two in Brazil reported increased risk of death for individuals with B.1.1.7 compared to the wild type; however, another three studies found no difference. Among the nine studies, three were apprised as high quality, three were medium quality, and two were low quality (one not appropriate for appraisal). Table 6 provides a summary of findings with additional detail provided in text below.

**<u>Increases in Death</u>**

Jabłońska et al. conducted a correlational study across 38 European countries to detect the association between COVID-19 mortality and proportion of VOC through the second wave in Europe using multivariate regression models.^33^ A higher proportion of B.1.1.7 across countries was associated with higher COVID-19 mortality peak and total mortality during the second wave of the pandemic in Europe. Between January 1^st^ to February 25^th^, 2021, “an increase of 0.1 in the proportion of B.1.1.7 was related to a 15.3% increase in the cumulative number of deaths during that period” (p.5).

Challen et al. conducted a matched cohort study in the UK to explore whether there is change in mortality at 28 days from infection with B.1.1.7 compared with wild type.^24^ The study consisted of 54,906 matched pairs who tested positive for COVID-19 between October 1, 2020 and January 29, 2021 (followed until February 12, 2021). In 54,902 matched cohort pairs, there were 227 deaths in B.1.1.7 individuals vs. 141 non-B.1.1.7 individuals. The mortality hazard ratio for people with B.1.1.7 compared to those with non-B.1.1.7 was 1.64 (95% CI 1.32 to 2.04). As a community-based study, this suggests that even in a relatively low-risk group, there was an increased risk of death from 32% to 104%.

Davies et al. conducted an epidemiological modeling study to describe the association between B.1.1.7 and hazard of death and disease severity within 28 days of positive test in the UK.^77^ Of the 2,245,263 individuals with a positive community test between November 1, 2020 and February 2021, half (51.1%) had a conclusive SGTF reading and, of these, 58.8% had SGTF, indicative of B.1.1.7. Among those with known SGTF status, the crude COVID-19 death rate was 1.86 deaths per 10,000 person-days of follow-up in the B.1.1.7 group versus 1.42 deaths in the non-B.1.1.7 group. The hazard ratio for B.1.1.7 was 1.55 (95% CI 1.39– 1.72), meaning that the risk of mortality in the 28 days following a positive test was 55% higher for B.1.1.7 than for non-B.1.1.7 cases. After correcting for misclassification of SGTF and missing SGTF status, there was a 61% (95% CI: 42–82%) higher hazard of death associated with B.1.1.7; however, this was not consistent across age groups. In females aged 70–84, the estimated risk of death within 28 days of a positive SARS-CoV-2 test increased from 2.9% without B.1.1.7 to 4.4% with B.1.1.7 (95% CI 4.0–4.9%) and for males 70-84, it increased from 4.7% to 7.2% (95% CI: 6.4–7.9%). Similarly, in females 85 or older, the estimated risk increased from 13% to 19% (95% CI: 17–21%) and for males 85 or older, it increased from 17% to 25% (95% CI: 23-27%). These estimates reflect a substantial increase in absolute risk in older age groups, but the risk of COVID-19 death following a positive test in the community remains below 1% in most individuals younger than 70 years old.

Grint et al. conducted a cohort study to estimate the risk of death following COVID-19 infection in England by comparing B.1.1.7 to non-B.1.1.7.^26^ They found that B.1.1.7 was consistently associated with increased risk of death within 28 days compared to non-B.1.1.7 cases with the hazard ratio at 1.67 (95%CI, 1.34-2.09, p<0.0001). The risk of death was low for those under 65 years of age without comorbidities, though higher for males than females (B.1.1.7: Males 0.14%; Females: 0.07% vs. non-B.1.1.7: Males: 0.09%; Females: 0.05%). The risk of death was consistently higher for males and increased with age and with comorbidities. The highest risk was seen among those aged 85 years or older with 2+ comorbidities (B.1.1.7: Males 24.3%; Females: 14.7% vs. non-B.1.1.7: Males: 16.7%; Females: 9.7%).

De Oliveira et al. conducted an epidemiology study to explore data from the state of Parana, in the south of Brazil, where the P.1 variant was identified on February 16^th^, 2021, to assess trends in mortality data as reported CFRs among different age-groups.^39^ Prior to the introduction of P.1, all age groups had either a decline or stable CFR, however, in February 2021, an increase in CFR for almost all age groups was observed. While there were no changes in CFR in children or adolescent, all other groups above 20 years of age had statistically significant increases in CFR when diagnosed in February 2021 as opposed to January 2021. For individuals between 20 and 29 years of age, there was a 3-fold higher risk of death when diagnosed in February 2021 compared to January (RR: 3.15 [95%CI: 1.52-6.53], p<0.01). This risk of death was also higher in other age groups, although to a lesser extent – 93% for 30–39- year-old (1.93 [95%CI:1.31-2.85], p<0.01), 11-% for 40-49-year-old (RR: 2.10 [95%CI:1.62-2.72], p<0.01), and 80% for 50-59-year-old (RR: 1.80 [95%CI:1.50-2.16], p<0.01).

De Andrade et al. conducted an epidemiological study during the first year of the pandemic to compare the age profile of patients hospitalized by COVID-19 as well as hospital mortality and use of ICUs by age group in large geographic regions of Brazil.^38^ They compared across three time periods: (1) February 16^th^ and June 20^th^ 2020; (2) June 21^st^ and October 24^th^ 2020; (3) October 25^th^, 2020 and February 20^th^, 2021. The third timepoint corresponds with the increasing prevalence of P.1 in Brazil. Each region varied in terms of their mortality over the three periods, with the North region being the hardest hit, experiencing a collapse in the provision of healthcare in the first and last periods with high mortality in all age groups, with a rise in hospital deaths among adults aged 18-60 years. The high mortality in the third wave was among adults aged 18-70 years, reflecting the severity of the pandemic in the region and the impact of P.1.

**<u>No Changes in Death</u>**

Frampton et al. conducted a cohort study in the UK to describe emergence of B.1.1.7 in two North Central London hospitals including clinical outcomes in patients with and without the VOC.^25^ Of the included 399 patients, the proportion of patients who had severedisease (i.e., WHO level of ≥6) or death were similar: 28% in the non-B.1.1.7 group vs. 36% in the B.1.1.7. The proportion of patients at level 6 or levels 7–9 on the WHO ordinal scale or who died were not statistically different: 18% in the non-B.1.1.7 group were at level 6 and 2% were at levels 7–9; 15% in the B.1.1.7 group were at level 6 and 6% were at levels 7–9. Similar rates of mortality were found, with 16% patients with B.1.1.7 dying within 28 days versus 17% with non-B.1.1.7. In both the unadjusted and adjusted analysis (controlling for hospital, sex, age, comorbidities, and ethnicity), there was no increased risk of mortality or severe disease with B.1.1.7 compared to non-B.1.1.7.

Patone et al. conducted a retrospective cohort study in England to explore the risk of critical care admission for patients with B.1.1.7 compared with the wild type.^34^ They compared two cohorts: a ‘primary care cohort’ and ‘critical care cohort’ described above. There was no difference in death within 28 days between the B.1.1.7 group and non-B.1.1.7 group (0.3% both). In the unadjusted analysis, there was no difference in 28-day mortality risk for B.1.1.7 compared to non-B.1.1.7 patients, but after adjusting for confounders, there was a higher risk of 28-day mortality in B.1.1.7 patients, (adjusted HR: 1.59; 95% CI: 1.25- 2.03), mainly explained by age (HR: 1.53, 95% CI: 1.28-1.82 with adjustment for age alone). In the critical care cohort, after adjusting for confounders, critical care mortality did not differ significantly between B.1.1.7 and non-VOC B.1.1.7 groups (adjusted HR: 0.93, 95% CI 0.76-1.15). Both cohorts had no evidence of an interaction between B.1.1.7 and ethnic group, age group, or sex.

Dabrera et al. conducted a matched cohort study in the UK to explore whether B.1.1.7 was associated with more severe clinical outcomes compared to wild type.^31^ In the matched cohort study of 5,642 individuals, 76 died within 28 days of a positive test; of which, 36 (1.3%) were infected with B.1.1.7 and 40 (1.4%) were infected with wild-type SARS-CoV-2. There was no association between B.1.1.7 and non-B.1.1.7 groups and death within 28 days of COVID-19 positive test (OR: 0.90, 95%CI 0.57-1.41, p=0.64), with similar median time between positive test and death (8 days for B.1.1.7 and 9 days for non-B.1.1.7 (Kruskal Wallis p=0.79). In the unadjusted analysis, there was a negative relationship between risk of death and B.1.1.7 infection (HR: 0.54, 95%CI:0.42-0.69, p=0.00); however, after adjusting for confounders (sex, age, ethnicity, residential property classification, week of specimen date and testing Pillar), there was no difference in risk of death among B.1.1.7 cases compared to non-B.1.1.7 (HR: 1.06, 95%CI:0.82-1.38, p=0.65).

##### Question 2B

Adjusting personal protective equipment (PPE) procedures for health workers

One modeling study, which was not critically appraised, reported on this outcome. Pham et al. evaluated the impact of different interventions on transmission, HCW absenteeism and test positivity as a marker of intervention efficiency against B.1.1.7 transmission through modeling.^78^ In the baseline scenario, it was assumed that HCWs were using PPE while in COVID wards when seeing patients but not during breaks or when in other parts of the hospital, assuming 95% of HCW worked in same wards over time. While specific PPE was not defined, PPE efficiency was defined as percentage reduction of droplet transfer. Assuming 90% effective PPE use in COVID wards, they found that extending PPE use to non-COVID wards (all HCW used PPE with 90% effectiveness when on ward) would prevent 93.7% of all transmissions and would also prevent outbreaks among patients and HCWs. Even if PPE effectiveness was reduced to 70%, findings did not change significantly, however, if was reduced to 50% or below, screening HCW every 3 days was more effective than PPE use in all wards. Overall, PPE use in all wards was model to be more effective than all other interventions.

###### Guidance documents related to PPE procedures

- Four guidance documents (one low-quality,^79^ two medium-quality,^69, 70^ and one high-quality^75^) located from the UK, Ontario, and Saskatchewan report on PPE procedures. If stringent procedures were already in place prior to the emergence of VOC, PPE procedures in healthcare settings remain relatively unchanged in response to VOC.^68, 69^ Saskatchewan has added eye protection to its list of required PPE for healthcare staff, physicians, and visitors unless physical distancing can be maintained,^79, 80^ as this was not already a requirement. **Please note that this guidance is not representative of all provinces (see note about guidance documents included in this review, page 4).**

##### Question 2C

Adjusting restrictions to and screening staff and visitors (e.g., visitor policy changes, approach to and frequency of screening)

No studies to date have reported on this outcome, but guidance documents (without citation of evidence) were located from Ontario, Saskatchewan, the UK, and Ireland. In Canada, where visitor restrictions have already been in place for some time, there were few changes in response to VOC. In the UK and Ireland, greater emphasis was placed on obtaining patients’ travel history.

###### Guidance documents related to staff and visitor screening/restrictions

- Public Health Ontario guidelines (high quality) recommend reduction of visitors to healthcare facilities in response to increased VOC cases.^72^ Similarly, in Saskatchewan (low quality), only one designated family member or support person can travel to support residents/patients in end-of-life situations or with essential care needs. Any visitor restrictions should not be based solely on the VOC situation in the visitor’s geography of origin, but rather continue to be informed by an array of vigilant screening measures.^79^ In the UK and Ireland (medium quality), more emphasis is placed on requiring a recent travel history from anyone presenting for care.^69, 70^
- In Ontario, any VOC cases originating in a healthcare facility should trigger immediate testing of all patients and staff in the affected areas.^72^
- **Please note that this guidance is not representative of all provinces (see note about guidance documents included in this review, page 4).**

##### Question 2D

Adjusting service provision based on VOC status (e.g., cohorting patients in hospitals based on the SARS-CoV-2 variants they have)

No studies to date have reported on this outcome, but guidance documents (without citation of evidence) were located from Ontario, Saskatchewan, and the UK. Notably, Ontario does not recommend cohorting B.1.1.7 patients but does recommend the use of private rooms for B.1.351 and P.1 patients, despite evidence that B.1.1.7 is the most transmissible VOC of the three.^9^

###### Guidance documents related to adjusting service provision

- In Saskatchewan, guidelines (low quality) do not recommend cohorting based on VOC diagnosis.^79^ In Ontario (high quality), cohorting is not recommended for B.1.1.7 patients, but those with B.1.351 or P.1 should be housed in private rooms where possible.^72^
- In emergency departments in the UK, it is recommended that any patients at risk of being SARS-CoV-2 positive (VOC or not) should be accommodated in a single room with ensuite bathroom facilities, regardless of their reason for presentation (medium quality).^69^
- **Please note that this guidance is not representative of all provinces (see note about guidance documents included in this review, page 4).**

##### Question 2E

Adjusting patient accommodations, shared spaces and common spaces (e.g., improvement to HVAC systems)

No studies to date have reported on this outcome, but two guidance documents recommend improving building ventilation whenever possible.

###### Guidance documents related to adjusting patient accommodations

- Public Health Ontario requires that all healthcare facilities review their HVAC systems in accordance with CSA Z317.2:19, a standard for special requirements for HVAC systems in health facilities by the Canadian Standards Association.^81^ Guidelines for the community from the UK (medium quality) also recommend improving building ventilation whenever possible, or using air-cleaning alternatives.^54^ **Please note that this guidance is not representative of all provinces (see note about guidance documents included in this review, page 4).**

## Discussion

This rapid scoping review sought to identify, appraise, and summarize evidence related to the impact of VOC known in April 2021 (B.1.1.7, B.1.351, and P.1) on public health measures and health system arrangements. Our search identified 22 articles on public health measures and 17 articles related to health system arrangements. We also identified 21 guidance documents that provided information on public health and/or health systems arrangements.

### Critical Appraisal

A total of 16 original research studies were assessed for quality (excluding 17 lab or modeling studies and 4 grey literature sources), indicating a strength of this review. While observational studies ranged in quality from low to high, the majority are preprints that have not yet been peer reviewed. Therefore, findings should be interpreted with caution to inform health system and public health recommendations. The studies which scored higher on the NOS were those which selected participants from large, representative samples and controlled for most confounders. Studies which tended to score lower on the NOS were those where self-reported data were used or when limited description of the non-respondent group was provided for cross-sectional studies. It is important to note that the NOS was originally developed for cohort and case-control studies, rather than studies of cross-sectional design. Although an adapted version of NOS was used to score cross-sectional studies,^20^ there may be some limitations in applying this adapted version. We applied an additional quality control measure to observational studies by decreasing preprint study scores by two points. While this approach of downgrading preprints provides further appraisal of study quality, it is not considered in the standard NOS scoring instructions. For the purpose of presenting the most recent evidence on this topic, it was important to include preprint studies, which are typically excluded from systematic reviews, but are an essential consideration in COVID-19 reviews.

Twenty-one guidance documents were appraised, highlighting the breadth of grey literature included in this review. No guidance documents were classified as clinical practice guidelines, but the AGREE II tool was applied to all guidance documents as it is recognised as the gold standard for quality assessment of guidelines.^19^ While we acknowledge the AGREE II tool was designed for clinical practice guidelines, due to a lack of alternative standard critical appraisal options, it was the best choice for this review. Guidance documents ranged from low to high quality, with sources consistently scoring low in Domain 3: Rigour of Development. This was largely due to the limited inclusion of methodological consideration and the heterogeneity across sources. It is important to note that within Domain 3, one item assessed whether a link between references and recommendations were made. Few sources cited original research, highlighting a gap in evidence-informed public health guidance across organizations and countries. The accuracy of quality appraisal of guidance documents could have been enhanced by including additional appraisers, however, the research team agreed that the overall scores are generally reflective of the quality of papers. Although certain guidance documents were considered low quality based on AGREE II, they may be useful when considering changes specific to settings for which the document was intended (e.g., salons, schools).

Interestingly, the majority of observational studies were related to health systems, while the majority of guidance documents were related to public health measures. Of the four observational studies which reported on public health measures, only one was considered to be high quality. This further highlights the finding that public health guidance documents tended to be based on limited evidence, or on low quality evidence at best.

### Guidance Documents

In almost all cases, guidance documents included in this review did not cite published evidence. Of note, guidance documents included in this review were found in topic areas where published evidence was scarce or not available, whereas topics with available published research were not reflected in guidance documents. Guidance included in this review may be based on evidence generated prior to the emergence of VOC, or on expert opinion.

### Public Health Measures

Eighteen studies and 4 reports contributed data relevant to modifying existing public health measures. Modeling studies were the most common design (10/18) followed by observational studies (4/18) and laboratory studies (3/18). Of note, most studies (16/18) are preprints and have not undergone peer-review. In general, although available evidence is varied and scarce, findings from the included studies overwhelmingly support the implementation of strong public health measures (i.e., lockdowns, distancing, testing, contact tracing), running in parallel with a timely vaccine schedule. The increased transmissibility of the VOC signals the need for more pre-emptive (close and then open) versus reactive (open and then close) strategies. Most studies relevant to this question focused broadly on social distancing as a strategy, with no recommendation regarding objective metrics such as time or distance or type of social distancing strategy. No studies were found that focused on modification to handwashing or masking related to the emergence of variant strains. Age and gender may be an important target to consider when developing a population vaccine promotion campaign.

### Health Systems Arrangements

Overall, there were 17 studies that reported on health system impacts, with most reporting on impact on hospitalization, ICU admissions and deaths. Among the studies that reported on the impact of VOC on hospitalization, trends suggest there is an increase in hospitalization due to B.1.1.7 but no difference in length of stay. There seems to be less agreement on the impact of B.1.1.7 on intensive care admissions, with two studies reporting increased admission to ICU with B.1.1.7 compared to the wild type, and three studies reporting no difference. While there are mixed findings on impact of VOC (B.1.1.7 and P.1) on death, six studies found an increased risk compared to three studies that reported no change. For studies that reported an increase in death, B.1.1.7 was found to increase the risk 15% to 67% compared to non-VOC, suggesting that B.1.1.7 could be linked with higher mortality than non-B.1.1.7 strains. One study reported on the effectiveness of PPE in reducing VOC transmission in the hospital. No studies reported on screening staff and visitors, adjusting service provisions (e.g., cohorting), or adjusting patient accommodations and shared spaces, which is a significant gap in the literature.

### Limitations

While this rapid scoping review has several strengths, there are limitations that must be acknowledged. First, due to the rapid production of the literature on COVID-19 and VOC, most of the studies included in this review were preprints, and have thus not yet undergone peer review. As mentioned above, this must be considered when interpreting the findings.

Additionally, our search strategy was limited to articles that specified reporting on one of the recognized VOC (B.1.1.7, P.1, or B.1.351). Given the growing trend that VOC are replacing the wild type as the dominant strain as well as the continued emergence of other variants of interest, future consideration of expanding the search strategy may be warranted. For example, this review did not consider the variant of interest that is emerging in India (B.1.617) or other variants of interest which may warrant future evaluation as these situations evolve.^82^

A third limitation is that our review is limited to studies that reported specifically on VOC, which makes it difficult to interpret some of the findings without taking into consideration the wider literature on SARS-CoV-2. For example, we report on attitudes towards vaccines only in context of VOC, without wider acknowledgement of the extensive body of literature on vaccine hesitancy. Canadian provincial and national guidance documents were also excluded if they did not specify recommendations for VOC.

Finally, the identification of guidance documents through the grey literature search may have been limited by different jurisdictional guidance document indexing protocols and practices. As such, the relevance of guidance documents included in this review may vary depending on different contexts across jurisdictions and do not necessarily reflect the breadth and scope of available guidance from across Canada. A comprehensive jurisdictional scan of provincial guidance documents is needed to address this gap.

### Research Gaps

As evident in this rapid review, the nature of and findings on the impact of VOC on public health measures and health systems arrangements are quickly changing and emerging. We have identified several specific research gaps that need to be addressed to provide more robust evidence around public health measures and health system arrangement decisions.

1. Evidence is needed related to best practices for screening staff and visitors in health service organizations and adjusting service provisions
2. Evidence is needed to support/refute adjusting patient accommodations and shared spaces in a hospital setting with the presence of different strains
3. Standardized approaches and tools are needed to track adherence to different public health measures
4. Methods for appraising modeling studies need to be developed
5. Novel methods to collect and analyze data are needed to inform infection-prevention strategies for safer workplace environments with the emergence of highly transmissible strains
6. Standards for sharing surveillance data nationally to rapidly inform health policy and health system guidance documents are recommended worldwide
7. Information to support changes in guidance related to masking in light of more transmissible strains is needed
8. Evidence to guide best practice standards for screening and testing for variants of concern under different conditions is needed
9. Need for comprehensive jurisdictional scan to identify, compare, and contrast provincial strategies and guidelines

## Conclusion

This rapid scoping review provides synthesized evidence related to the public health and health system impacts of the three major SARS-CoV-2 VOC (B.1.1.7, B.1.351, and P.1). While the findings should be interpreted with caution as most of the sources identified were preprints, findings suggest a combination of non-pharmaceutical interventions (e.g., masking, physical distancing, lockdowns, testing, contact tracing) should be employed alongside a vaccine strategy to improve population and health system outcomes. Additionally, while the findings are mixed on the impact of VOC on health systems arrangements, the evidence is trending towards increased hospitalization and death.

## Data Availability

Data is available upon reasonable request from corresponding author.

## Funding Acknowledgement(s)

The SPOR Evidence Alliance (<u>SPOR EA</u>) is supported by the Canadian Institutes of Health Research (<u>CIHR</u>) under the Strategy for Patient-Oriented Research (<u>SPOR</u>) initiative.

COVID-19 Evidence Network to support Decision-making (<u>COVID-END</u>) is supported by the Canadian Institutes of Health Research (<u>CIHR</u>) through the Canadian 2019 Novel Coronavirus (COVID-19) Rapid Research Funding opportunity.

## Project Contributors

Janet Curran, Dalhousie University, Co-Investigator

Justine Dol, Dalhousie University, Research Coordinator

Leah Boulos, MSSU, Evidence Synthesis Coordinator

Mari Somerville, Dalhousie University, Postdoctoral Fellow

Holly McCulloch, IWK Health Centre, Project Administrator

Bearach Reynolds, Mb BCh BAO BA MRCPI, ESI Fellow

Allyson Gallant, Dalhousie University, Research Assistant

Lynora Saxinger, University of Alberta, Content Expert

Alexander Doroshenko, Faculty of Medicine & Dentistry, University of Alberta

Danielle Shin, Dalhousie University, Research Assistant

Helen Wong, Dalhousie University, Research Assistant

Daniel Crowther, Dalhousie University, Research Assistant

Marilyn Macdonald, JBI Centre of Excellence, School of Nursing, Dalhousie University

Ruth Martin-Misener, JBI Centre of Excellence, School of Nursing, Dalhousie University

Jill Hayden, Dalhousie University, Quality Appraisal Methods

Jason LeBlanc PhD FCCM, D(ABMM), Director of Virology, Immunology, Molecular Microbiology, Content Expert

Lisa Barrett MD PhD FRCPC, Clinician Scientist, Infectious Diseases, NSHA, Content Expert

Jeannette Comeau MD MSc FRCPC FAAP, Pediatric Infectious Diseases Consultant, Content Expert

Third-Party Materials

If you wish to reuse non-textual material from this report that is attributed to a third party, such as tables, figures or images, it is your responsibility to determine whether permission is required for such use and to obtain necessary permission from the copyright holder. The risk of claims resulting from infringement of any third-party-owned material rests solely with the user.

## General Disclaimer

This report was prepared by Nova Scotia COVID-END Evidence Synthesis Group on behalf of the SPOR Evidence Alliance and COVID-END. It was developed through the analysis, interpretation and synthesis of scientific research and/or health technology assessments published in peer-reviewed journals, institutional websites and other distribution channels. It also incorporates selected information provided by experts and patient partners with lived experience on the subject matter. This document may not fully reflect all the scientific evidence available at the time this report was prepared. Other relevant scientific findings may have been reported since completion of this synthesis report.

SPOR Evidence Alliance, COVID-END and the project team make no warranty, express or implied, nor assume any legal liability or responsibility for the accuracy, completeness, or usefulness of any information, data, product, or process disclosed in this report. Conclusions drawn from, or actions undertaken on the basis of, information included in this report are the sole responsibility of the user.

## Definitions & Abbreviations

ABM: Agent-based Model
AGREE II: Appraisal of Guidelines Research and Evaluation
aIRR: adjusted incidence rate ratios
B.1.1.7: variant of concern originating in the United Kingdom, also known as VUI 202012/01 and VOC 202012/01
B.1.351: variant of concern originating in South Africa, also known as 20H/501Y.V2
BMI: body mass index
CanCOGen: Canadian COVID Genomics Network
CENTRAL: Central Register of Controlled Trials
CFR: Case Fatality Rates
CI: confidence interval
CIDRAP: Center for Infectious Disease Research and Policy
Ct: cycle threshold, provides a relative measure of viral quantity
COG-UK: COVID-19 Genomics UK
CDSR: Cochrane Database of Systematic Reviews
dQALY: discounted quality-adjusted life years
E484K: escape mutation in the SARS-CoV-2 virus, present in B.1.1.7
ECDC: European Centres for Disease Control
FDA: Food and Drug Administration
HCW: healthcare workers
HR: Hazard Ratio
HVAC: heating, ventilation, and air conditioning
ICU: Intensive Care Unit
IQR: interquartile range
IR: incidence rate
LOS: length of stay
mRNA: messenger ribonucleic acid
NGS: next generation sequencing
NOS: Newcastle-Ottawa scale
NPI: Non-Pharmaceutical Interventions
NRW: North-Rhine Westphalia
OR: odds ratio
P.1: variant of concern originating in Brazil, also known as B.1.28.1
PCR: polymerase chain reaction, method for DNA replication and genome sequencing
PHU: Public Health Unit
PPE: personal protective equipment
PR: prevalence ratio
R: reproduction
R_0_: basic reproduction number, expected number of cases generated by one case in a population when everyone is susceptible to infection
R_t_: effective reproduction number
RR: risk ratio
RT-LAMP: reverse transcription loop-mediated and transcription-mediated amplification isothermal amplification
RTD: Rapid Antigen Test
SA: South Africa
SAPSII: severity score at admission
SIDARTHE: a type of model
SGTF: spike OR S gene target failure, correlates with the increase of confirmed, sequenced variants
SGTL: spike gene late detection
SD: standard deviation
UK: United Kingdom
US: United States
VE: vaccine efficiency
VOC: variant of concern
WHO: World Health Organization
WGS: whole genome sequencing

## Appendix 1 Literature Search Strategy

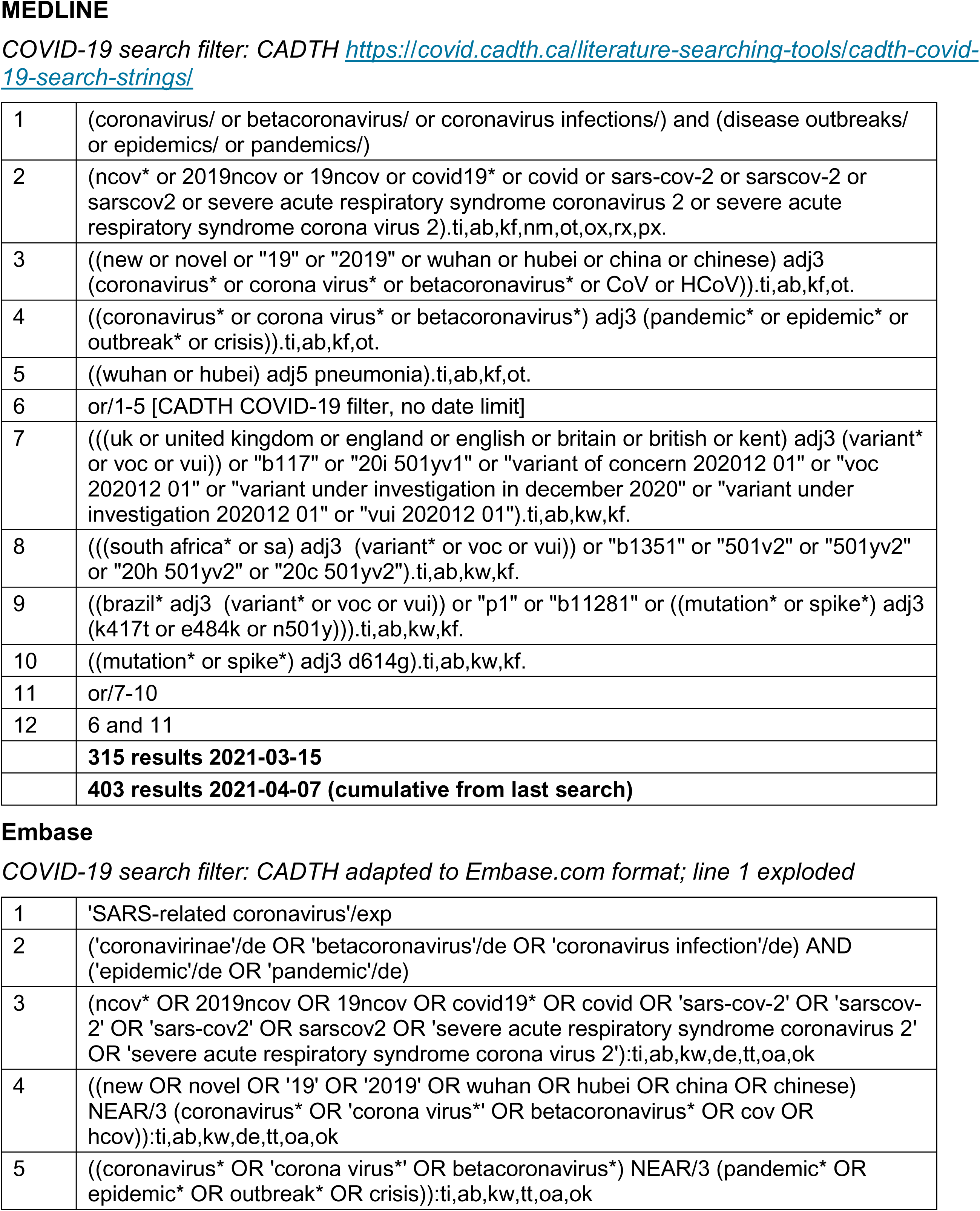
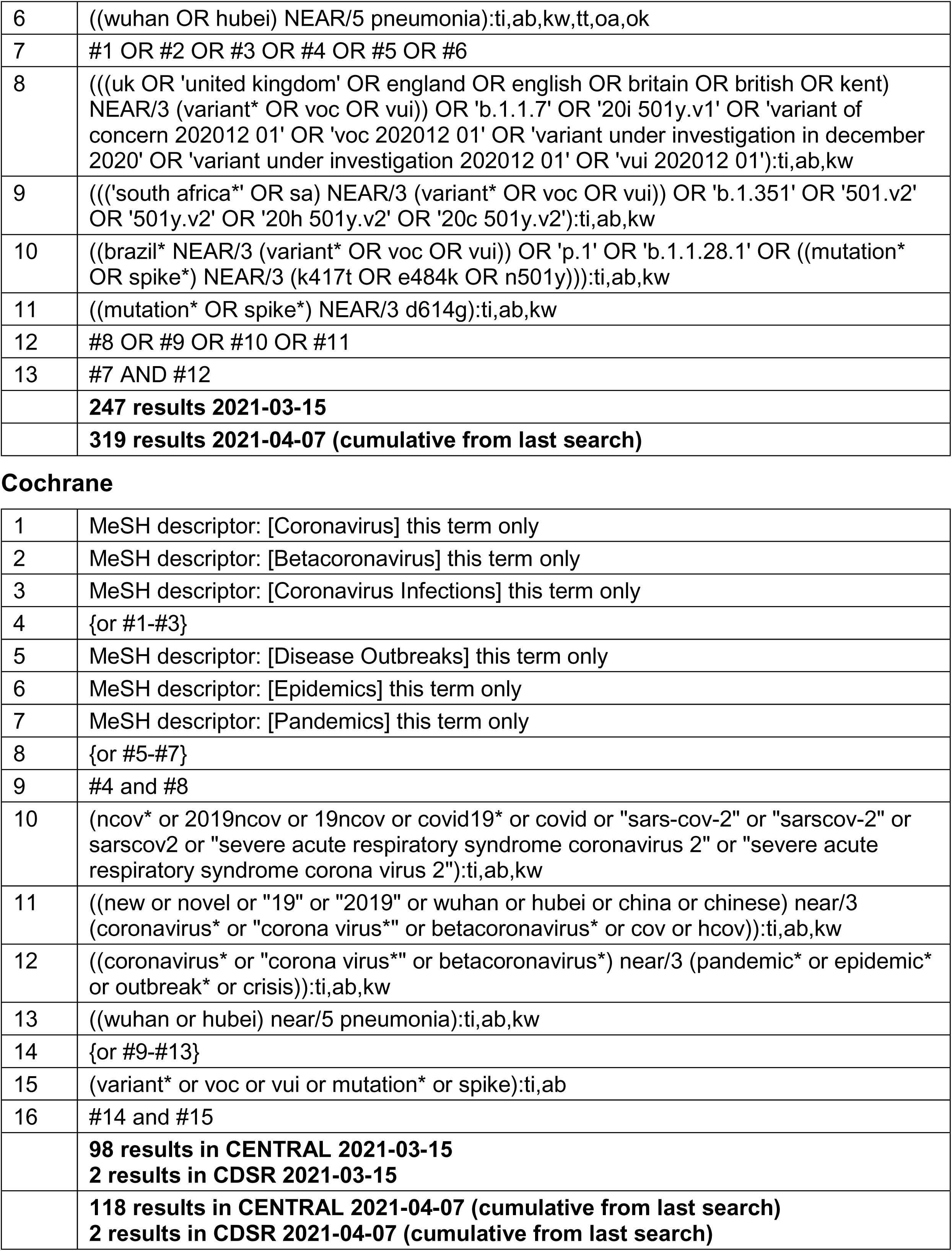

|  |  |
| --- | --- |
| 1 | (coronavirus/ or betacoronavirus/ or coronavirus infections/) and (disease outbreaks/ or epidemics/ or pandemics/) |
| 2 | (ncov* or 2019ncov or 19ncov or covid19* or covid or sars-cov-2 or sarscov-2 or sarscov2 or severe acute respiratory syndrome coronavirus 2 or severe acute respiratory syndrome corona virus 2).ti,ab,kf,nm,ot,ox,rx,px. |
| 3 | ((new or novel or "19" or "2019" or wuhan or hubei or china or chinese) adj3 (coronavirus* or corona virus* or betacoronavirus* or CoV or HCoV)).ti,ab,kf,ot. |
| 4 | ((coronavirus* or corona virus* or betacoronavirus*) adj3 (pandemic* or epidemic* or outbreak* or crisis)).ti,ab,kf,ot. |
| 5 | ((wuhan or hubei) adj5 pneumonia).ti,ab,kf,ot. |
| 6 | or/1-5 [CADTH COVID-19 filter, no date limit] |
| 7 | ((((uk or united kingdom or england or english or britain or british or kent) adj3 (variant* or voc or vui)) or "b117" or "20i 501yv1" or "variant of concern 202012 01" or "voc 202012 01" or "variant under investigation in december 2020" or "variant under investigation 202012 01" or "vui 202012 01")).ti,ab,kw,kf. |
| 8 | ((((south africa* or sa) adj3 (variant* or voc or vui)) or "b1351" or "501v2" or "501yv2" or "20h 501yv2" or "20c 501yv2").ti,ab,kw,kf. |
| 9 | ((brazil* adj3 (variant* or voc or vui)) or "p1" or "b11281" or ((mutation* or spike*) adj3 (k417t or e484k or n501y))).ti,ab,kw,kf. |
| 10 | ((mutation* or spike*) adj3 d614g).ti,ab,kw,kf. |
| 11 | or/7-10 |
| 12 | 6 and 11 |
|  | **315 results 2021-03-15** |
|  | **403 results 2021-04-07 (cumulative from last search)** |
| 1 | 'SARS-related coronavirus'/exp |
| 2 | ('coronavirinae'/de OR 'betacoronavirus'/de OR 'coronavirus infection'/de) AND ('epidemic'/de OR 'pandemic'/de) |
| 3 | (ncov* OR 2019ncov OR 19ncov OR covid19* OR covid OR 'sars-cov-2' OR 'sarscov-2' OR 'sars-cov2' OR sarscov2 OR 'severe acute respiratory syndrome coronavirus 2' OR 'severe acute respiratory syndrome corona virus 2'):ti,ab,kw,de,tt,oa,ok |
| 4 | ((new OR novel OR '19' OR '2019' OR wuhan OR hubei OR china OR chinese) NEAR/3 (coronavirus* OR 'corona virus*' OR betacoronavirus* OR cov OR hcov)):ti,ab,kw,de,tt,oa,ok |
| 5 | ((coronavirus* OR 'corona virus*' OR betacoronavirus*) NEAR/3 (pandemic* OR epidemic* OR outbreak* OR crisis)):ti,ab,kw,tt,oa,ok |
| 6 | ((wuhan OR hubei) NEAR/5 pneumonia):ti,ab,kw,tt,oa,ok |
| 7 | #1 OR #2 OR #3 OR #4 OR #5 OR #6 |
| 8 | ((uk OR 'united kingdom' OR england OR english OR britain OR british OR kent) NEAR/3 (variant* OR voc OR vui)) OR 'b.1.1.7' OR '20i 501y.v1' OR 'variant of concern 202012 01' OR 'voc 202012 01' OR 'variant under investigation in december 2020' OR 'variant under investigation 202012 01' OR 'vui 202012 01'):ti,ab,kw |
| 9 | ((('south africa*' OR sa) NEAR/3 (variant* OR voc OR vui)) OR 'b.1.351' OR '501.v2' OR '501y.v2' OR '20h 501y.v2' OR '20c 501y.v2'):ti,ab,kw |
| 10 | ((brazil* NEAR/3 (variant* OR voc OR vui)) OR 'p.1' OR 'b.1.1.28.1' OR ((mutation* OR spike*) NEAR/3 (k417t OR e484k OR n501y))):ti,ab,kw |
| 11 | ((mutation* OR spike*) NEAR/3 d614g):ti,ab,kw |
| 12 | #8 OR #9 OR #10 OR #11 |
| 13 | #7 AND #12 |
|  | **247 results 2021-03-15** |
|  | **319 results 2021-04-07 (cumulative from last search)** |
| Cochrane |  |
| 1 | MeSH descriptor: [Coronavirus] this term only |
| 2 | MeSH descriptor: [Betacoronavirus] this term only |
| 3 | MeSH descriptor: [Coronavirus Infections] this term only |
| 4 | {or #1-#3} |
| 5 | MeSH descriptor: [Disease Outbreaks] this term only |
| 6 | MeSH descriptor: [Epidemics] this term only |
| 7 | MeSH descriptor: [Pandemics] this term only |
| 8 | {or #5-#7} |
| 9 | #4 and #8 |
| 10 | (ncov* or 2019ncov or 19ncov or covid19* or covid or "sars-cov-2" or "sarscov-2" or sarscov2 or "severe acute respiratory syndrome coronavirus 2" or "severe acute respiratory syndrome corona virus 2"):ti,ab,kw |
| 11 | ((new or novel or "19" or "2019" or wuhan or hubei or china or chinese) near/3 (coronavirus* or "corona virus*" or betacoronavirus* or cov or hcov)):ti,ab,kw |
| 12 | ((coronavirus* or "corona virus*" or betacoronavirus*) near/3 (pandemic* or epidemic* or outbreak* or crisis)):ti,ab,kw |
| 13 | ((wuhan or hubei) near/5 pneumonia):ti,ab,kw |
| 14 | {or #9-#13} |
| 15 | (variant* or voc or vui or mutation* or spike):ti,ab |
| 16 | #14 and #15 |
|  | **98 results in CENTRAL 2021-03-15**<br>**2 results in CDSR 2021-03-15** |
|  | **118 results in CENTRAL 2021-04-07 (cumulative from last search)**<br>**2 results in CDSR 2021-04-07 (cumulative from last search)** |

**Epistemonikos**

Basic search of the following terms within the LOVE:

variant* OR voc OR vui OR “B.1.1.7” OR “20I/501Y.V1” OR “202012/01” OR “B.1.351” OR “501.V2” OR “501Y.V2” OR “20H/501Y.V2” OR “20C/501Y.V2” OR “P.1” OR “B.1.1.28.1” OR “K417T” OR “E484K” OR “N501Y” OR “D614G”

**1102 results 2021-03-15**

**1330 results 2021-04-07 (cumulative from last search)**

**medRxiv / bioRxiv**

medRxiv and bioRxiv simultaneous search; Date limit: October 1 2020 – present; Title and Abstract search; All words (unless otherwise specified); 50 per page; Best Match; **round 2, update date limit February 1 2021 – present; round 3, update date limit March 1 2021 – present**

**315 unique results total (all searches) up to and including 2021-04-07**

Searches:

|  |
| --- |
| uk variant<br>united kingdom variant<br>england variant<br>english variant<br>britain variant<br>british variant<br>kent variant<br>south africa variant<br>brazil variant<br>variant of concern (*phrase search*)<br>variants of concern (*phrase search*)<br>B.1.1.7<br>20I/501Y.V1<br>202012/01<br>B.1.351<br>501.V2<br>501Y.V2<br>20H/501Y.V2<br>20C/501Y.V2<br>P.1<br>B.1.1.28.1<br>K417T<br>E484K<br>N501Y<br>D614G |

**Google**

Google screening inclusion/exclusion criteria:

1. Exclude results from academic journals (e.g. JAMA)
2. Exclude results from PubMed
3. Exclude news articles, but check to see if they link to studies, preprints, or public health guidance
4. Include results from preprint servers (e.g. medRxiv; Research Square)

At time of record capture, indicate the **specific variant** (or combination), and whether it is related to any combination of the following using the grey lit tracking form:

▪ Transmission
▪ Public health measures
▪ Health systems arrangements

|  |
| --- |
| uk variant \| united kingdom variant \| british variant \| kent variant \| uk voc \| united kingdom voc \| british voc \| kent voc \| B117 \| B.1.1.7 \| 20I/501Y.V1 \| 202012/01 \| 501Y.V1 |
| south africa variant \| south africa voc \| B1351 \| B.1.351 \| 501.V2 \| 501Y.V2 \| 20H/501Y.V2 \| 20C/501Y.V2 |
| brazil variant \| brazil voc \| P1 variant \| P1 voc \| P.1 variant \| P.1 voc \| B11281 \| B.1.1.28.1 |
| K417T \| E484K \| N501Y \| D614G |
| variants of concern \| COVID-19 variants \| SARS-CoV-2 variants |

**Twitter**

Basic search; Top results

|  |
| --- |
| uk variant |
| uk voc |
| B.1.1.7 |
| B117 |
| south africa variant |
| south african variant |
| sa variant |
| south africa voc |
| south african voc |
| sa voc |
| B.1.351 |
| B1351 |
| brazil variant |
| brazilian variant |
| brazil voc |
| brazilian voc |
| P.1 |
| variants of concern |
| covid-19 voc |
| covid19 voc |
| covid voc |
| sars-cov-2 voc |

**Other websites**

Manual searches of the following websites for terms like:

- variants
- surveillance data

|  |  |
| --- | --- |
| WHO | https://www.who.int/ |
| Canadian government websites (all) – includes Health Canada and PHAC | https://www.canada.ca/en.html |
| CDC | https://www.cdc.gov/ |
| CADTH COVID-19 Evidence Portal | https://covid.cadth.ca/ |
| Public Health England | https://www.gov.uk/government/organisations/public-health-england |
| South Africa Dept of Health | http://www.health.gov.za/ |
| Brazil – Ministério da Saúde<br>*search "variante" and check the box that says "Apenas em Ministério da Saúde" | https://www.gov.br/saude/pt-br |
| NZ Ministry of Health | https://www.health.govt.nz/ |
| Australian Dept of Health | https://www.health.gov.au/ |
| Italy - Dipartimento della Protezione Civile<br>*Italian: "variante" | http://www.protezionecivile.gov.it/ |

## Appendix 2 PRISMA Diagram

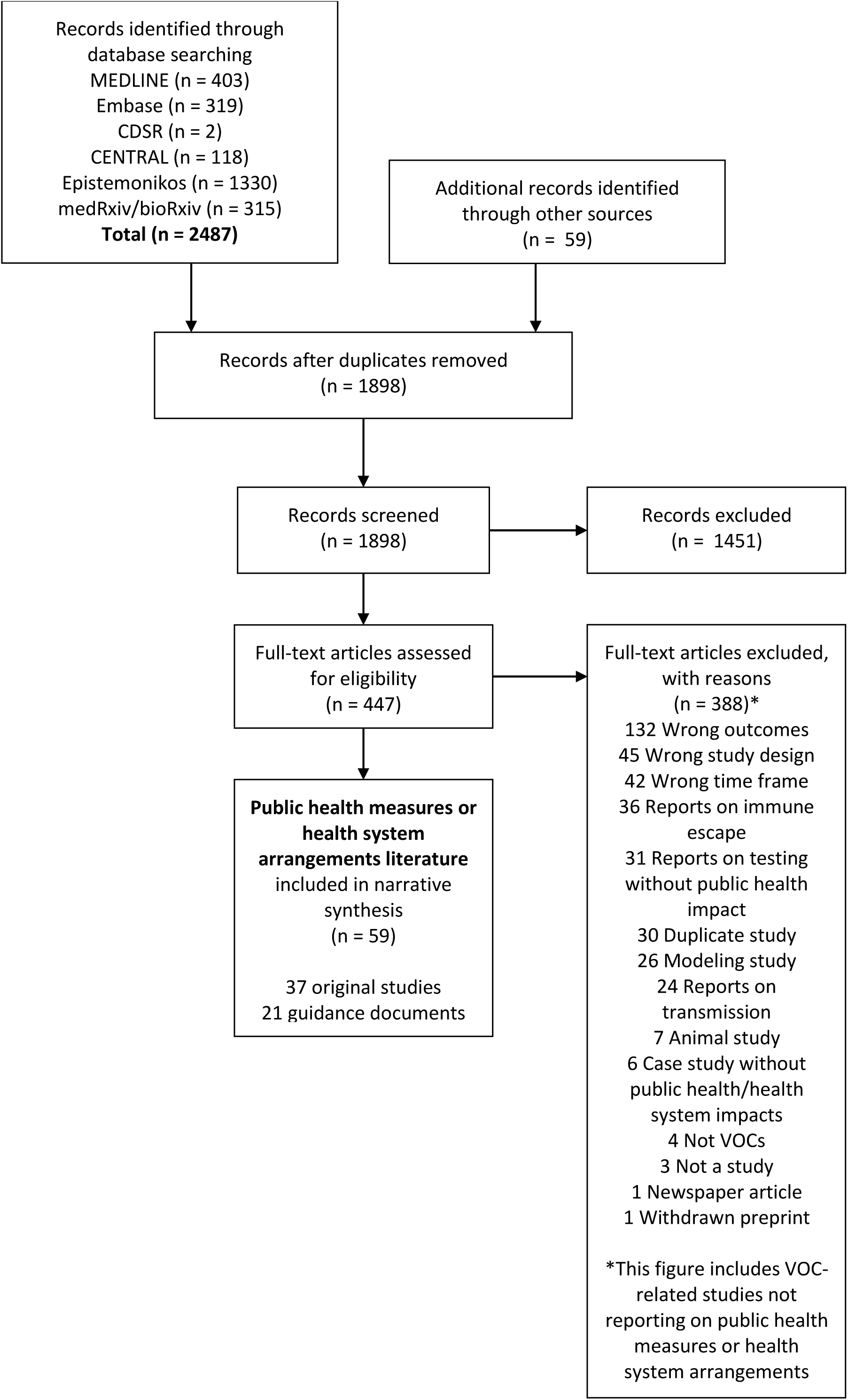

## Appendix 3 Summary of Findings Tables for Public Health Sources

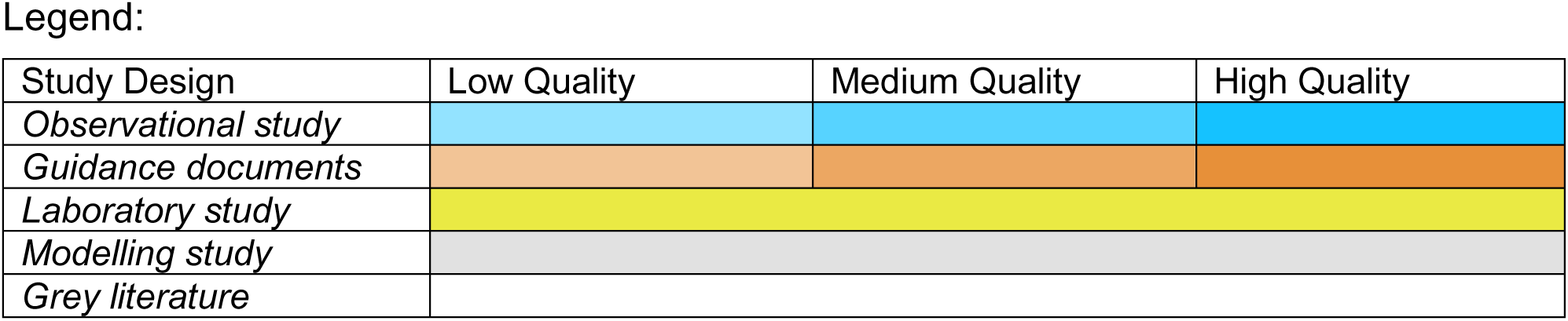

**Table 1:**
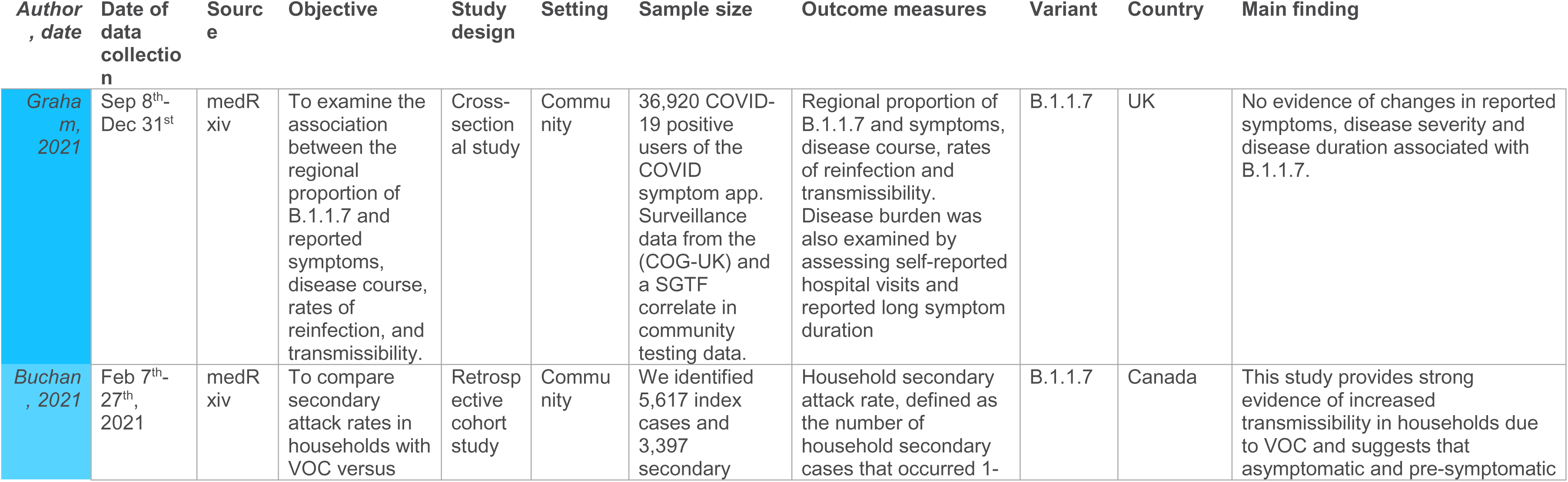
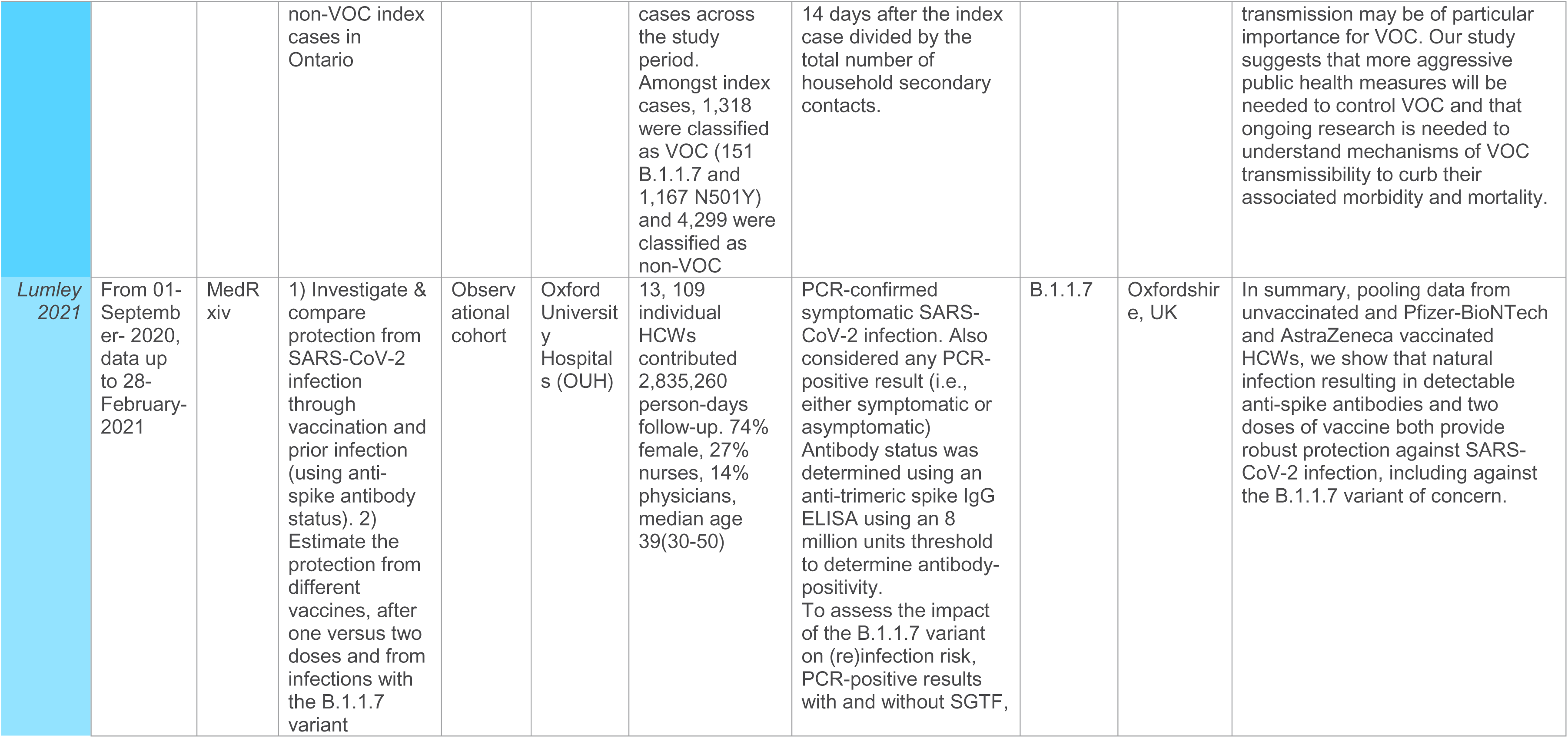
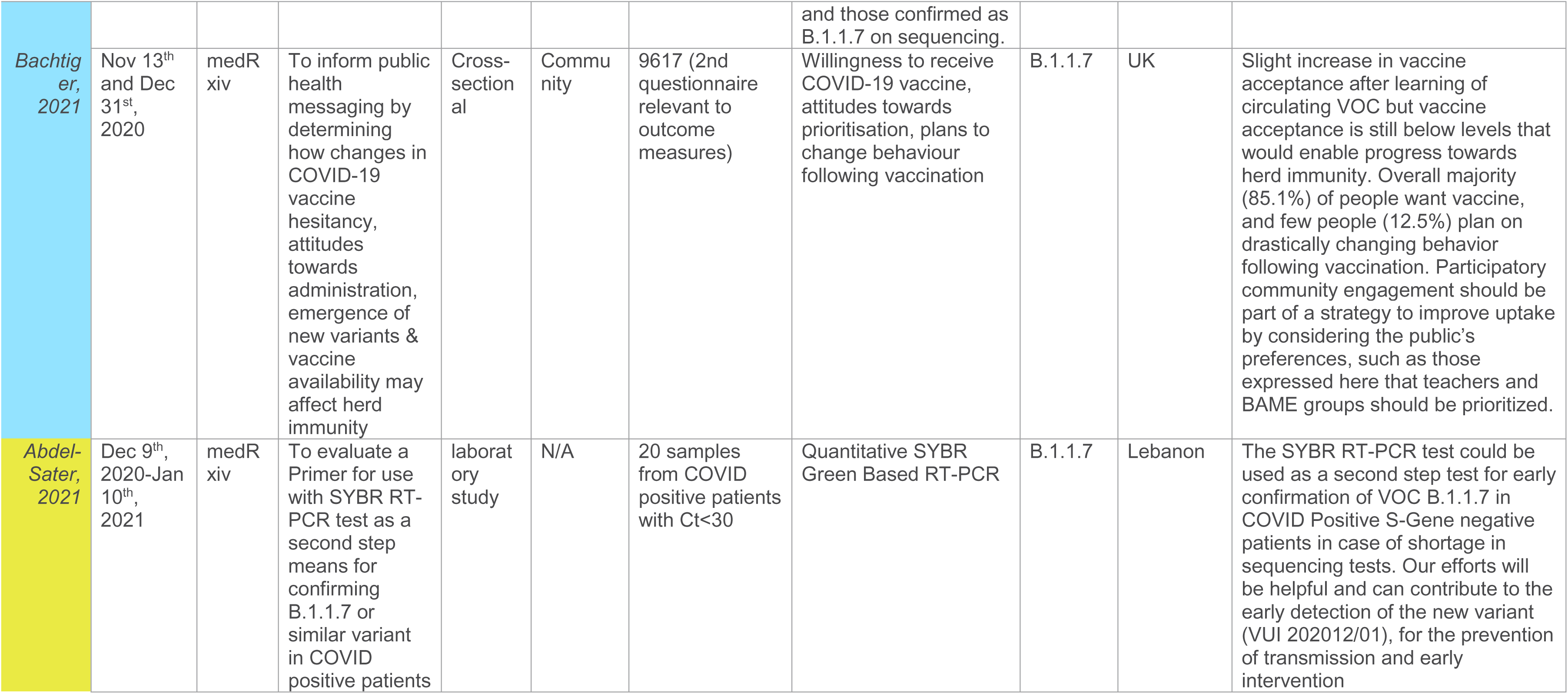
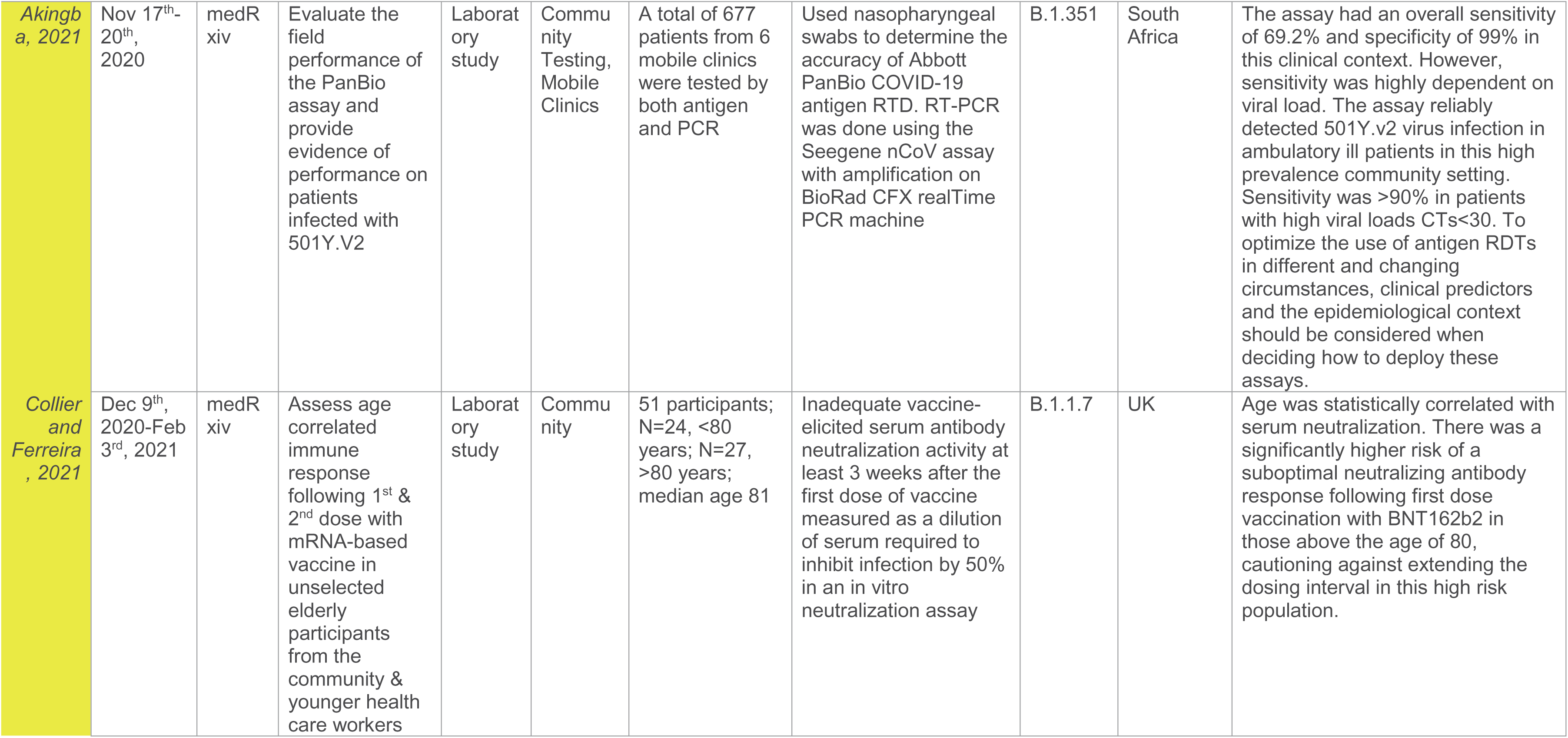
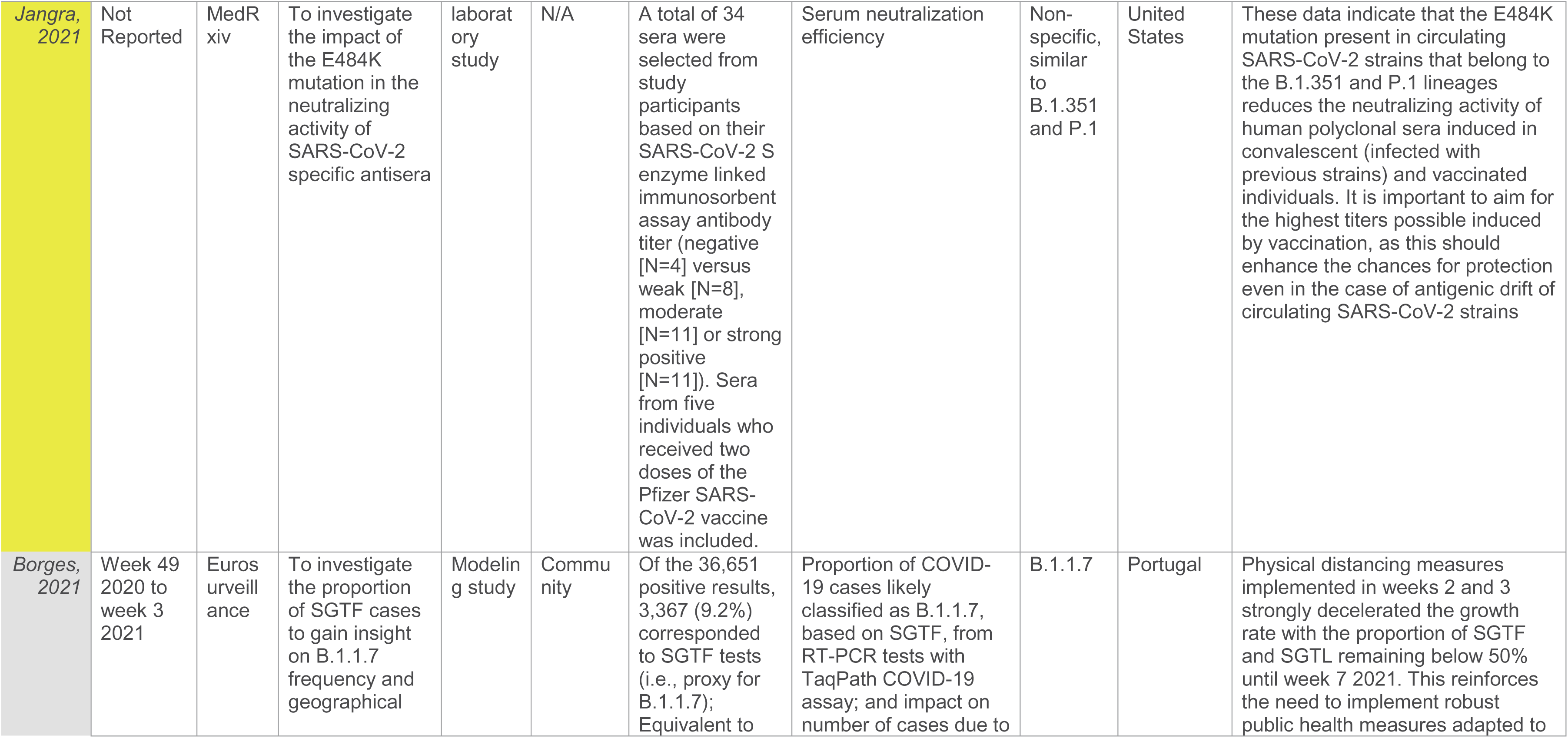
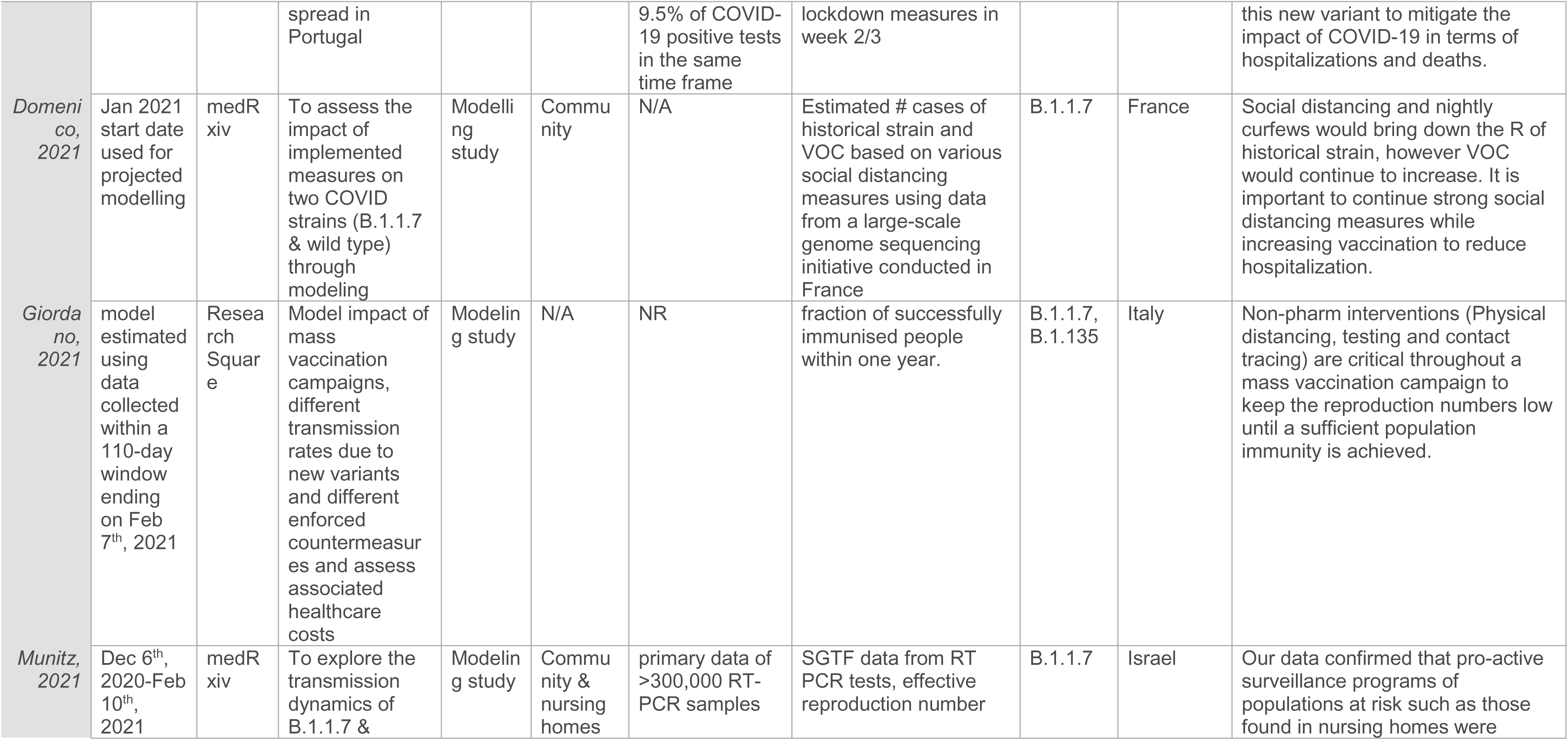
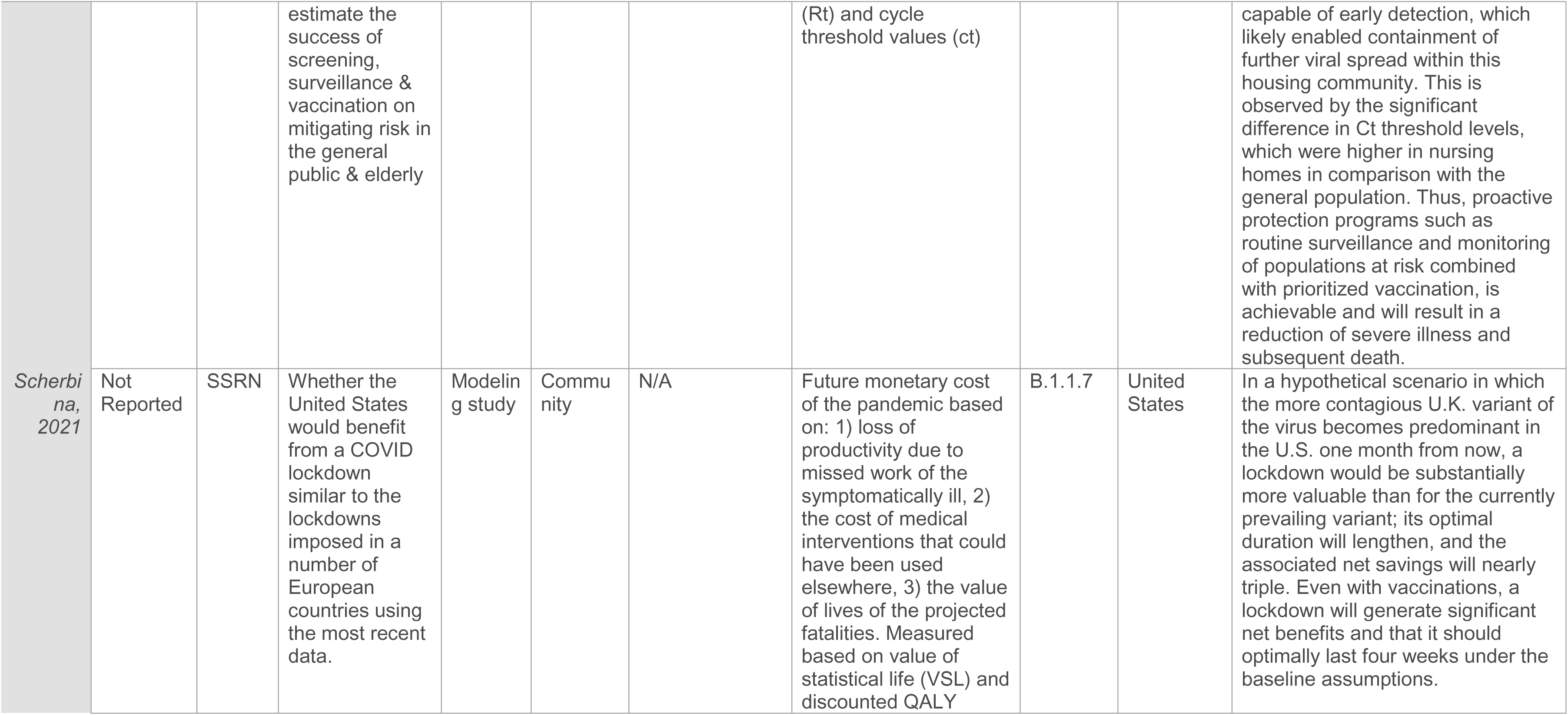
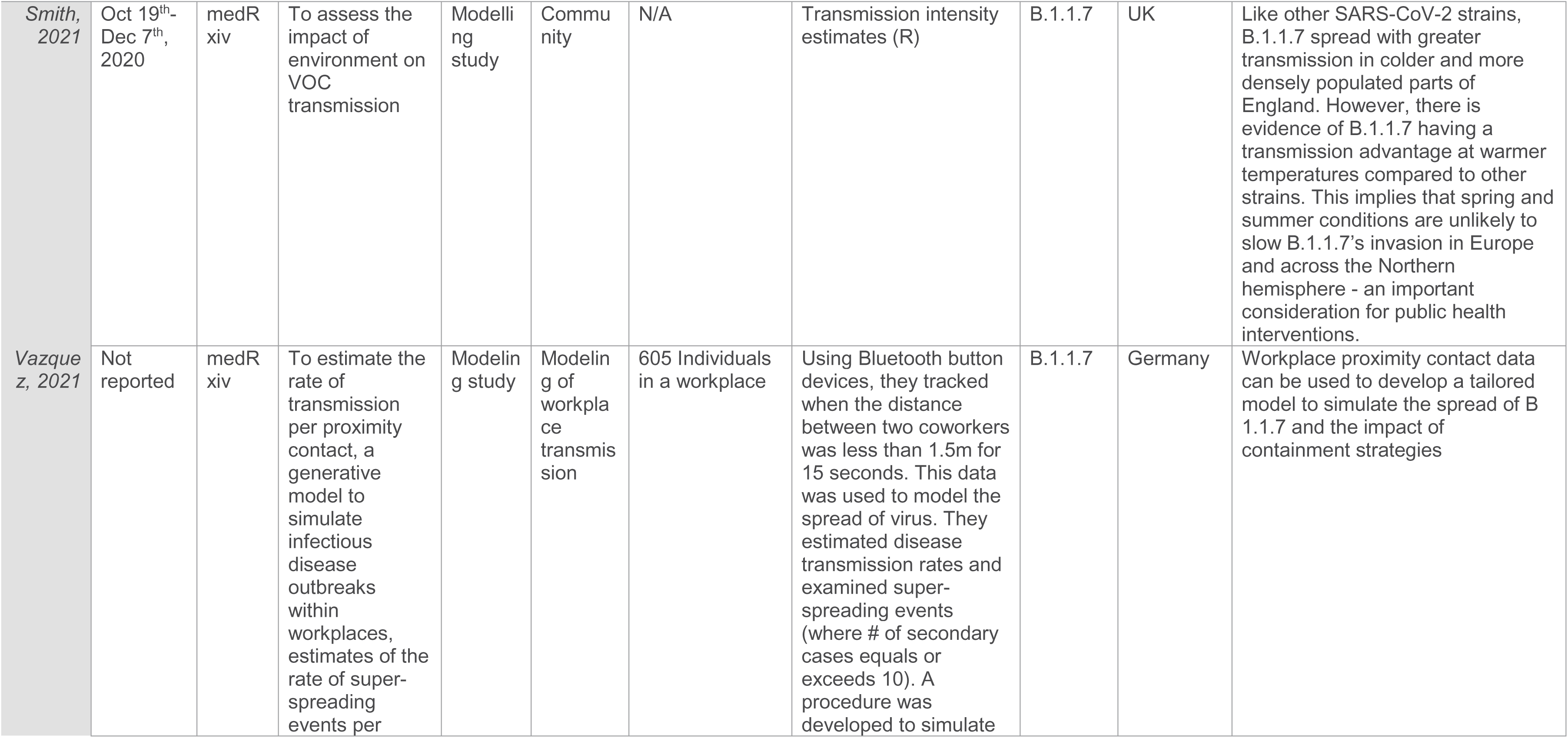
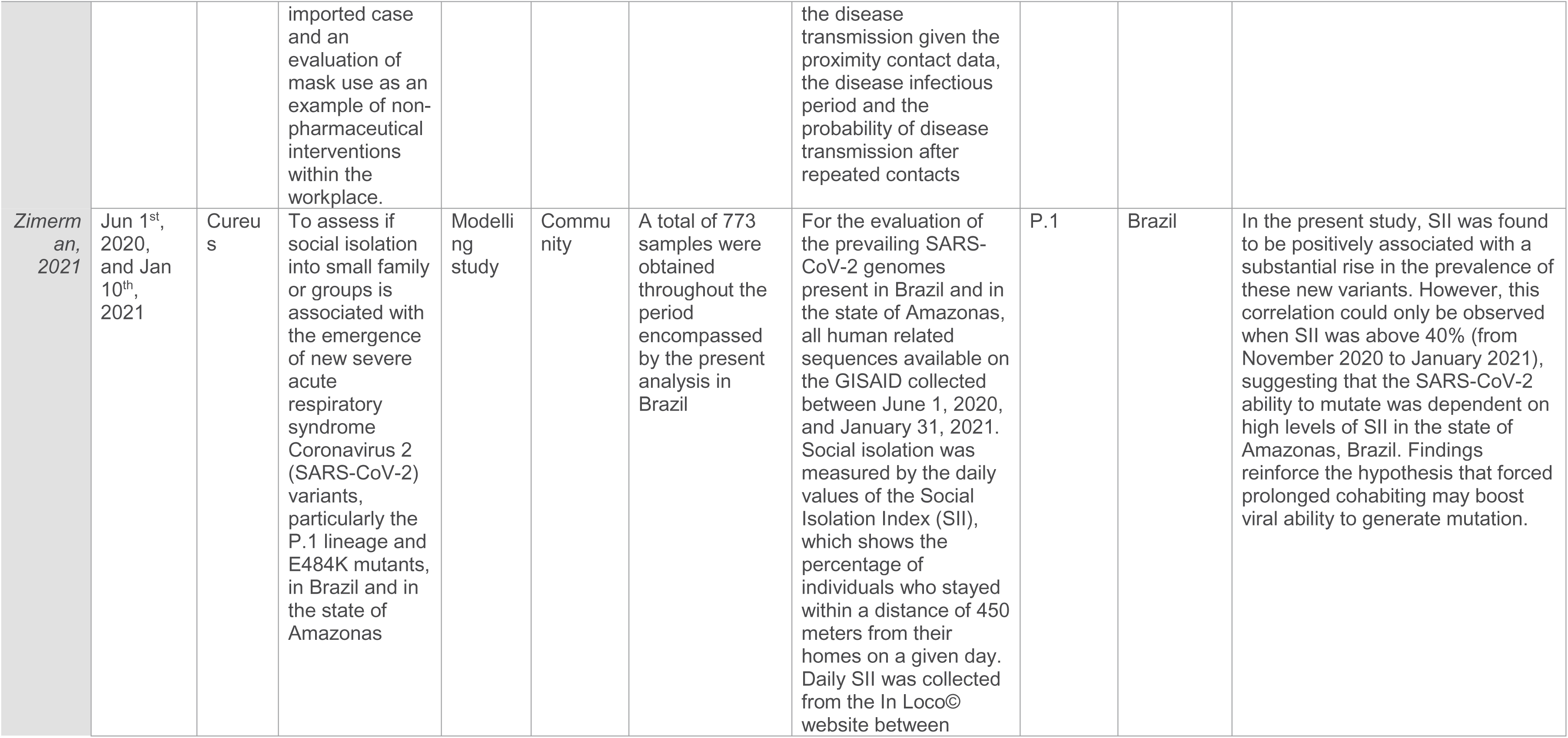
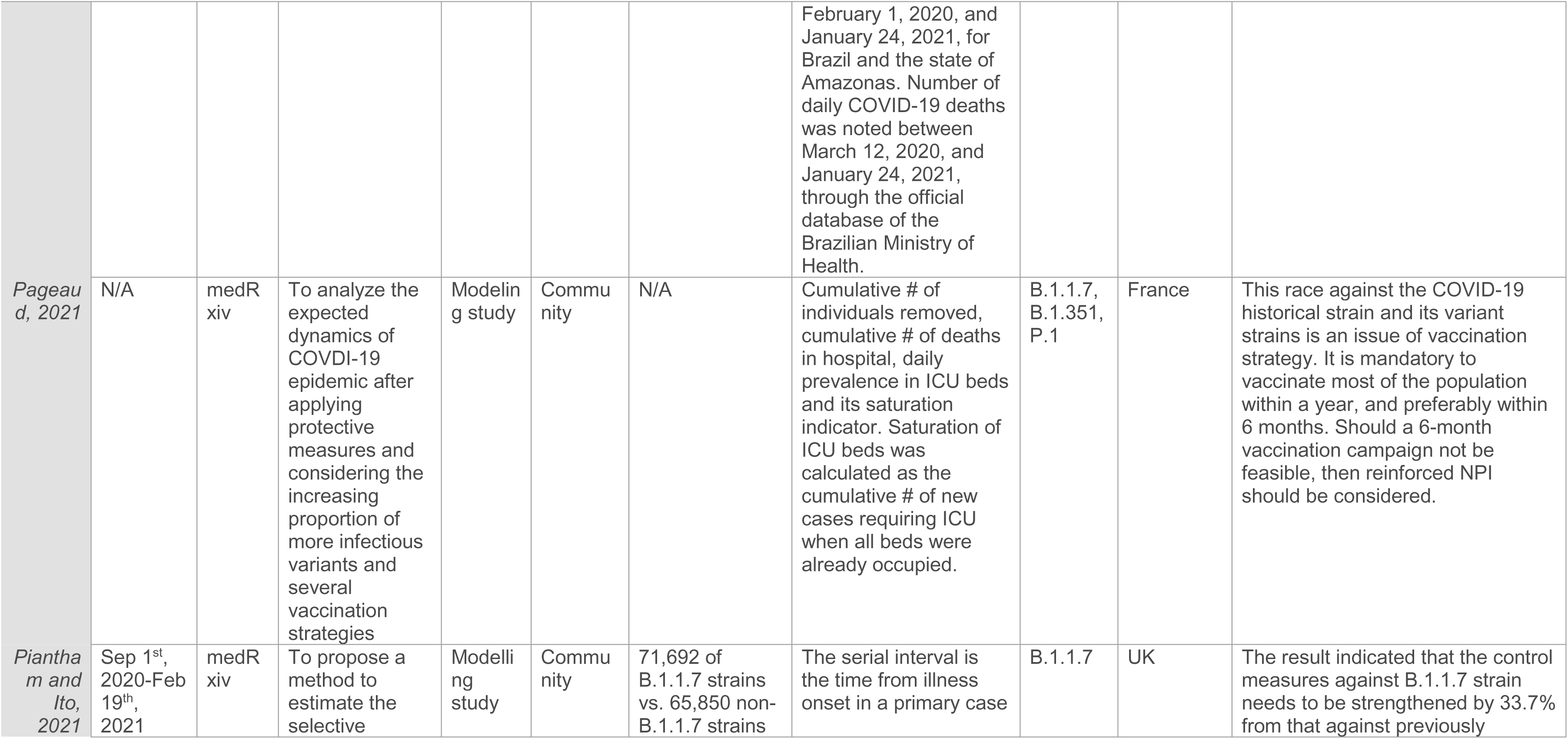
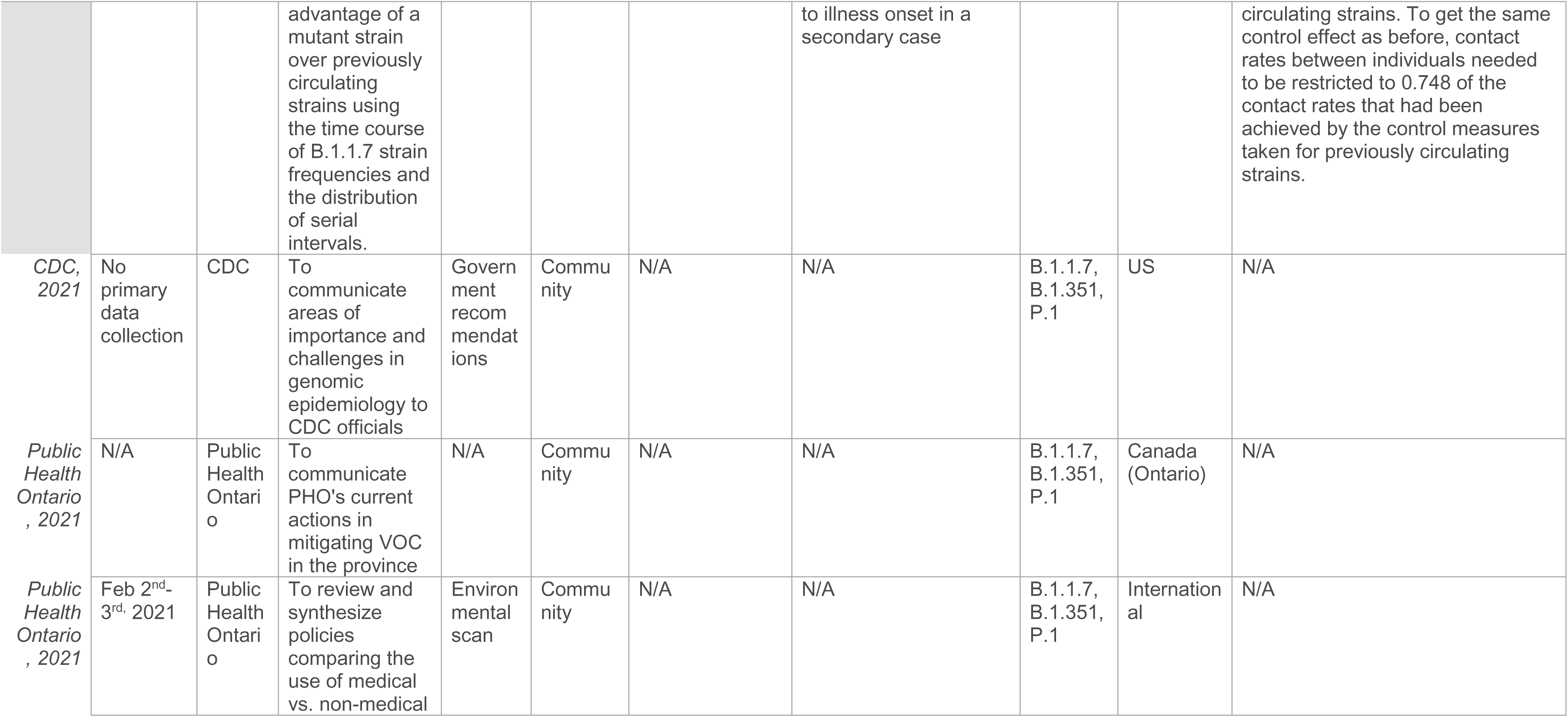
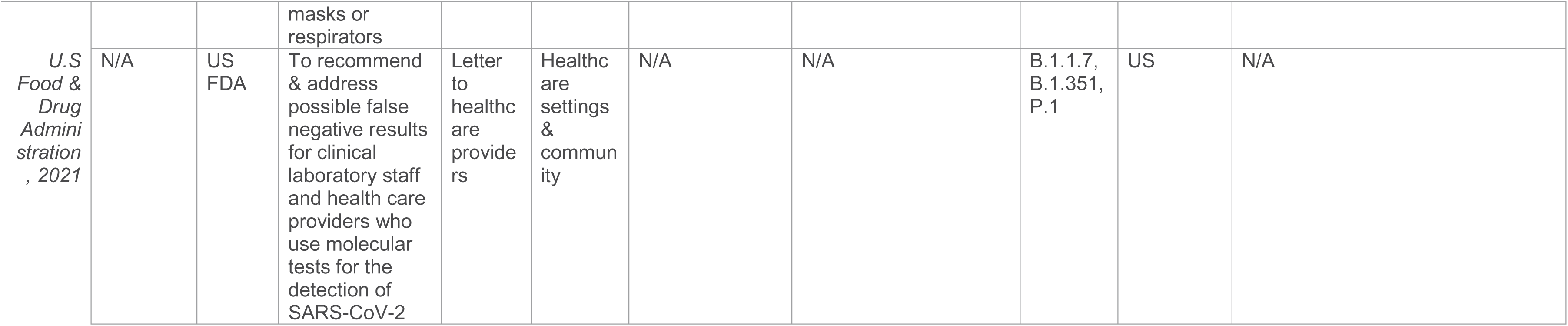
Summary of Public Health Sources

**Table 2:**
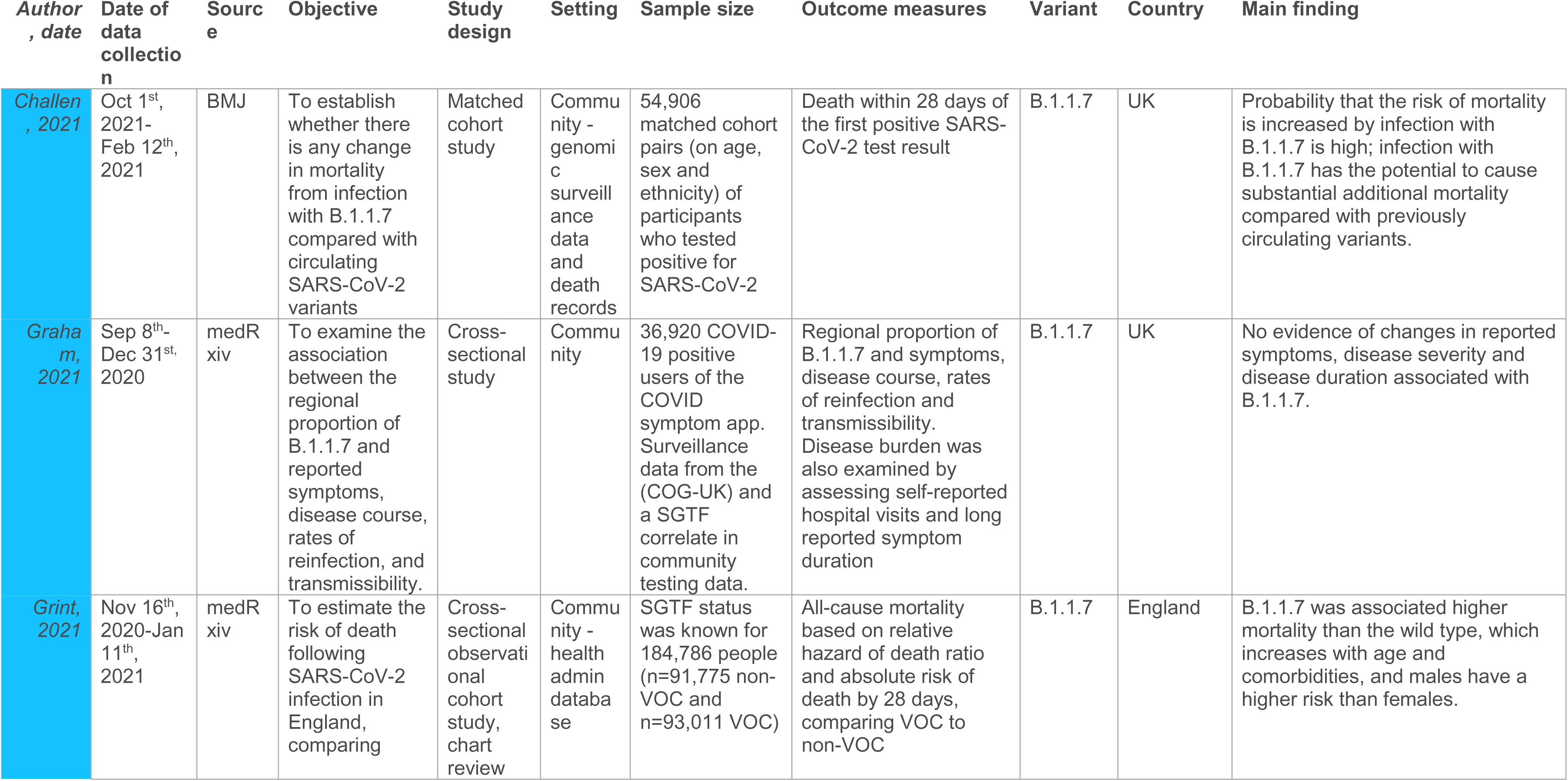
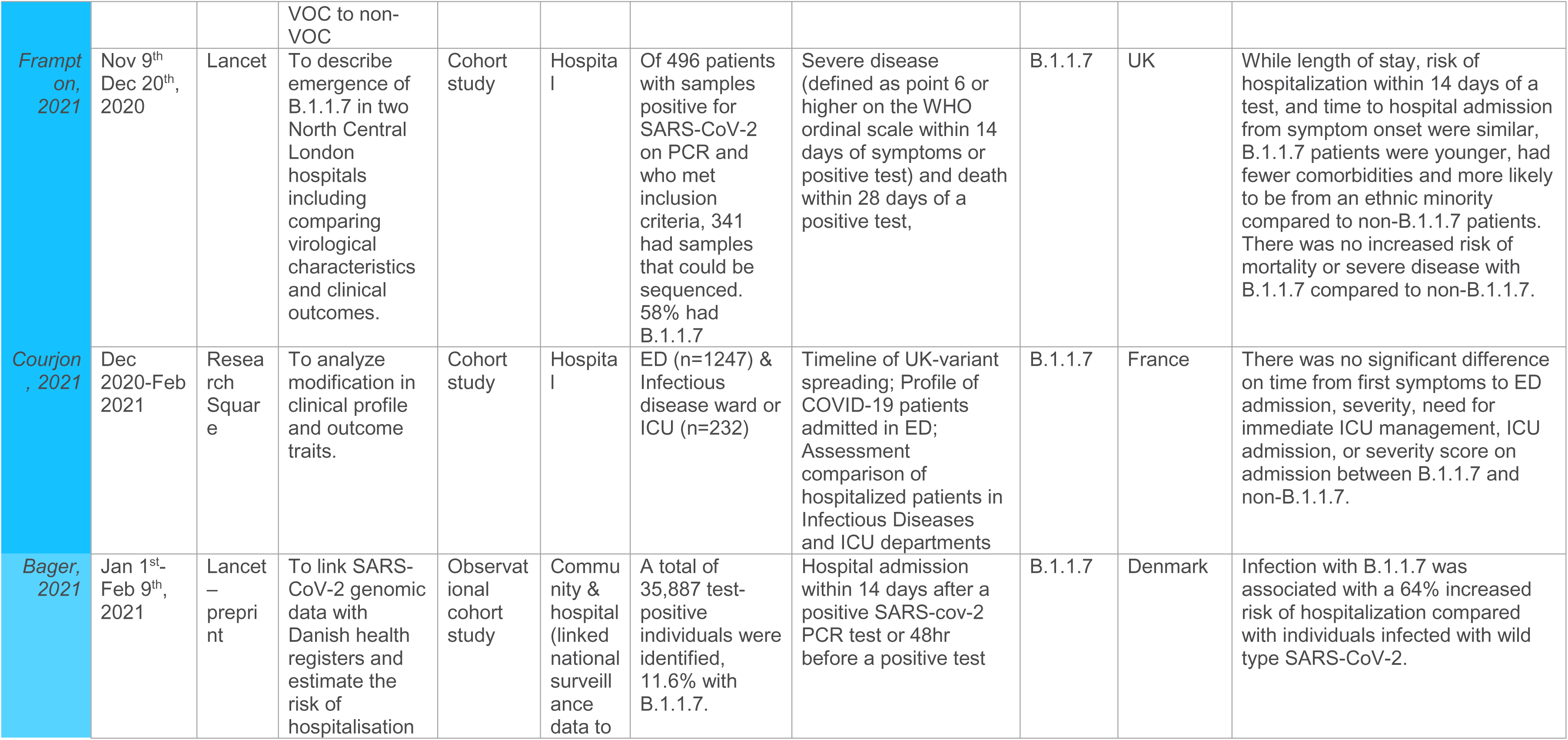
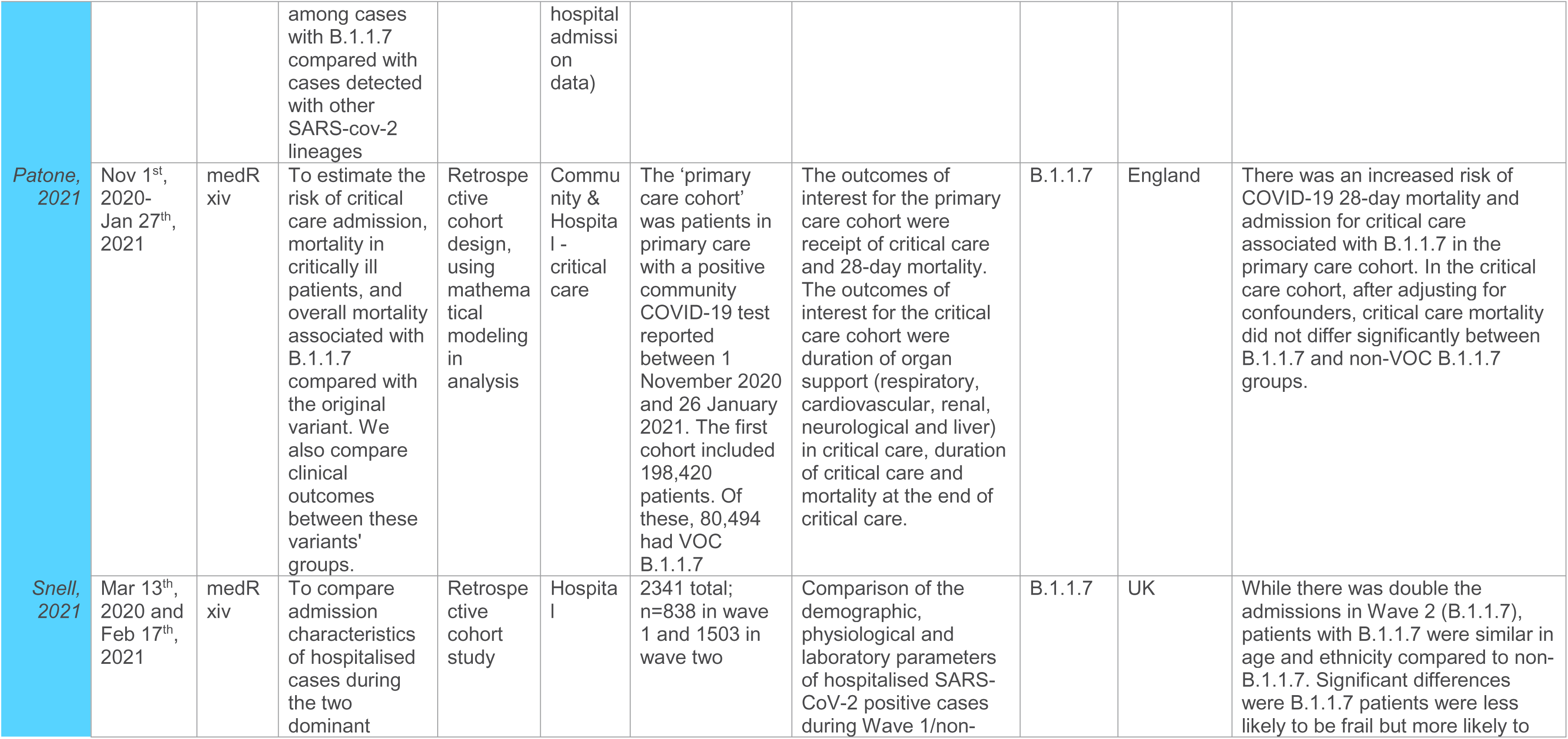
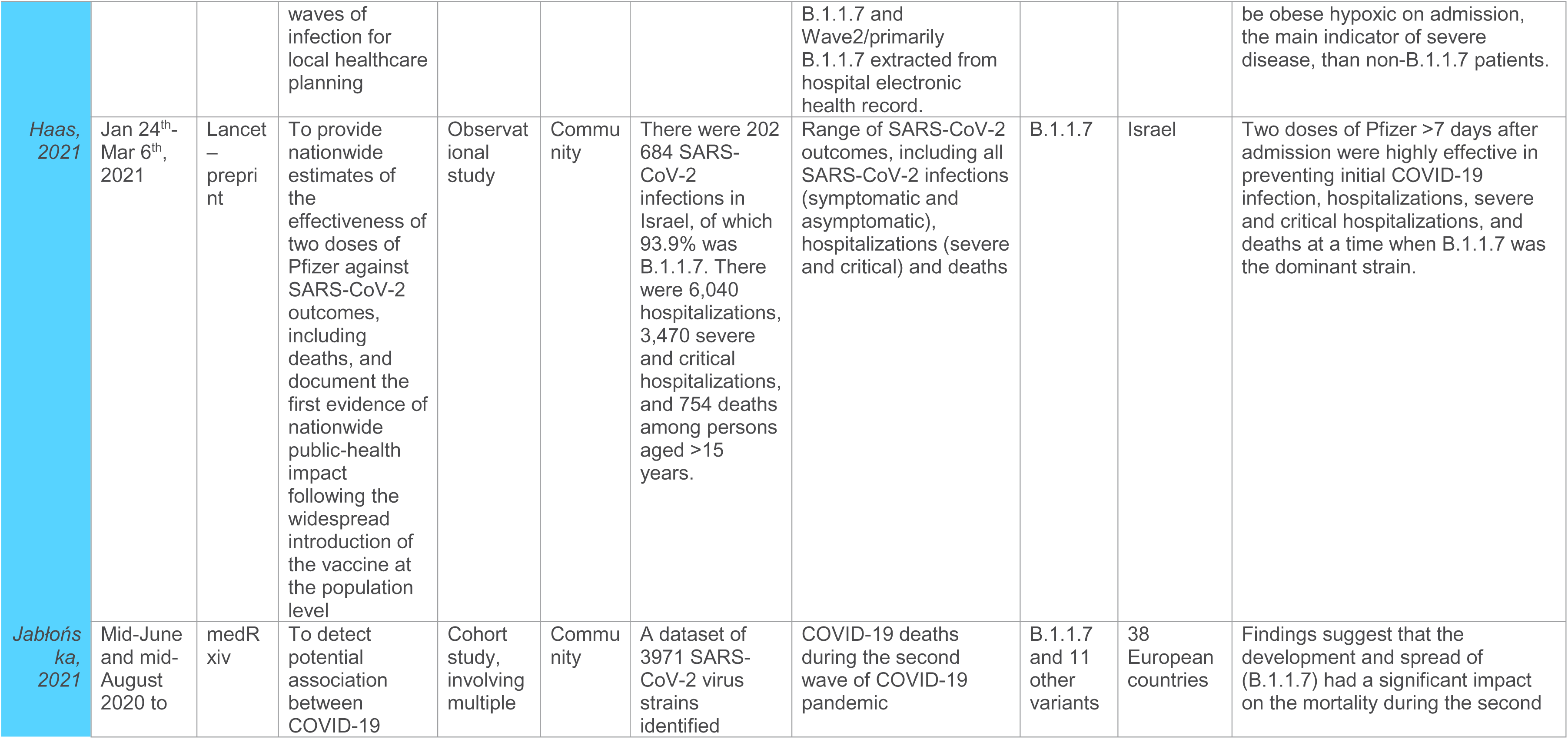
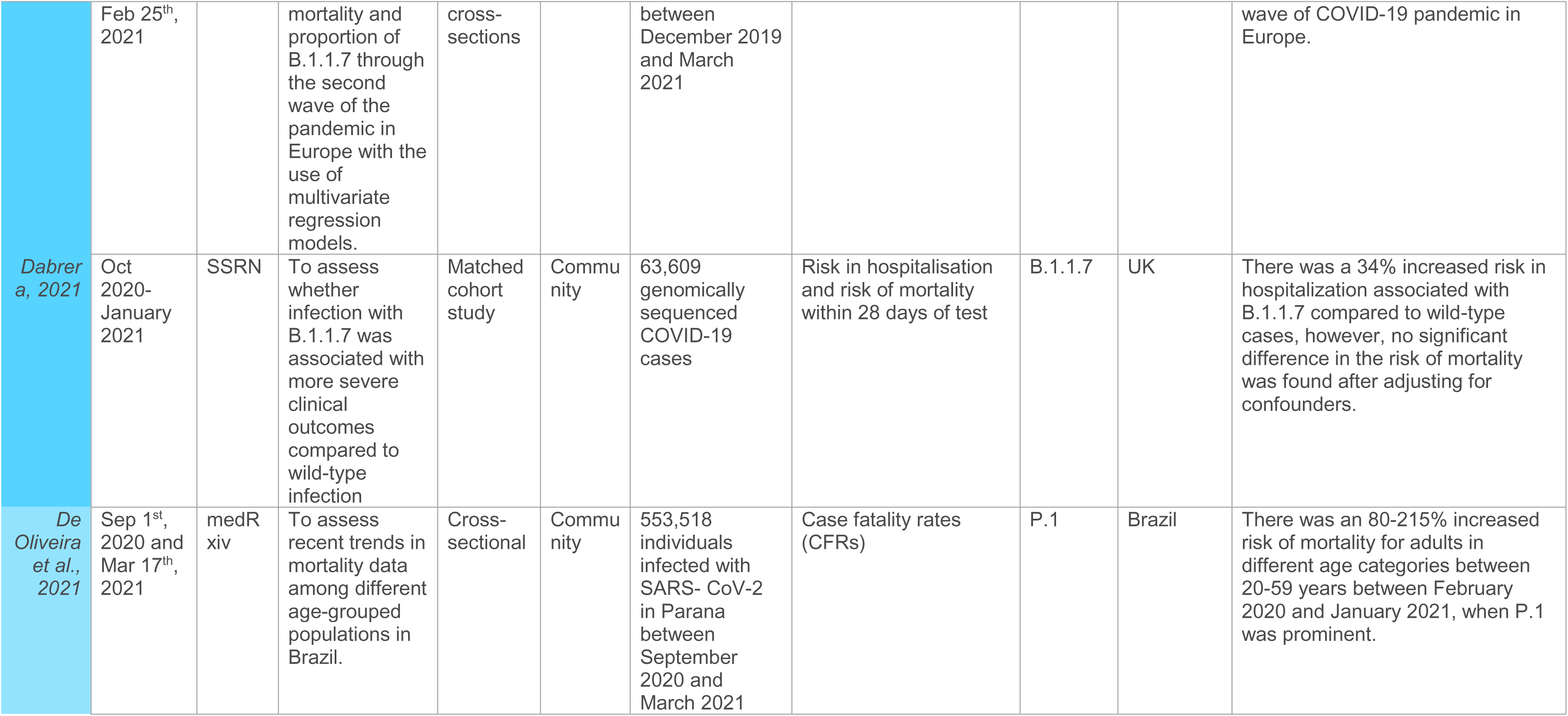
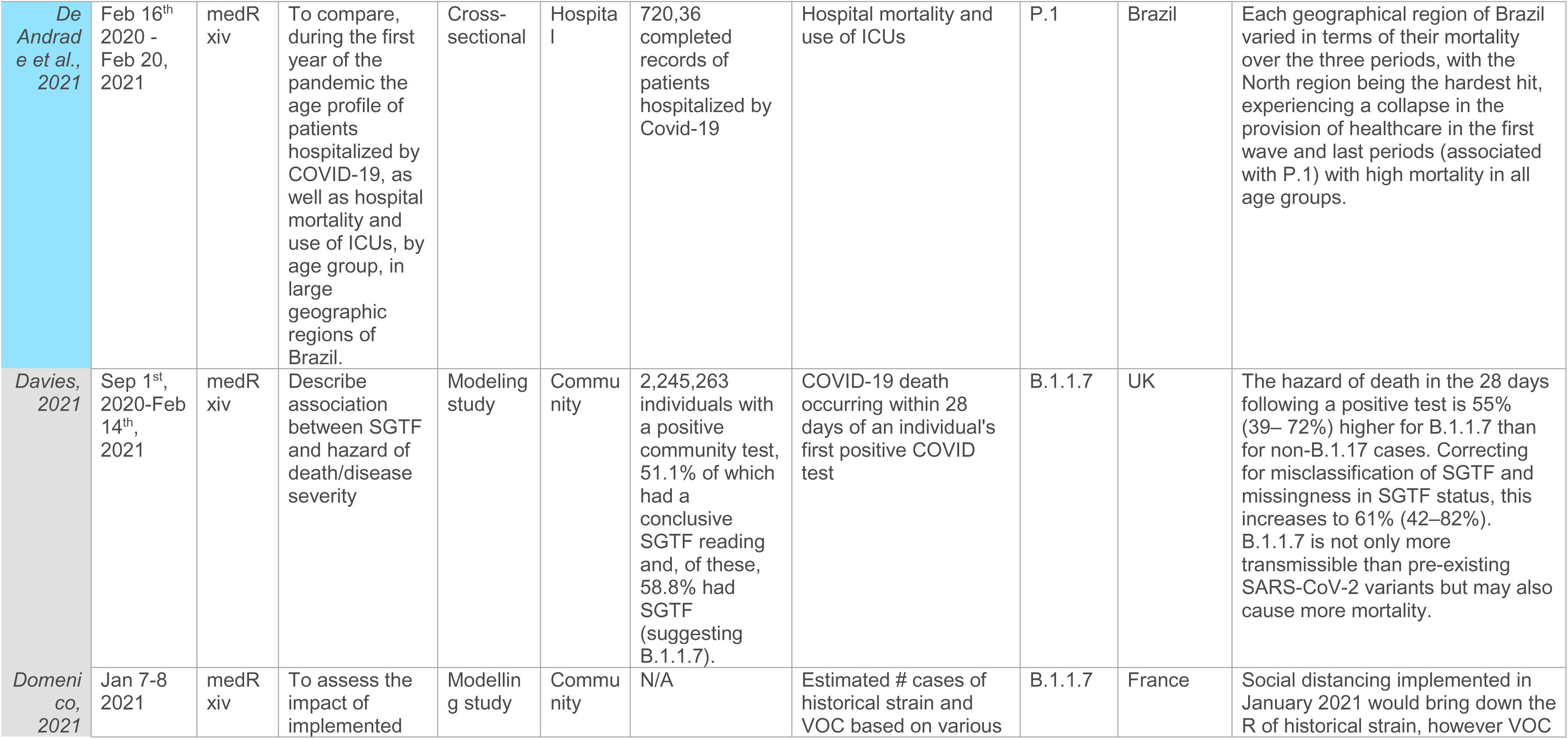
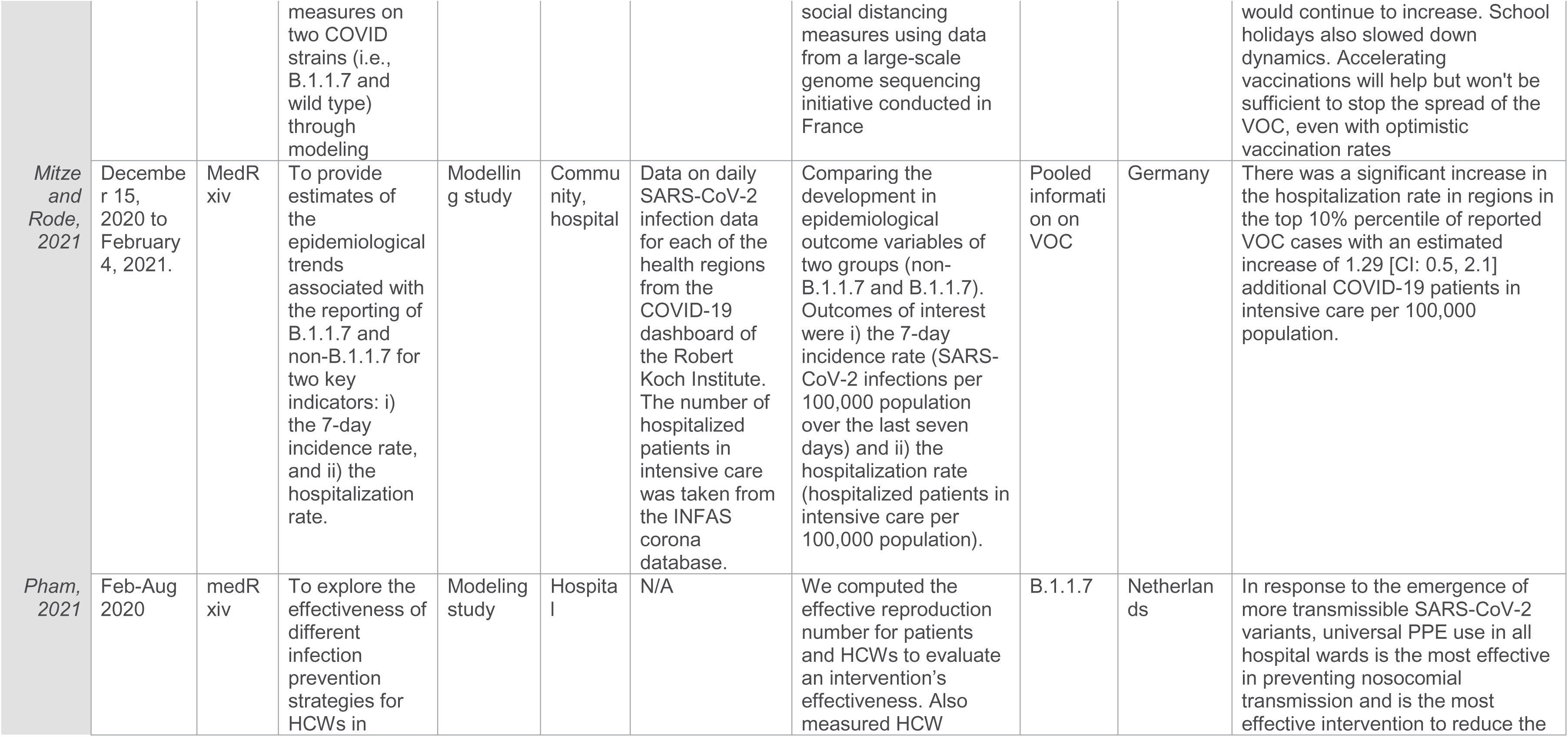
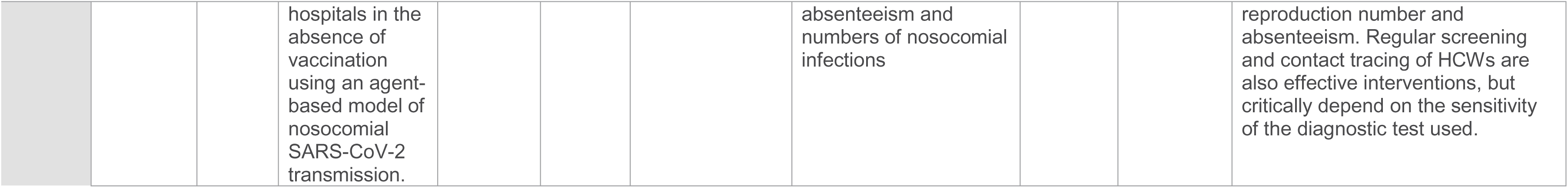
Summary of Health Systems Sources

**Table 3:**
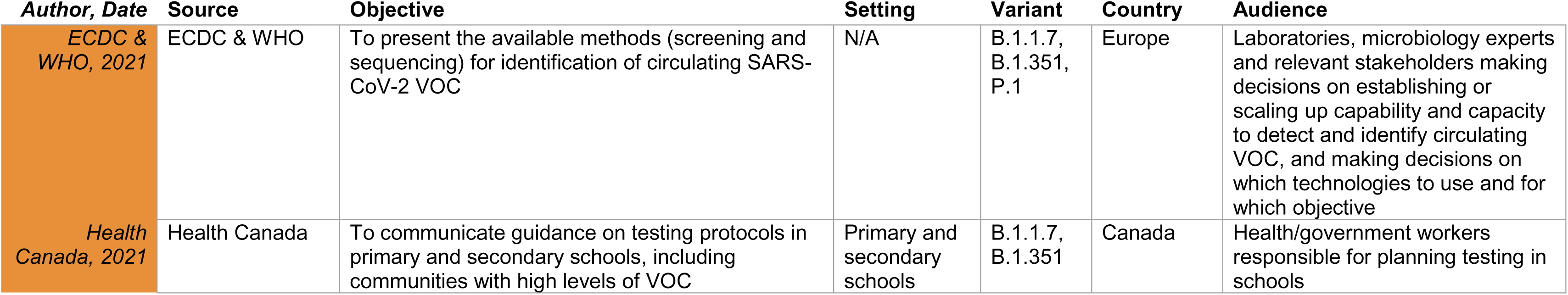
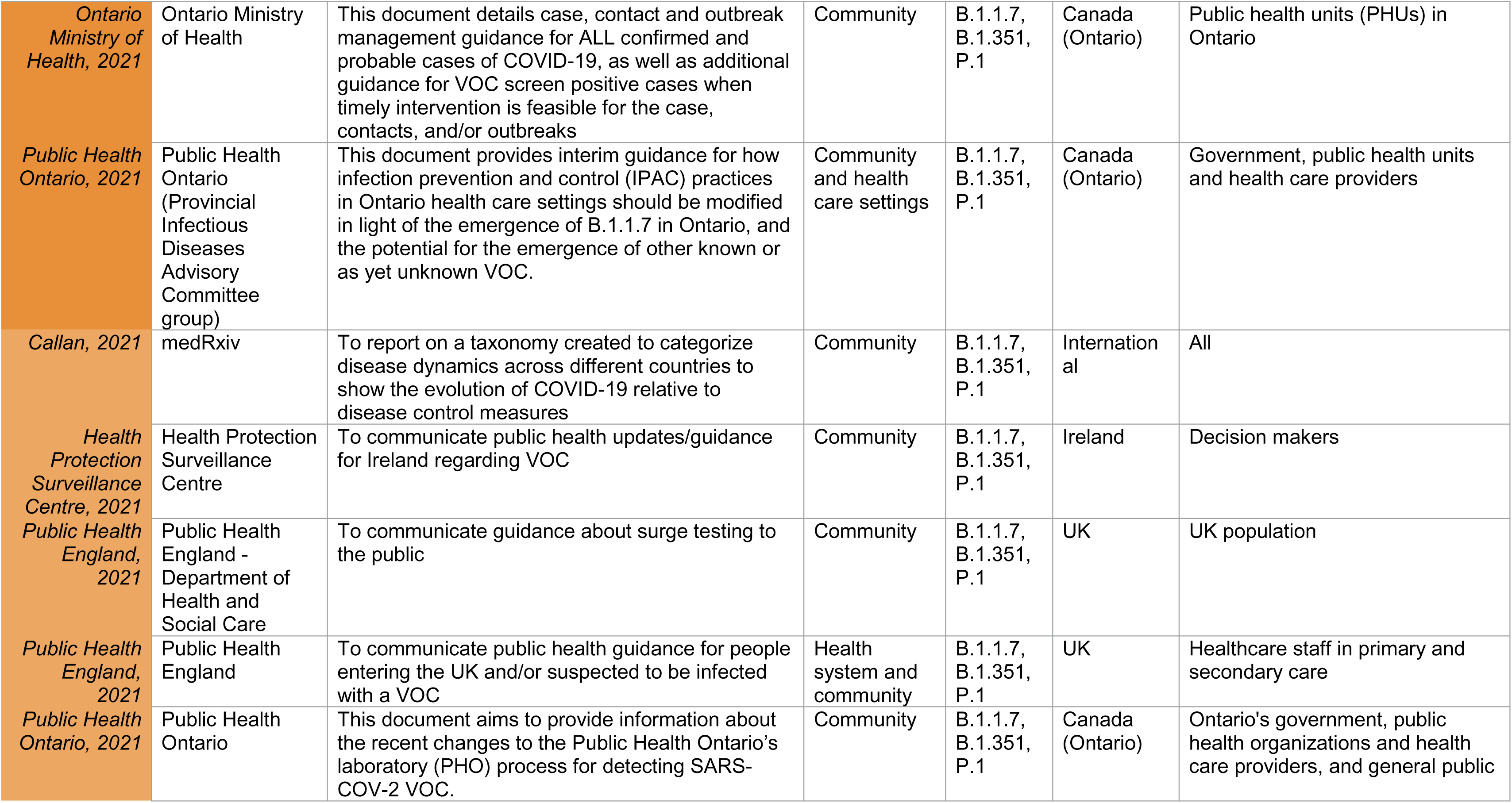
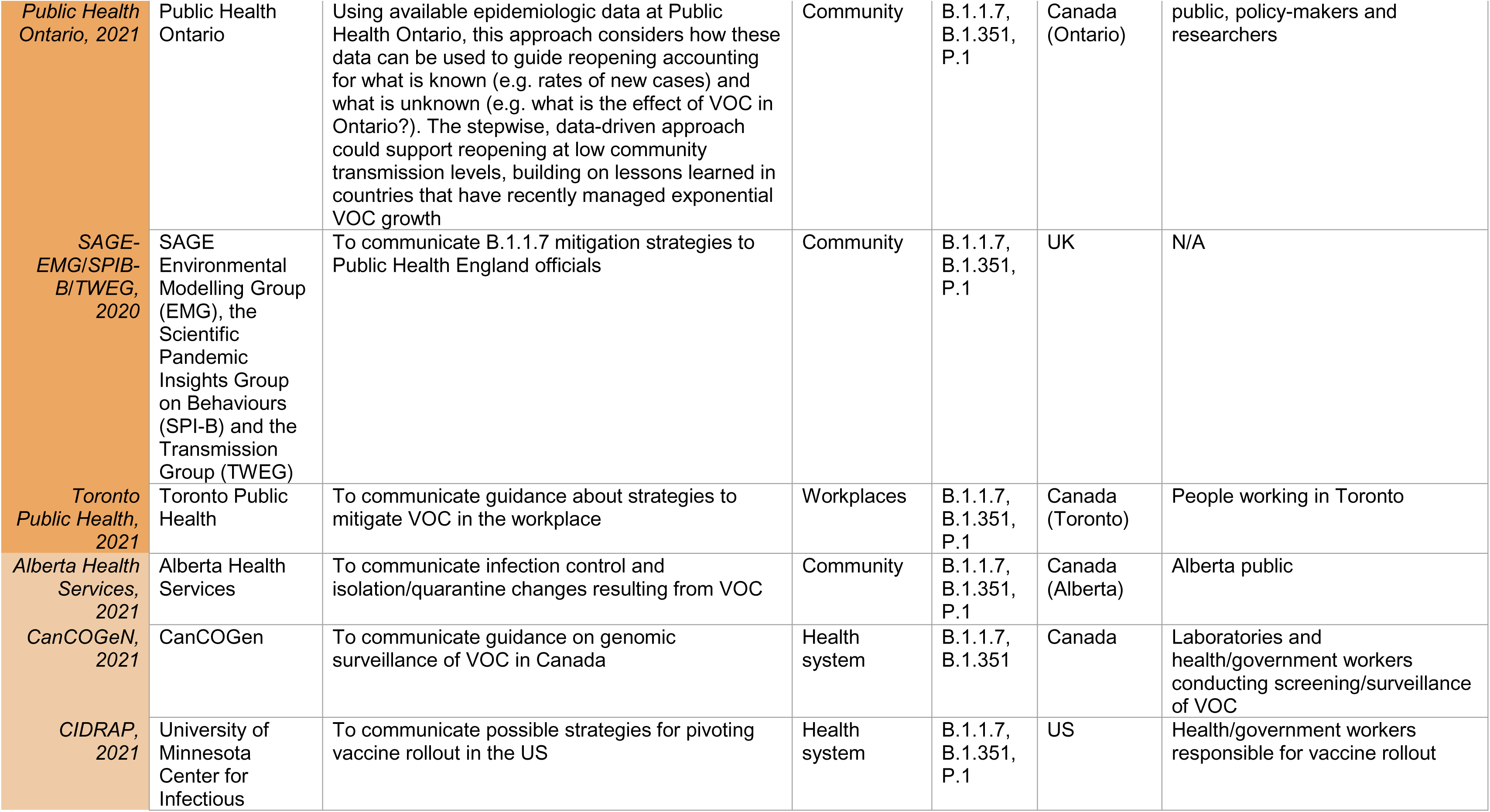
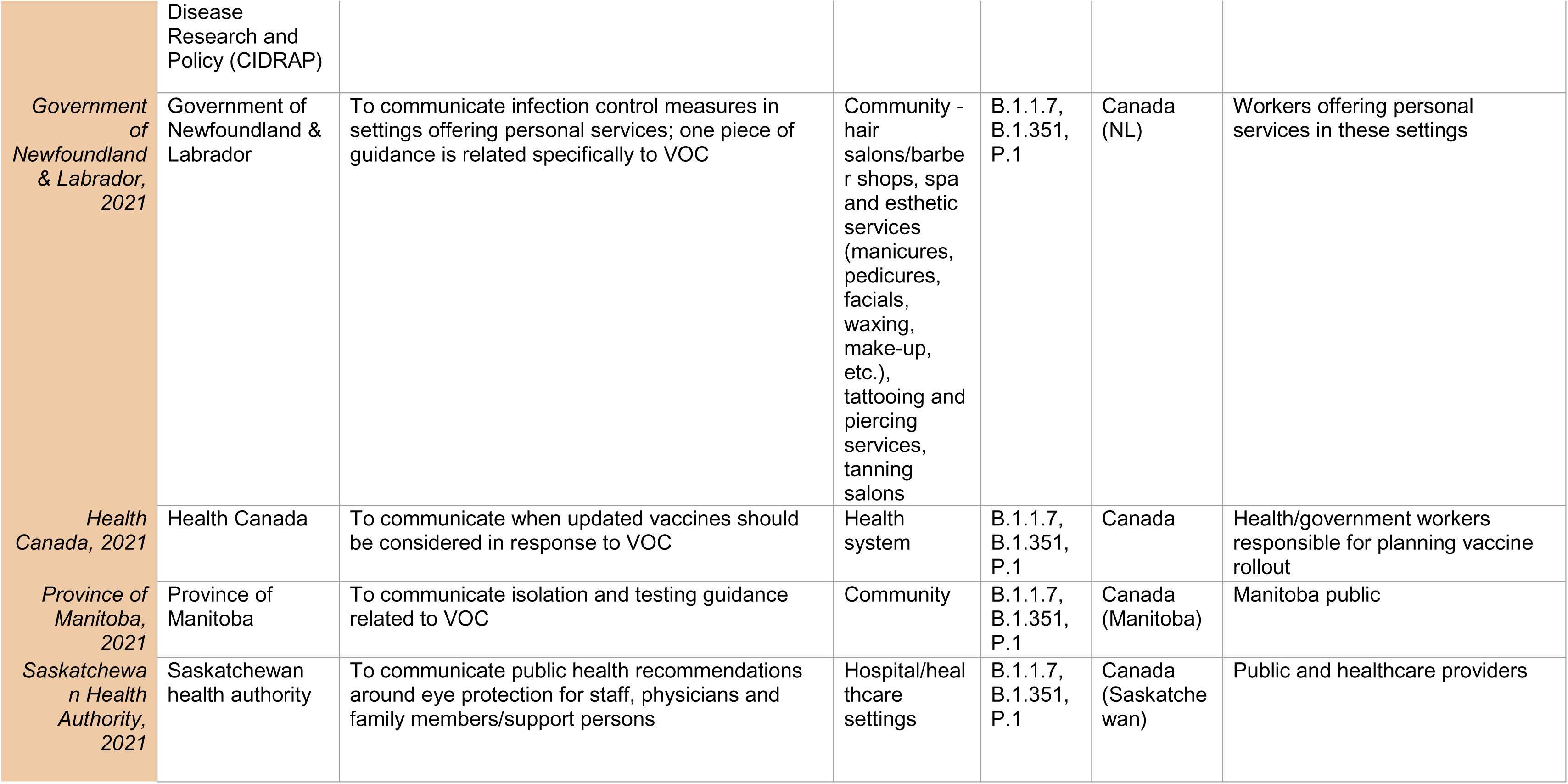
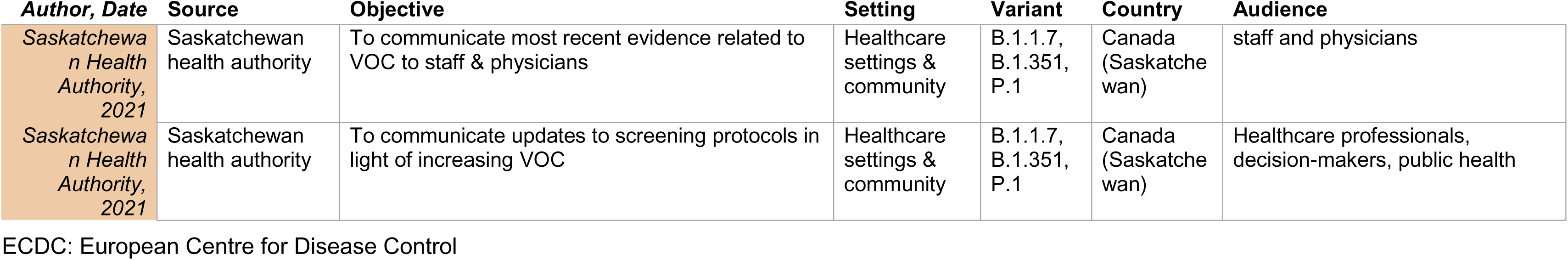
Summary of Guidance Documents

## Appendix 4 Quality Appraisal

**Table 1.** Quality Appraisal of research articles using the Newcastle-Ottawa scale (NOS) tool

| Author, year | Pre-print (PP) or Peer Review (PR) | Source | Average score per category |  |  | Minus 2* if preprint | Total Score (%) out of 9 | Overall Quality* (low, medium, high) |
| --- | --- | --- | --- | --- | --- | --- | --- | --- |
|  |  |  | Selection | Comparability | Outcome |  |  |  |
| Cohort Study Design |  |  |  |  |  |  |  |  |
| Bager, 2021 | PP | SSRN | 3.5 | 2 | 2.5 | -2 | 6 (67) | Medium |
| Buchan, 2021 | PP | MedRxiv | 3.5 | 2 | 2.5 | -2 | 6 (67) | Medium |
| Challen, 2021 | PR | BMJ | 4 | 2 | 3 | N/A | 9 (100) | High |
| Courjon, 2021 | PP | Research Square | 4 | 0.5 | 2.5 | -2 | 5 (56) | Medium |
| Dabrera, 2021 | PP | SSRN | 4 | 2 | 2 | -2 | 6 (67) | Medium |
| Frampton, 2021 | PR | Lancet | 4 | 2 | 3 | N/A | 9 (100) | High |
| Grint, 2021 | PR | Eurosurveillance | 4 | 2 | 3 | N/A | 9 (100) | High |
| Haas, 2021 | PP | SSRN | 4 | 2 | 2 | -2 | 6 (67) | Medium |
| Jablonska, 2021 | PP | MedRxiv | 4 | 1 | 2 | -2 | 5 (56) | Medium |
| Lumley, 2021 | PP | MedRxiv | 3 | 2 | 1 | -2 | 4 (44) | Low |
| Patone, 2021 | PP | MedRxiv | 4 | 2 | 2 | -2 | 6 (67) | Medium |
| Cross-sectional Study Design |  |  |  |  |  |  |  |  |
|  |  |  | Selection | Comparability | Outcome |  |  |  |
| *Bachtiger, 2021* | PP | MedRxiv | 2 | 1 | 0 | -2 | 1 (10) | Low |
| *DeAndrade, 2021* | PP | MedRxiv | 2.5 | 1 | 2.5 | -2 | 4 (40) | Low |
| *DeOliveira, 2021* | PP | MedRxiv | 2 | 0 | 2 | -2 | 2 (20) | Low |
| *Graham, 2021* | PR | Lancet | 3 | 2 | 3 | N/A | 8 (80) | High |
| *Snell, 2021* | PP | MedRxiv | 3 | 1 | 3 | -2 | 5 (50) | Medium |

\*High quality: 80-100%; Medium quality: 50-80%; Low quality: <50%

**Table 2.** Quality Appraisal of guideline documents using the AGREE II tool

| Author/Organization, Year | Domain Scores (%) |  |  |  |  |  | Domain 7: Overall Quality (%)* | Overall Quality (low, medium, high)** |
| --- | --- | --- | --- | --- | --- | --- | --- | --- |
|  | 1 | 2 | 3 | 4 | 5 | 6 |  |  |
| *Alberta Health Services, 2021* | 30.5 | 13.9 | 0.0 | 72.2 | 16.7 | 0.0 | 25.0 | Low |
| *Callan, 2021* | 13.9 | 13.9 | 11.5 | 47.2 | 33.3 | 25.0 | 50.0 | Medium |
| *CanCOGeN, 2021* | 18.8 | 25.0 | 5.2 | 38.9 | 18.8 | 62.5 | 25.0 | Low |
| *CIDRAP, 2021* | 44.4 | 41.7 | 13.5 | 33.3 | 20.8 | 8.3 | 41.7 | Low |
| *ECDC & WHO, 2021* | 86.1 | 80.5 | 35.4 | 52.8 | 41.7 | 12.5 | 66.7 | High |
| *Government of Newfoundland and Labrador, 2021* | 44.4 | 25.0 | 0.0 | 63.9 | 20.8 | 0.0 | 16.7 | Low |
| *Health Canada, 2021* | 36.1 | 27.8 | 8.3 | 33.3 | 29.2 | 16.7 | 16.7 | Low |
| *Health Canada, 2021* | 44.4 | 63.9 | 22.9 | 69.4 | 47.9 | 16.7 | 66.7 | High |
| *Health Protection Surveillance Centre, 2021* | 44.4 | 25.0 | 12.5 | 44.4 | 37.5 | 41.7 | 58.3 | Medium |
| *Ontario Ministry of Health, 2021* | 58.3 | 47.2 | 22.9 | 80.6 | 35.4 | 16.7 | 75.0 | High |
| *Province of Manitoba, 2021* | 44.4 | 25.0 | 2.0 | 38.9 | 14.6 | 0.0 | 25.0 | Low |
| *Public Health England, 2021* | 47.2 | 22.2 | 3.1 | 75.0 | 20.8 | 16.7 | 58.3 | Medium |
| *Public Health England, 2021* | 86.1 | 50.0 | 3.1 | 63.9 | 27.1 | 8.3 | 50.0 | Medium |
| *Public Health Ontario, 2021* | 72.2 | 69.4 | 24.0 | 77.8 | 35.4 | 8.3 | 83.3 | High |
| *Public Health Ontario, 2021* | 69.4 | 47.2 | 8.3 | 55.6 | 25.0 | 16.7 | 41.7 | Medium |
| *Public Health Ontario, 2021* | 69.4 | 44.4 | 18.7 | 52.8 | 37.5 | 16.7 | 41.7 | Medium |
| *SAGE-EMG/SPIB-B/TWEG, 2020* | 27.8 | 27.8 | 22.9 | 55.6 | 29.2 | 12.5 | 58.3 | Medium |

| **Author/Organization, Year** | **Domain Scores (%)** |  |  |  |  |  | **Domain 7:**<br>**Overall Quality**<br>**(%)\*** | **Overall Quality (low,**<br>**medium, high)\*\*** |
| --- | --- | --- | --- | --- | --- | --- | --- | --- |
|  | 1 | 2 | 3 | 4 | 5 | 6 |  |  |
| *Saskatchewan Health Authority, 2021* | 8.3 | 8.3 | 3.1 | 50.0 | 10.4 | 0.0 | 25.0 | Low |
| *Saskatchewan Health Authority, 2021* | 44.4 | 22.2 | 4.2 | 47.2 | 6.2 | 8.3 | 16.7 | Low |
| *Saskatchewan Health Authority, 2021* | 25.0 | 19.4 | 2.1 | 44.4 | 4.2 | 4.2 | 16.7 | Low |
| *Toronto Public Health, 2021* | 41.7 | 30.6 | 3.1 | 75.0 | 41.7 | 8.3 | 58.3 | Medium |

\*Subjective overall score calculated as an average between two appraiser scores
\*\*Low: Low overall score & appraisers do not recommend use; Medium: middle overall score and/or appraisers have mixed views on whether guideline is recommended; High: middle to high score & appraisers both recommend guideline

